# Is there an anticholinergic effect of drugs beyond polypharmacy? A simulation study on death, dementia, and delirium in UK Biobank

**DOI:** 10.1101/2024.08.06.24311533

**Authors:** J. Mur, L. E. Stirland, G. Muniz-Terrera, A. K. Leist

## Abstract

The use of anticholinergic drugs has been associated with adverse health outcomes. However, their effects cannot be completely separated from the effects of general polypharmacy using standard methods. The objective of this study was to explore the extent to which the detrimental health effects attributed to anticholinergic burden measured by anticholinergic burden scales (ABS) were distinct from those of polypharmacy. We compared observed effects of ABS against simulated effects of generated pseudoscales intended to measure polypharmacy using UK Biobank primary care data. We randomly sampled from 525 anticholinergic and non-anticholinergic drugs prescribed in the year 2015 to ∼200,000 participants with an average age of 65 years. We then created 1,000 pseudoscales, the score of which was designed to represent the strength of the background effect of polypharmacy, differentiating pseudoscales constructed to capture either general polypharmacy or putative anticholinergic polypharmacy, and exhibiting similar distributional properties to 23 real-world ABS (statistical equivalence). We performed individual logistic regressions for each scale to estimate associations between ABS scales and pseudoscales, respectively, and risk of death, dementia, or delirium. Across outcomes, odds ratios for anticholinergic-polypharmacy pseudoscales were on average 0.03-0.05 greater than those of general-polypharmacy pseudoscales. The number of drugs composing the scales was correlated with the size of adverse effects for both pseudoscales (r=∼0.5, p<0.001) and ABS (r=∼0.7, p<0.001). In total, 50-90% of ABS showed stronger effects than the majority of pseudoscales. ABS exhibited stronger associations with the studied adverse health outcomes than would be expected from polypharmacy alone (range of differences in odds ratios: -0.05 to 0.20). Most existing ABS capture more variance in the association with death, dementia, and delirium than polypharmacy alone, but with varying degrees of strength.

## Introduction

Anticholinergic burden scales (ABS) assign numerical values to drugs based on their presumed propensities to block muscarinic receptors. The sum of an individual’s values yields a weighted burden score - the anticholinergic burden - that can be used by clinicians or researchers to identify patients or populations vulnerable to adverse side effects. Anticholinergic burden has been associated with adverse long-term health outcomes, including all-cause mortality^1^, dementia^2^, and delirium^3^.

Polypharmacy, the concurrent use of multiple medications, is also associated with negative health outcomes, including mortality and dementia^4,5^. We can divide polypharmacy into polypharmacy due to non-anticholinergic drugs (non-anticholinergic polypharmacy) and polypharmacy due to anticholinergic drugs (anticholinergic polypharmacy); the latter is effectively an unweighted ABS. Although some studies have attempted to discern the effects of non-anticholinergic polypharmacy and anticholinergic burden^6^ or of polypharmacy and anticholinergic burden^7^, collinearity prevents complete separation of the effects of anticholinergic polypharmacy and anticholinergic burden. Consequently, it is unknown whether some of the reported associations between anticholinergic use and health outcomes are specific to the presumed anticholinergic properties of these drugs or consequences of more general effects of polypharmacy.

In this study, we attempted to isolate the effect of anticholinergics from that of polypharmacy. We constructed burden scales, with scores assigned based on random sampling among commonly prescribed drugs (hereafter referred to as pseudoscales); the first set of pseudoscales sampled only among putative anticholinergic drugs to capture anticholinergic polypharmacy, the second set of pseudoscales sampled across all drugs to capture general polypharmacy. We then compared the distributions of effect sizes for the associations between the scores of the ‘real-world’ ABS with the risk of all-cause mortality, dementia, or delirium against the associations between the scores of pseudoscales and the risk across the three outcomes. The rationale of our study was that effect estimates of ABS higher than effects of random combinations of commonly prescribed drugs may capture ‘true’ effects due to anticholinergic burden beyond effects of polypharmacy.

## Methods

### Hypotheses

In the absence of a validated test of anticholinergic potency, anticholinergic drugs are defined as such and included in ABS partly based on their past associations with health outcomes. Thus, we first hypothesised that effect sizes of ‘real-world’ ABS and effect sizes of pseudoscales sampled among anticholinergic drugs (anticholinergic polypharmacy) would be on average greater than effect sizes of pseudoscales sampled among all drugs (general polypharmacy). Second, as described above, general polypharmacy is associated with negative health outcomes. Thus, we hypothesised that the number of drugs composing the pseudoscales would be positively associated with the magnitude of adverse health effects of these scales. Finally, we hypothesised that ‘real-world’ ABS – due to their inclusion of a weighting of anticholinergic drugs – would exhibit greater effects than those observed for either general or anticholinergic pseudoscales.

### Sample

UK Biobank is a cohort study of ∼500,000 participants from England, Scotland, and Wales, whose ages ranged from 37 to 73 years during the first assessment between the years 2006 and 2010^8,9^. Demographic and lifestyle information was acquired, and cognitive tests and blood-based diagnostics were administered. Data were linked to inpatient secondary hospital care (i.e., data on participants admitted to hospital overnight), death records, and - for about half of participants - to primary care. The latter includes diagnoses, clinical events, and prescriptions recorded by general practitioners (GPs).

### Prescription preparation and scale construction

Prescriptions in the sample were matched with generic drug names. Because the search for individual drugs in the sample and the quality control of the name matchings required manual work on a large dataset, we did not identify all drugs. Instead, we identified the most common character strings (>5,000 total occurrences), drugs listed on anticholinergic scales, and all existing combinations of the above drugs according to the British National Formulary (https://bnf.nice.org.uk/). In this way, ∼98.6% of prescriptions in the sample were assigned drug names.

For the assignment of anticholinergic potency, we used those ABS that were available as lists of medicines that assigned unambiguous numerical scores to each drug, thus updating a previous selection^10^. We identified 23 scales (**Suppl. Table 1**); drugs with ophthalmic, topic (including transdermal patches), otic, and nasal administration routes (∼12.5% of prescriptions) were assigned potency scores of 0, as previously^6,11-13^.

We used two different procedures to construct pseudoscales, ‘across-sampling’ and ‘within-sampling’. Across-sampling was intended to compare the effects of anticholinergic polypharmacy with those of general polypharmacy and yielded 1,000 pseudoscales. For each pseudoscale, the number of drugs to be included on the scale was determined by drawing from a uniform distribution with the minimum (n=15) and maximum (n=150) corresponding to the minimum and maximum numbers of drugs prescribed in the year 2015 that were scored as anticholinergic across the 23 studied ABS. Second, the probabilities for each drug to be assigned a potency score were determined based on how frequent these potency scores were across ABS (probabilities of assignment: 0.017, 0.25, 0.20, 0.52, and 0.009 for potency scores 4, 3, 2, 1, and 0.5, respectively). Within-sampling was intended to generate pseudoscales with equivalent properties to the individual ABS to account for correlations between effect sizes of scales with the count of drugs constituting them (see results). We generated 250 within-sampling pseudoscales for each of the 23 ABS. Each resulting pseudoscale had the same number of included drugs and the same frequency distribution of potency scores as its corresponding ‘real-world’ ABS.

Across-sampling and within-sampling were performed among all named drugs in the sample (general polypharmacy) and among those named drugs in the sample that were considered anticholinergic by at least one ABS (anticholinergic polypharmacy). The total number of drugs with valid routes of administration that could contribute to general and anticholinergic polypharmacy in the initial sample in the year 2015 was 525 and 214, respectively. The seed for the simulation was defined once at the start of each simulation set. We used the Mersenne Twister pseudorandom number generator from the *numpy* library^14^ in Python on the Windows system. The number of performed simulations was based on when the distributions of interest stabilised and not on predefined requirements to estimate the true effect to any specific degree of accuracy. Data preparation and analyses were performed in Python 3.11.5 and in R 4.3.2. The code used to clean and analyse the data is available at https://github.com/JuM24/Pseudoscales_simulation.

### Data preparation

We used pseudoscales to assign a potency score to each prescription. The initial sample consisted of 222,048 participants with primary care data and 57,691,961 prescriptions. We removed prescriptions with an assigned participant and date, but without prescription content, those issued before or on the date of birth, and those issued in the future, assuming that they were erroneous. This led to the exclusion of <1% of prescriptions (**Suppl. Figure 1**). We calculated annual burden scores and restricted the observation period to the year 2015. We chose this year to maximise the completeness of the primary care record which increases with time and ends at the date of extraction (years 2016/2017). This choice also maximised the age of the sample, as previous studies on this topic have mostly focused on older individuals. Periods of continuous primary-care ascertainment were inferred as previously described^15^.

### Causal structure and covariates

We formulated a causal structure for drug burden (**Suppl. Figure 2**), defining confounders as assumed common causes of exposure and outcome based on the previous literature on risk factors for dementia^16^, delirium^17,18^, and mortality^19^. All models were adjusted for the count of prescribed drugs not included in the exposure scale for that model to reflect residual polypharmacy (for ABS this reflected non-anticholinergic polypharmacy), age, data provider (computer system supplier to GP practices), sex, education, socioeconomic deprivation, alcohol consumption, waist circumference, smoking status, physical activity, and history of cerebrovascular disease. Depending on the outcome, the models could also include further covariates. For death: history of chronic lower respiratory disease, diabetes, liver disease, influenza/pneumonia, ischaemic heart disease, colon cancer, prostate/ovarian cancer, lung cancer, and breast cancer. For dementia: general cognitive ability, air pollution, hypertension, hypercholesterolemia, depressive mood, social isolation, loneliness, history of mood disorders, central nervous system disorders, diabetes, and hearing impairment. For delirium: general cognitive ability, social isolation, loneliness, history of psychotic disorders, mood disorders, visual impairment, hearing impairment, sleep disorders, endocrinopathy, nutritional deficiencies, and metabolic disorders. The history of each disorder except hearing impairment was based on a combination of self-report, hospital, and death records as recorded in the UK Biobank first occurrences or algorithmically-defined outcome categories^20,21^. Hearing impairment was based on a custom algorithm that accounted for health records, self-report, and objective hearing-in-noise tests administered during the UK Biobank assessment^22^. To ascertain a single measure of air pollution, we used the first principal component for the measurements of nitrogen dioxide, nitrogen oxide, and particulate matter (pm_2.5_). Due to a lack of data on pollution for participants from Scotland, these participants (∼11% of the sample) were excluded from the analyses of dementia. Social isolation, loneliness, and depressed mood were based on self-report. Cognitive ability was derived from tests of executive functioning and reaction time^10,23^. Further details are in **Suppl. Tables 2, 3, 4** and **Suppl. Text 1**.

### Modelling

We estimated effect sizes using logistic regression. Due to the high number of models, we randomly selected a subset of models (<0.5%) that were tested for assumptions. Burden scores generally did not show linear relationships with the log odds. Moreover, large residuals were often disproportionately represented among participants who experienced the outcome. Because of this, before running the models, we applied Synthetic Minority Oversampling Technique (SMOTE), which changes the distribution of the outcome by creating synthetic examples of the minority class^24^. We increased the number of observations in the minority group to achieve one third of the number of observations present in the majority group. This led to a decrease in the observed imbalance of residuals. Drug burden scores and general polypharmacy assessed as counts of prescribed drugs sometimes exhibited unusually high values (>4 standard deviations above the mean) and were removed as outliers. The effect sizes are expressed as odds ratios (OR). The overlap between distributions of ORs was calculated by the kernel density estimation using the Sheather-Jones method^25^. We performed a sensitivity analysis using the period from the year 2004 to 2006 instead of 2015, assuming that result patterns would be similar but would show overall smaller effects due to the lower age of the sample.

## Results

Among the ∼160,000-190,000 participants (numbers differ between models), ∼4.8% died, ∼1.2% were diagnosed with dementia, and ∼1.1% with delirium (**Table 1, Suppl. Table 5**). In the year 2015, between 10% and 45% of participants – depending on the scale – were prescribed at least one anticholinergic drug. Among these participants, the median anticholinergic burden ranged from 4 to 18 across scales (**Suppl. Table 1**). Effects of the 23 ‘real-world’ ABS-derived scores ranged from 1.01 to 1.17, 0.99 to 1.23, and 0.98 to 1.21 on the risk of death, dementia, and delirium, respectively (**Suppl. Tables 7-9**). In across-sampling, the ORs for general pseudoscales ranged from 0.92 to 1.22 (death), from 0.91 to 1.36 (dementia), and from 0.87 to 1.28 (delirium). ORs for anticholinergic pseudoscales ranged from 0.96 to 1.24 (death), from 0.92 to 1.41 (dementia), and from 0.92 to 1.26 (delirium). The 95% simulation intervals (SI) that express the values in which 95% of ORs are situated were of similar magnitude for both effects of general and anticholinergic pseudoscales (**Table 2**). The distributions of ORs for general and anticholinergic polypharmacy exhibited an overlap of 0.76, 0.70, and 0.78 for death, dementia, and delirium, respectively (**Figure 1**).

**Table 1:**
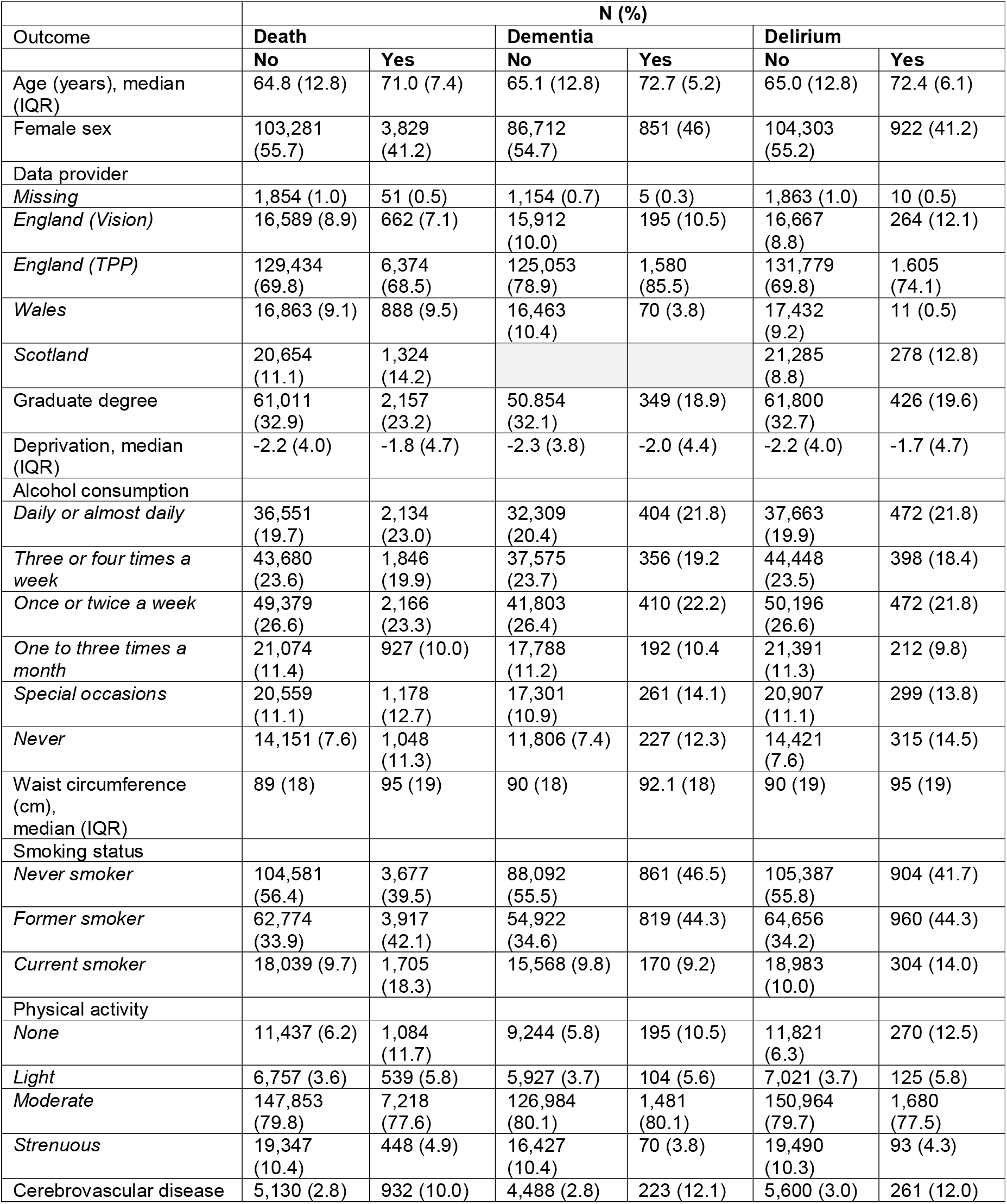
descriptive statistics for covariates used in models of all outcomes. For the descriptive statistics of other covariates specific to each outcome, see **Suppl. Table 5**. The below numbers hold for the sample before the removal of outliers of burden scales and before application of SMOTE.

| Outcome | N (%) |  |  |  |  |  |
| --- | --- | --- | --- | --- | --- | --- |
|  | Death |  | Dementia |  | Delirium |  |
|  | No | Yes | No | Yes | No | Yes |
| Age (years), median (IQR) | 64.8 (12.8) | 71.0 (7.4) | 65.1 (12.8) | 72.7 (5.2) | 65.0 (12.8) | 72.4 (6.1) |
| Female sex | 103,281 (55.7) | 3,829 (41.2) | 86,712 (54.7) | 851 (46) | 104,303 (55.2) | 922 (41.2) |
| Data provider |  |  |  |  |  |  |
| Missing | 1,854 (1.0) | 51 (0.5) | 1,154 (0.7) | 5 (0.3) | 1,863 (1.0) | 10 (0.5) |
| England (Vision) | 16,589 (8.9) | 662 (7.1) | 15,912 (10.0) | 195 (10.5) | 16,667 (8.8) | 264 (12.1) |
| England (TPP) | 129,434 (69.8) | 6,374 (68.5) | 125,053 (78.9) | 1,580 (85.5) | 131,779 (69.8) | 1,605 (74.1) |
| Wales | 16,863 (9.1) | 888 (9.5) | 16,463 (10.4) | 70 (3.8) | 17,432 (9.2) | 11 (0.5) |
| Scotland | 20,654 (11.1) | 1,324 (14.2) |  |  | 21,285 (8.8) | 278 (12.8) |
| Graduate degree | 61,011 (32.9) | 2,157 (23.2) | 50,854 (32.1) | 349 (18.9) | 61,800 (32.7) | 426 (19.6) |
| Deprivation, median (IQR) | -2.2 (4.0) | -1.8 (4.7) | -2.3 (3.8) | -2.0 (4.4) | -2.2 (4.0) | -1.7 (4.7) |
| Alcohol consumption |  |  |  |  |  |  |
| Daily or almost daily | 36,551 (19.7) | 2,134 (23.0) | 32,309 (20.4) | 404 (21.8) | 37,663 (19.9) | 472 (21.8) |
| Three or four times a week | 43,680 (23.6) | 1,846 (19.9) | 37,575 (23.7) | 356 (19.2) | 44,448 (23.5) | 398 (18.4) |
| Once or twice a week | 49,379 (26.6) | 2,166 (23.3) | 41,803 (26.4) | 410 (22.2) | 50,196 (26.6) | 472 (21.8) |
| One to three times a month | 21,074 (11.4) | 927 (10.0) | 17,788 (11.2) | 192 (10.4) | 21,391 (11.3) | 212 (9.8) |
| Special occasions | 20,559 (11.1) | 1,178 (12.7) | 17,301 (10.9) | 261 (14.1) | 20,907 (11.1) | 299 (13.8) |
| Never | 14,151 (7.6) | 1,048 (11.3) | 11,806 (7.4) | 227 (12.3) | 14,421 (7.6) | 315 (14.5) |
| Waist circumference (cm), median (IQR) | 89 (18) | 95 (19) | 90 (18) | 92.1 (18) | 90 (19) | 95 (19) |
| Smoking status |  |  |  |  |  |  |
| Never smoker | 104,581 (56.4) | 3,677 (39.5) | 88,092 (55.5) | 861 (46.5) | 105,387 (55.8) | 904 (41.7) |
| Former smoker | 62,774 (33.9) | 3,917 (42.1) | 54,922 (34.6) | 819 (44.3) | 64,656 (34.2) | 960 (44.3) |
| Current smoker | 18,039 (9.7) | 1,705 (18.3) | 15,568 (9.8) | 170 (9.2) | 18,983 (10.0) | 304 (14.0) |
| Physical activity |  |  |  |  |  |  |
| None | 11,437 (6.2) | 1,084 (11.7) | 9,244 (5.8) | 195 (10.5) | 11,821 (6.3) | 270 (12.5) |
| Light | 6,757 (3.6) | 539 (5.8) | 5,927 (3.7) | 104 (5.6) | 7,021 (3.7) | 125 (5.8) |
| Moderate | 147,853 (79.8) | 7,218 (77.6) | 126,984 (80.1) | 1,481 (80.1) | 150,964 (79.7) | 1,680 (77.5) |
| Strenuous | 19,347 (10.4) | 448 (4.9) | 16,427 (10.4) | 70 (3.8) | 19,490 (10.3) | 93 (4.3) |
| Cerebrovascular disease | 5,130 (2.8) | 932 (10.0) | 4,488 (2.8) | 223 (12.1) | 5,600 (3.0) | 261 (12.0) |

**Table 2:** comparisons between general and anticholinergic pseudoscales in their respective effects of association with the outcomes. The rows depict for all three outcomes the 95% SI, mean and standard deviation (SD) of the OR, and the results of the paired T-test of the difference between the means of the ORs of general and anticholinergic pseudoscales.

| Outcome<br>Pseudoscale<br>type | Death |  | Dementia |  | Delirium |  |
| --- | --- | --- | --- | --- | --- | --- |
|  | general | antichol. | general | antichol. | general | antichol. |
| 95% OR SI | 0.97-1.16 | 0.99-1.20 | 0.96-1.21 | 0.96-1.29 | 0.94-1.18 | 0.96-1.22 |
| Mean (SD) | 1.07 | 1.10 | 1.06 | 1.11 | 1.06 | 1.09 |
| SD | 0.047 | 0.055 | 0.065 | 0.094 | 0.059 | 0.066 |
| T-test<br>(T, p) | 16.9, <0.001 |  | 17.4, <0.001 |  | 13.8, <0.001 |  |

**Figure 1:**
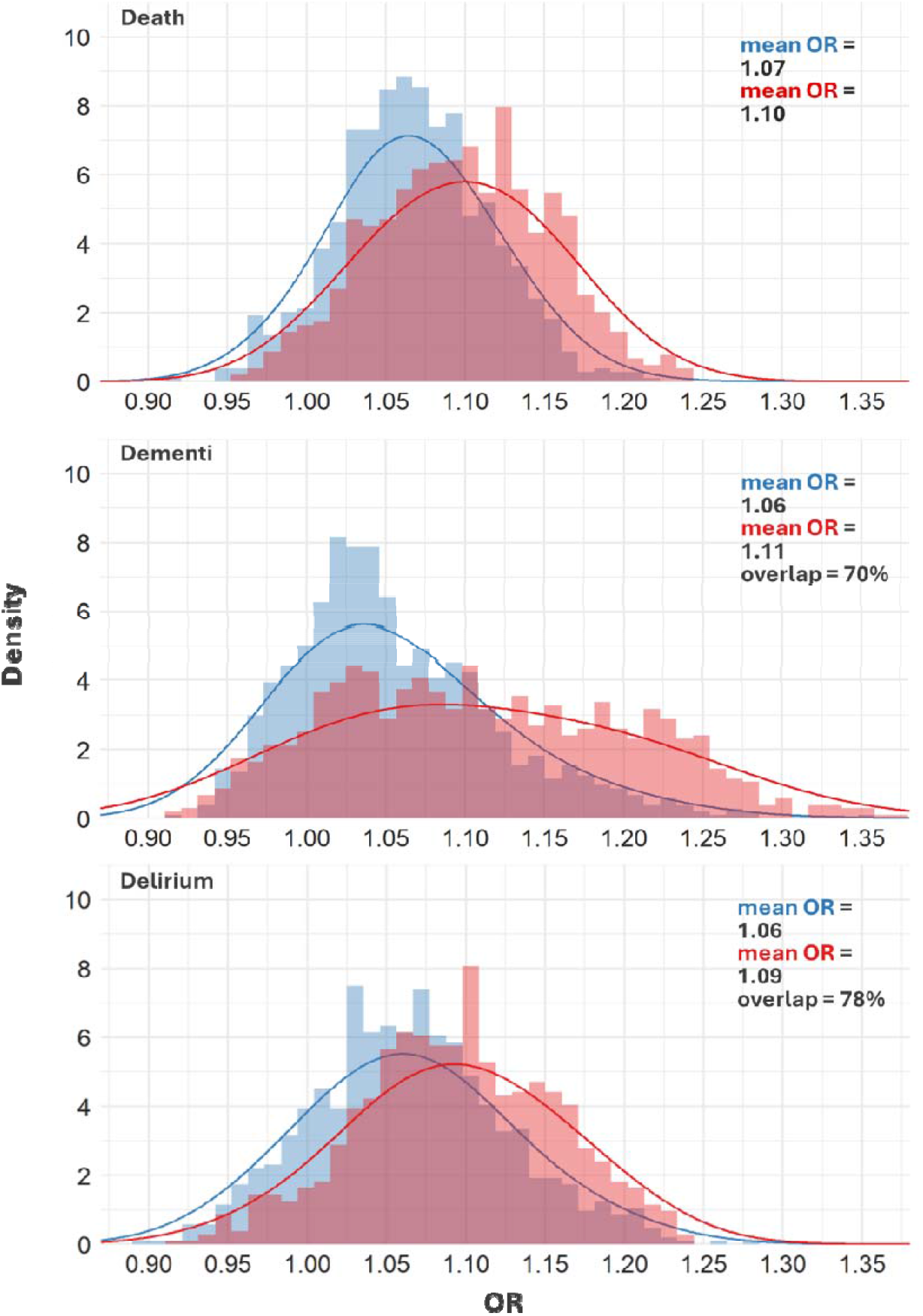
histograms and density plots for effects of general (**blue**) and anticholinergic (**red**) across-sampling pseudoscales when estimating the effect of drug burden on death, dementia, or delirium.

Anticholinergic pseudoscales exhibited larger effect sizes than general pseudoscales (**Table 2**). At p=0.05, the association between drug burden and the outcome would be significantly positive for 896/1000, 753/1000, and 815/1000 general pseudoscales for death, dementia, and delirium respectively, i.e., between 75.3% and 89.6% of tests for effects of pseudoscales across outcomes would be significantly greater than zero. For anticholinergic pseudoscales, these numbers were 946/1000, 852/1000, and 883/1000. Thus, the associations between drug burden and the outcome were significant in most tests, with a 6% (death), 13% (dementia), and 8% (delirium) higher probability of being significant if the burden scale was constructed using only putatively anticholinergic drugs as opposed to all drugs. The OR correlated with the scale size (i.e., count of number of drugs included on a scale) for all types of scales but was greatest for ABS (r=0.83 (95% CI=0.63-0.93), r=0.65 (95% CI=0.33-0.84), and r=0.79 (95% CI=0.56-0.91) for death, dementia, and delirium, respectively. This correlation was smaller for pseudoscales (**Figure 2**).

**Figure 2:**
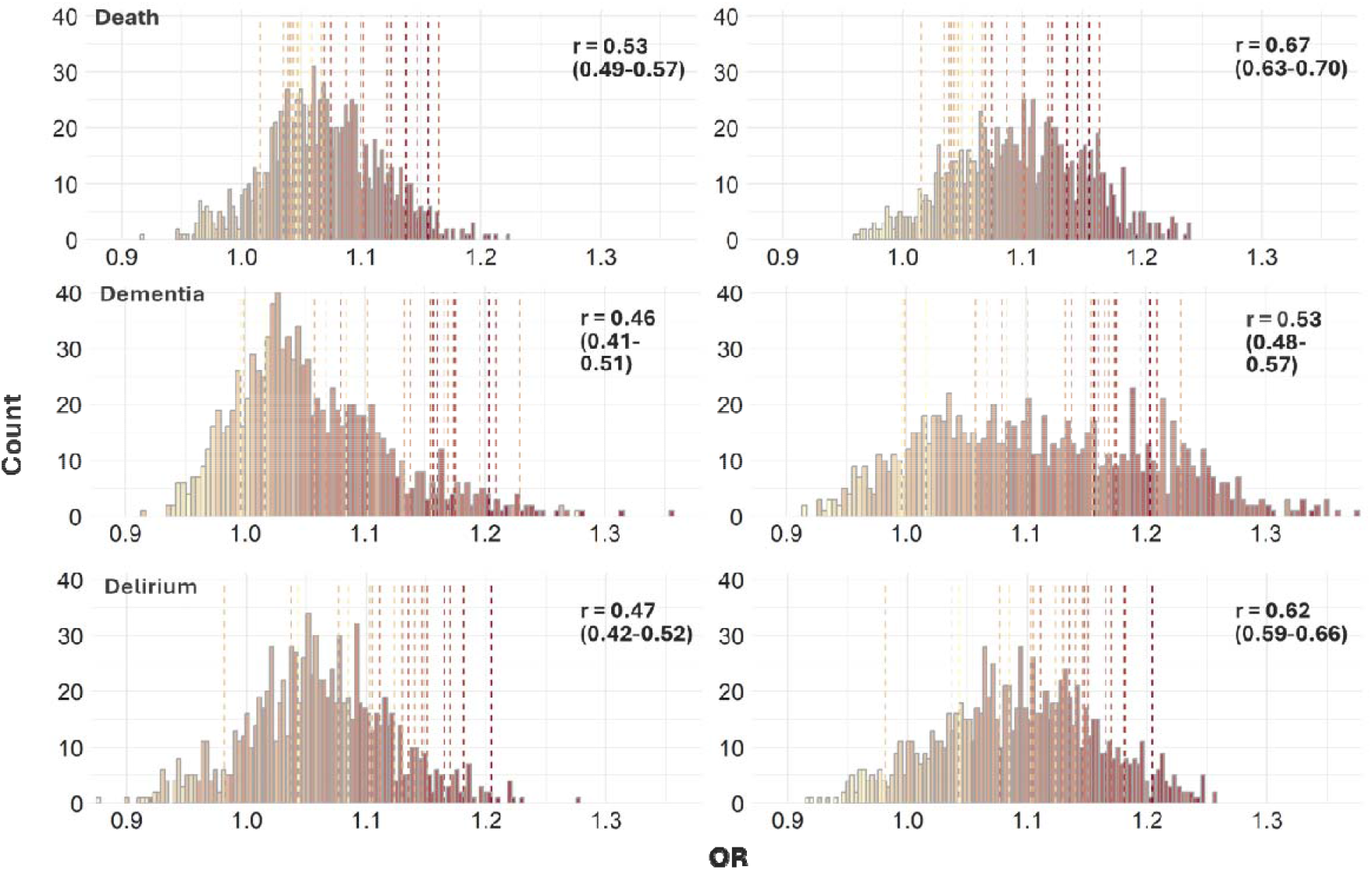
histograms and heatmaps of effect sizes for general (**left**) and anticholinergic pseudoscales (**right**) for death (**top**), dementia (**middle**), or delirium (**bottom**). The distributions are the same as presented in **Figure 1**, the colours indicate the average scale size for a given group of effect sizes. The dotted lines represent the effect sizes for existing ABS. The scale size ranges from 15 (light yellow) to 150 (dark red). The numbers in the top right corner of each graph represents Pearson’s correlation coefficient (and the 95% CI) for the correlation between the OR (indicated on the x-axis) and the size of the pseudoscales (indicated by the colour).

Because of the correlation between effect size and scale size, we constructed 250 within-sampling pseudoscales with equivalent properties to individual ABS. We then calculated for each ABS the proportion of effects of corresponding pseudoscales that were weaker than the effect of the ABS. On average, 73%, 92%, and 94% of general and 46%, 71%, and 82% of anticholinergic pseudoscales for death, dementia, and delirium, respectively, exhibited weaker effects than their corresponding ABS. The range in differences in the ORs between ABS and general polypharmacy were -0.03 to 0.10 for death, -0.02 to 0.20 for dementia, and -0.05 to 0.13 for delirium. The range in differences in the ORs between ABS and anticholinergic polypharmacy was -0.05 to 0.07, -0.07 to 0.16, and -0.09 to 0.09 for death, dementia, and delirium, respectively. ABS that exhibited the strongest effects beyond polypharmacy were DRS for death, YS, AAS, and AEC for dementia, and AAS, DDS, and AEC for delirium (**Figure 3, Suppl. Figure 3, Suppl. Tables 6, 7**).

**Figure 3:**
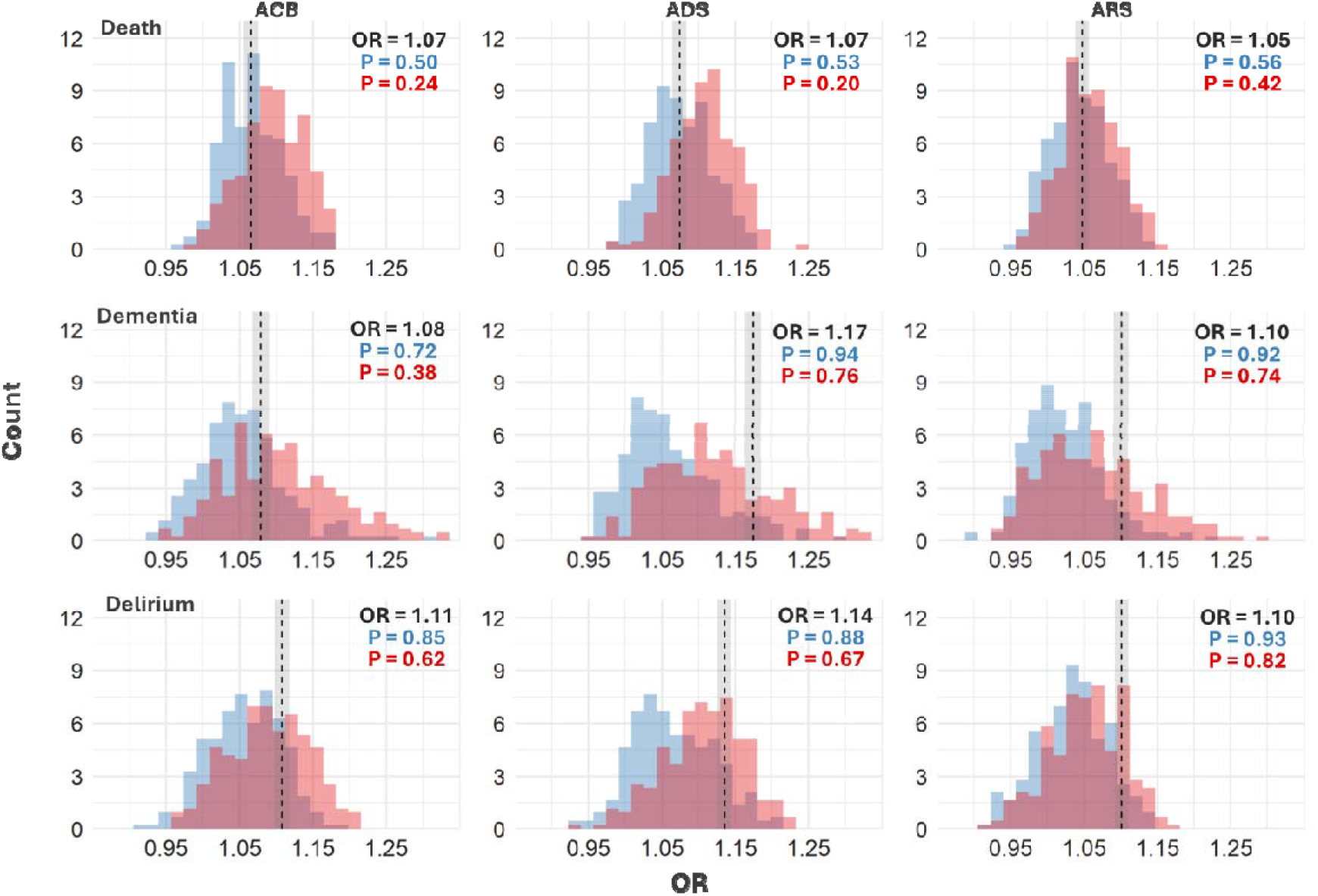
histograms of effect sizes for three exemplary ‘real-world’ ABS (dashed line) against distributions of general (**blue**) and anticholinergic (**red**) within-sampling pseudoscales when estimating the effects of drug burden on death (top row), dementia (middle row), or delirium (bottom row). Specifically, the dashed line represents the effect size for an ABS to which the distributions of effect sizes for the pseudoscales correspond (in terms of scale size and distribution of potency scores); the shaded rectangle represents the 95% CI. Depicted are the results for the Anticholinergic Cognitive Burden Scale (ACB) ^26^, the Anticholinergic Drug Scale (ADS) ^27^, and the Anticholinergic Risk Scale (ARS) ^12^ as they are commonly used in the literature. The plot displays in the top right corner the ORs for the three ABS and the cumulative probability at the ORs. The latter expresses the proportion of ORs of general or anticholinergic pseudoscales that are smaller than the OR of the ABS.

The plots for all other scales are in **Suppl. Figure 3**.

In sensitivity analyses where sampling was performed for the period from the year 2004 to 2006, the trends were broadly similar. However, there were some differences. First, compared to the year 2015, the effects for associations of all scales with the outcomes, especially with delirium, were reduced. Second, fewer pseudoscales exhibited smaller effects than their corresponding ABS in within-sampling. Third, the relative effect size for ABS compared to other ABS in within-sampling did not exactly correspond between the two time periods. For death, dementia, and delirium, the correlation between the ORs for ABS in 2015 and the ORs for ABS in 2004-2006 (n=23) was 0.83 (95% CI = 0.64-0.93), 0.66 (95% CI = 0.35-0.84), and 0.62 (95% CI = 0.28-0.82), respectively (**Suppl. Text 2, Suppl. Tables 8-10**).

## Discussion

The purpose of the present study was to assess the adverse health effects of anticholinergics independently of general polypharmacy. We showed that even after considerable adjustment for confounding, most pseudoscales - reflecting the prescribing of either general or anticholinergic drugs - were associated with an increased risk of death, dementia, and delirium. Further, the distributions of effect sizes of general and anticholinergic polypharmacy overlapped substantially. Finally, “real-world” ABS mostly exhibited larger effects than their corresponding pseudoscales.

The positive associations between the burden scores of most pseudoscales and adverse health outcomes imply a general harm of polypharmacy regardless of drug class. The high number of significant models for both general and anticholinergic polypharmacy raises questions about the utility of reporting deleterious effects of any drug score when these effects are not contrasted with valid comparators that control for the effect of polypharmacy.

The absolute difference between the cumulative prescribing of anticholinergic drugs and of general medications without putative anticholinergic effects was 0.03 to 0.05 (OR), depending on the outcome. If the effect of polypharmacy, instead of a null effect, is considered the reference value, the effect of anticholinergic use is thus much smaller than what was previously reported in studies that did not control for polypharmacy in this way^1,3,28-30^.

The finding in the present study of mostly stronger effects of ABS compared to their corresponding pseudoscales suggests that the choice of drugs to be included on ABS and the weights assigned to them provide additional prognostic value. However, the latter depended on the outcome of interest (death vs. dementia vs. delirium) and on the sampling period (year 2015 vs. years 2004-2006). This suggests that the prognostic ability of an ABS is neither completely transferable between different long-term outcomes, nor between populations with different distributions of age and drug prescription patterns.

It is also unclear whether any association between anticholinergic use and an outcome - in the present work or any other observational study - is due to true antimuscarinic action of the drugs, confounding by indication, or another effect entirely. Because construction details of ABS are often poorly reported^31^, we were unable to systematically compare ABS that were constructed using different methods. Just two ABS in our sample, AAS^32^ and YS^33^, were constructed using only serum anticholinergic activity – the only objective, if flawed^34^, procedure to measure anticholinergic potency. Moreover, although study designs in pharmacoepidemiology that enable the estimation of causal effects while minimising confounding by indication exist^35,36^, they cannot be applied on combined drug scores such as ABS that incorporate medicines prescribed for dozens of distinct indications.

### Strengths and Limitations

Our study has several strengths. First, we for the first time systematically distinguished between the effects of general polypharmacy and anticholinergic burden. Second, we adjusted for multiple potential confounders based on informed assumptions about the causal relationships between the studied variables, allowing a structured way of addressing the risk of bias. Third, the sample size was larger than in most other studies of anticholinergic burden, included a substantial number of cases, and the estimated effect sizes of commonly used ABSs were compared against a large set of effect sizes from pseudoscales constructed through different approaches.

We acknowledge several limitations of our study. First, the UK Biobank sample is not representative of the general population, which may hamper the reliable estimation of causal relationships^37-39^. Moreover, the lack of insight into the indication for prescribing prevented a thorough control for the confounding effects of underlying conditions. Second, we had no information on drug adherence and implicitly assumed complete adherence for all analyses. Third, in line with how current ABS are used, we assumed that scale scores were additive and that linear relationships existed between scale scores and the risks of the outcomes. Despite the ubiquity of such assumptions in previous work, they may not hold^40^. Fourth, we used scales to assess the effect of cumulative longitudinal use of drugs, although most ABS are not intended to be used in this way. Finally, due to the relatively young age of the participants, we could not assess anticholinergic prescribing in the oldest individuals, often identified as those most at risk of drug side effects^41^.

In conclusion, most existing ABS capture more than just polypharmacy for associations with most of the studied outcomes. The mixed results highlight the need to consider the choice of ABS for any given prognostic task, to further probe the cause of purportedly anticholinergic effects, and to focus more on individual drugs and drug groups instead of relying on drug scores that combine medicines for multiple indications.

## Data Availability

All data used in the present study are available to researchers with access to the UK Biobank dataset, which is available for a fee upon request from UK Biobank.

## Acknowledgements

JM is funded by the College of Medicine and Veterinary Medicine towards Wellcome grant 108890/B/15/Z. AKL is funded by the European Research Council under the European Union’s Horizon 2020 research and innovation program (grant number 803239). The funders were not involved in any aspect of study planning, analysis, or interpretation. The authors thank the participants of UK Biobank for providing their data for the study and Dr Elizabeth Rose Mayeda for helpful suggestions for the simulation approach. The study was performed using data from UK Biobank application 10279.

