## Supplementary material for "Is there an anticholinergic effect of drugs beyond polypharmacy? A simulation study on death, dementia, and delirium in UK Biobank": supplement_plain.docx

**A simulation study comparing anticholinergic drug use against polypharmacy on death, dementia, and delirium in UK Biobank**

**Supplementary material**

**Mur, J.^[1]^, Stirland, L. E.****^[2]^, Muniz-Terrera, G.^2,^^[3]^, & Leist, A. K.^[4]^**

**Suppl. Table 1**: some properties of the ABS used in the analysis. Two scales were updated since the initial publication (Aging Brain Care, 2012; Carnahan, 2014, personal communication on 21.10.2019) ^1,2^. The table also displays the scale size (i.e., the number of drugs with a score of >0 that were included in the scale and present in the sample in the year 2015), the proportion of participants with a burden score above zero, and the median burden score among participants with burden scores above zero. The last two rows display these statistics for pseudoscales. For general and anticholinergic pseudoscales, the median across the two pseudoscales groups, respectively, is shown for both the proportions and the median burden score values. The ABS list was updated from a list published previously^3^.

| **First author** | **Scale name** | **Year of publication** | **Size** | **Prop. >0** | **Median** |
| --- | --- | --- | --- | --- | --- |
| Summers^4^ | Drug Risk Number (DRN) | 1978 | 41 | 0.25 | 12 |
| Han^5^ | Clinician-rated Anticholinergic Scale (CrAS) | 2001 | 51 | 0.27 | 7 |
| Ancelin^6^ | Anticholinergic Burden Classification scale (ABC) | 2006 | 17 | 0.10 | 14 |
| Carnahan^2^ | Anticholinergic Drug Scale (ADS) | 2006 (2014) | 96 | 0.28 | 7 |
| Chew^7^ | Anticholinergic Activity Scale (AAS) | 2008 | 35 | 0.37 | 4 |
| Cancelli^8^ | Cancelli’s Anticholinergic Burden Scale (CABS) | 2008 | 13 | 0.12 | 18 |
| Rudolph^9^ | Anticholinergic Risk Scale (ARS) | 2008 | 40 | 0.19 | 7 |
| Ehrt^10^ | Revised Anticholinergic Activity Scale (AAS-r) | 2010 | 23 | 0.25 | 9 |
| Sittironnarit^11^ | Anticholinergic Loading Scale (ALS) | 2011 | 44 | 0.32 | 7 |
| Boustani^1^ | Anticholinergic Cognitive Burden (ACB) | 2008 (2012) | 80 | 0.29 | 8 |
| Sumukadas^12^ | Modified Anticholinergic Risk Scale (m-ARS) | 2013 | 57 | 0.20 | 8 |
| Durán^13^ | Durán Scale (DS) | 2013 | 74 | 0.28 | 6 |
| Hefner^14^ | Delirogenic Risk Scale (DRS) | 2015 | 88 | 0.43 | 12 |
| Nguyen^15^ | Drug Delirium Scale (DDS) | 2016 | 81 | 0.27 | 6 |
| Bishara^16^ | Anticholinergic effect on cognition scale (AEC) | 2017 | 56 | 0.24 | 7 |
| Briet^17^ | Anticholinergic impregnation scale (AIS) | 2017 | 109 | 0.38 | 8 |
| Kiesel^18^ | German Anticholinergic Burden Scale (GABS) | 2018 | 134 | 0.45 | 9 |
| Nery^19^ | Brazilian anticholinergic activity drug scale (BAAS) | 2019 | 100 | 0.37 | 8 |
| Jun^20^ | Korean Anticholinergic Burden Scale (KABS) | 2019 | 87 | 0.32 | 7 |
| Kable^21^ | Modified Anticholinergic Burden Scale (mACB) | 2019 | 72 | 0.33 | 8 |
| Ramos^22^ | CRIDECO Anticholinergic Load Scale (CALS) | 2020 | 150 | 0.43 | 9 |
| Al Rihani^23^ | AntiCholinergic and Sedative Burden Catalog (ACSBC) | 2021 | 133 | 0.44 | 9 |
| *Yamada^24^ | Yamada’s scale (YS) | 2023 | 55 | 0.27 | 9 |
|  | General pseudoscales |  | 83 | 0.39 | 9 |
|  | Anticholinergic pseudoscales |  | 83 | 0.35 | 9.5 |

**Note: Fluticasone fluorate and fluticasone propionate are scored differently according to this scale. Due to difficulties in distinguishing the two drugs in the sample, both were classified as having the same potency of 1 on this scale.*

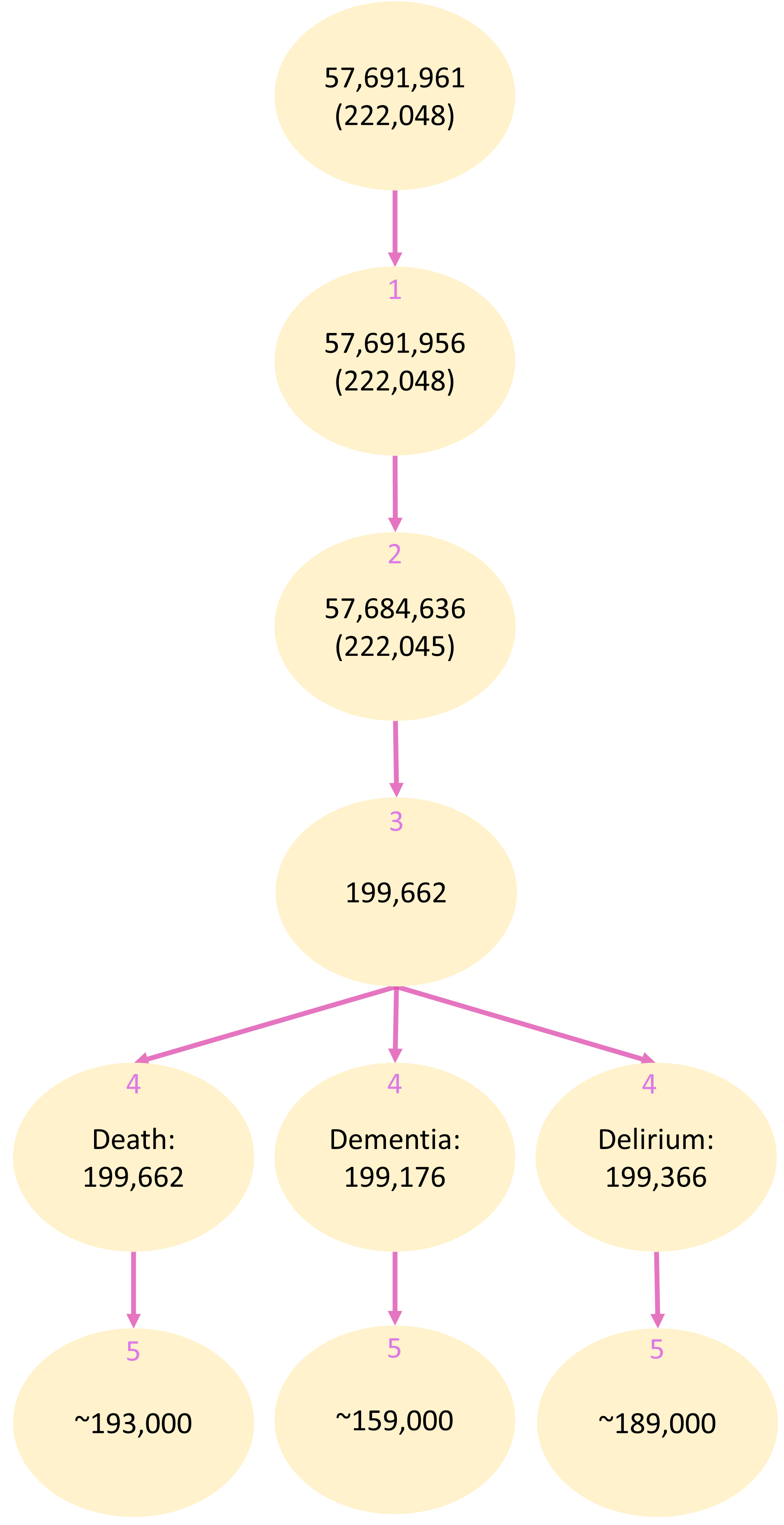

**Suppl. Figure 1**: changes in sample size during the data cleaning procedure. In each ellipse, the upper row represents the number of individual observations, the second row represents the number of participants. Where only a single row is present, participants are individual observations. The data cleaning steps are (1) removal of prescriptions missing their contents, (2) removal of invalid dates, (3) transformation from a prescription-based dataset into an id-year dataset and removal of years outside the year 2015, (4) removal of observations of the outcome that occurred before or during the year 2015, and (5) removal of observations with incomplete data for any of the covariates used in the model. The final number of observations for the modelling (after step 5) is the average across scales, because the number of removedoutlier observations for the main predictor (burden according to the relevant scale) differed between the scales.

**Suppl. Figure 2:** simplified directed acyclic graphs (DAGs) for the assumed causal relationships between drug burden (X) and an idealised health outcome (Y). For simplicity and illustration purposes, the DAGs do not depict any other common causes of Y and X other than underlying health conditions (U) and medications (B0 and B1; see below). The anticholinergic burden (X) is fully determined by the drugs that compose the burden list (anticholinergic polypharmacy, B1) and the potency scores that are assigned to those drugs (P). Non-anticholinergic polypharmacy (B0) may also cause the outcome. To determine the causal effect of X on Y, B0 can be statistically adjusted for, but B1 cannot be due to its high correlation with X. The **left** DAG assumes no control for confounding, the **right** DAG includes control for non-anticholinergic polypharmacy (B0) and underlying disorders (U). Note that the confounding pathway between X and Y persists even in the case of complete control for B0 and U. In our analysis, this residual confounding is adjusted through the simulation of pseudo-scales by sampling from anticholinergic polypharmacy (B1) and comparing the observed effects with those of existing scales (X). The DAGs were constructed using *dagitty* (<https://dagitty.net>)^25^.

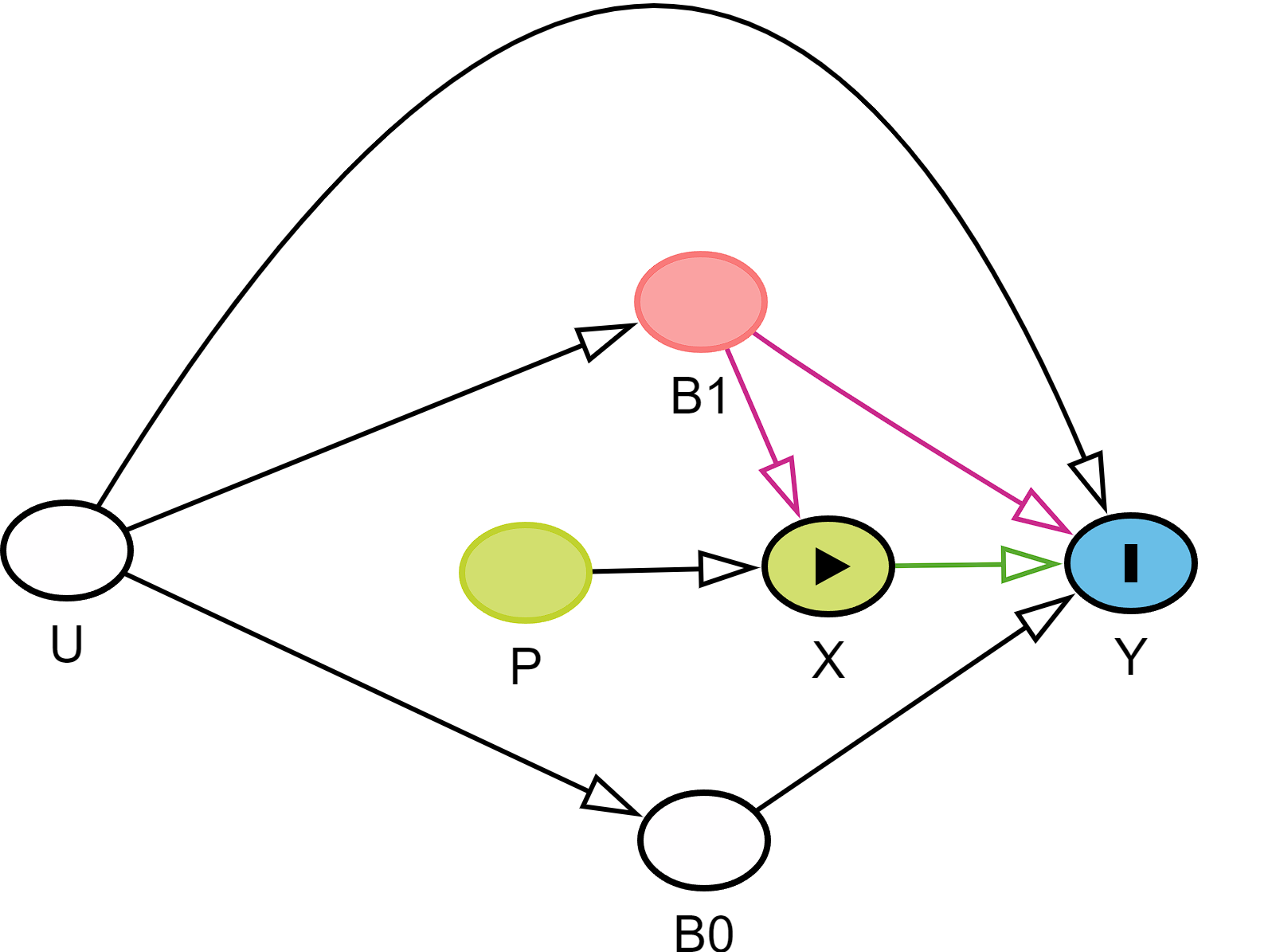

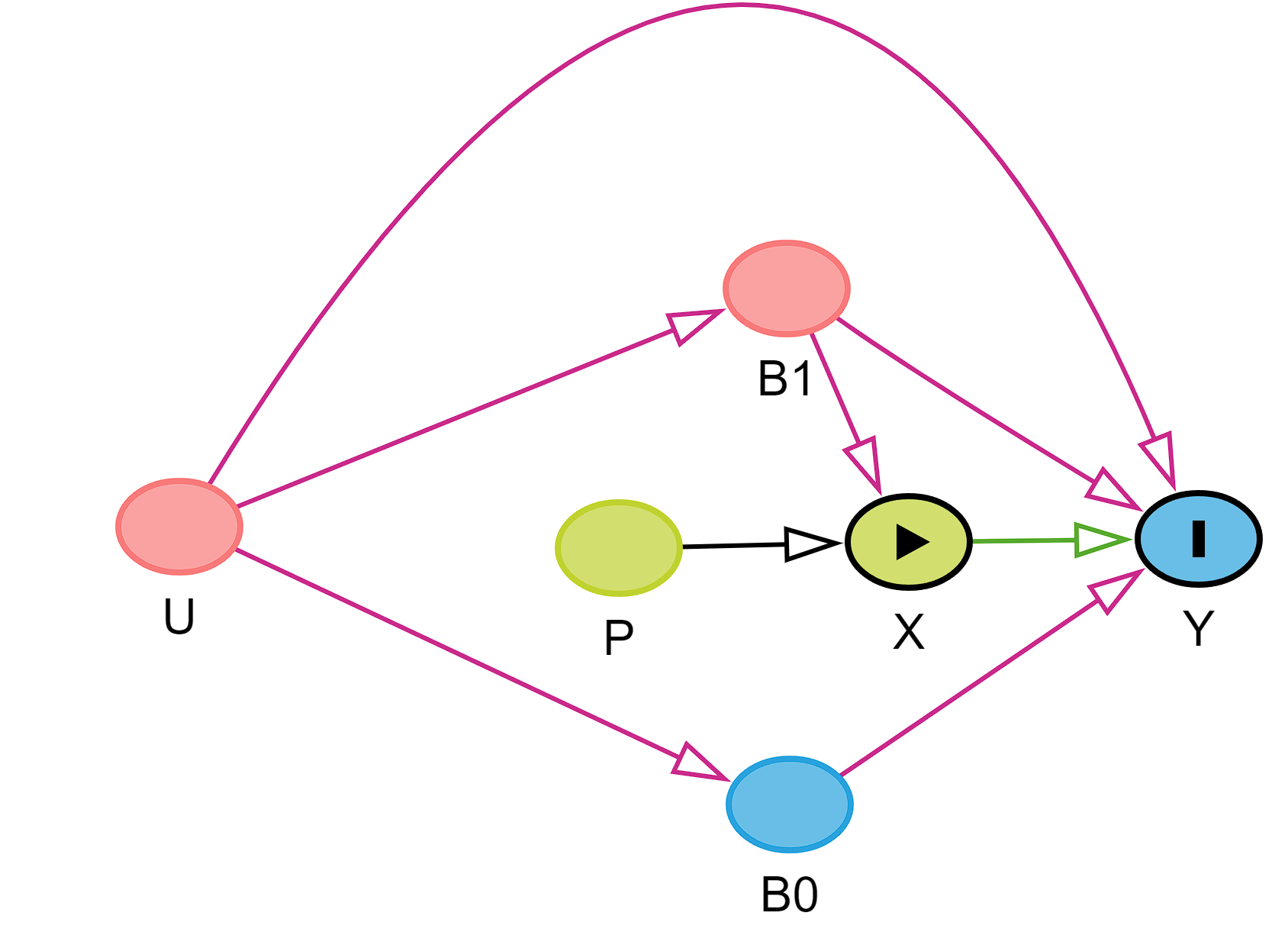

Legend: X: exposure (drug burden), Y: outcome, B1: (anticholinergic polypharmacy), B0: (non-anticholinergic polypharmacy), P: potency scoring, U: underlying disorders. Causal paths are represented with arrows; red arrows signify biasing paths, the green arrow is the causal paths from the exposure to the outcome, white arrows are paths that do not confound the estimation of the causal path between the exposure and the outcome. Green, blue, and red ellipses signify ancestors of exposure, ancestors of outcome, and ancestor of exposure and outcome, respectively.

**Suppl. Table 2**: variables used in our analyses. The individual columns give the names of the variables, the field IDs in the UK Biobank showcase (<https://biobank.ndph.ox.ac.uk/ukb/>) used to derive the variables, the role of the variables in the models, and brief descriptions.

| **Name** | **Field ID** | **Role** | **Description** |
| --- | --- | --- | --- |
| *Drug burden | 42039 | predictor (all) | *drug_name* column in record table 1062 |
| Death | 40000 | outcome |  |
| Dementia | 42018 | outcome, covariate (death, delirium) |  |
| Delirium | 130846 | outcome |  |
| *Data provider | 42039 | covariate (all) | *data_provider* column in record table 1062 |
| Age | 34, 52 | covariate (all) | date of birth approximated from month and year of birth. |
| Sex | 31 | covariate (all) | binary variable |
| Non-scale polypharmacy | 42039 | covariate (all) | *drug_name* column in record table 1062 |
| *Education | 6138 | covariate (all) |  |
| *Deprivation | 189 | covariate (all) |  |
| *General cognitive ability | 20016, 20018, 20023, 399, 4282 | covariate (dementia, delirium) |  |
| *Air pollution | 24004, 24006, 24016, 24017, 24018, 24003 | covariate (dementia) |  |
| *Alcohol use frequency | 1558 | covariate (all) |  |
| Waist circumference | 48 | covariate (all) |  |
| Smoking status | 20116 | covariate (all) | three categories: smokers, non-smokers, previous smokers |
| *Physical activity | 6164 | covariate (all) |  |
| Mood disorder | 130890, 130892, 130894, 130896, 130898, 130900, 130902 | covariate (dementia, delirium) |  |
| Diabetes | 130706, 130708, 130710, 130712, 130714 | covariate (dementia, death) |  |
| Hypertension | 131286 | covariate (dementia) |  |
| Hyperlipidaemia | 130814 | covariate (dementia) |  |
| Psychotic disorder | 130874, 130876, 130878, 130880, 130882, 130884, 130886, 130888 | covariate (delirium) |  |
| Visual impairment | 131212 | covariate (delirium) |  |
| *Hearing impairment | 2247, 2257, 131258, 131260, 20019, 20021, 41270, 41280, 41271, 41281, category ID 2006, 42040 | covariate (dementia, delirium) |  |
| Sleep disorder | 131060, 130920 | covariate (delirium) |  |
| Vascular CNS disorder | 42006, 131370, 131372, 131374, 131376, 131378 | covariate (dementia, delirium) |  |
| Inflammatory CNS disorder | 131000, 130992, 130994, 130996, 130998, 131002, 131004, 131006, 131008, 131010 | covariate (dementia, delirium) |  |
| Atrophic CNS disorder | 131012, 131014, 131016, 131018, 131020, 42028 | covariate (dementia, delirium) |  |
| Movement CNS disorder | 131022, 131024, 131026, 131028, 131030, 131032 | covariate (dementia, delirium) |  |
| Demyelinating CNS disorder | 131042, 131044, 131046 | covariate (dementia, delirium) |  |
| Paroxysmal CNS disorder | 131048, 131050, 131056, 131058 | covariate (dementia, delirium) |  |
| Brain tumour | 40005, 40006, 40013 | covariate (dementia, delirium) | for ICD-codes, see **Suppl. Table 3** |
| *Traumatic brain injury (TBI) | 41270, 41280, 41271, 41281, category ID 2006, 42040 | covariate (dementia, delirium) |  |
| Other CNS disorder | 131038, 131040, 131100, 131110, 131112, 131114, 131116, 131120 |  |  |
| Endocrinopathy | 130690, 130692, 130694, 130696, 130698, 130700, 130702, 130704, 130718, 130720, 130722, 130724, 130726, 130728, 130730, 130732, 130734, 130736, 130738, 130742, 130744, 130746, 130748, 40005, 40006, 40013 | covariate (delirium) | includes endocrine cancer in **Suppl. Table 3** |
| Nutritional deficiency | 130750, 130752, 130756, 130758, 130760, 130762, 130764, 130766, 130768, 130770, 130772, 130774, 130776, 130778, 130780, 130782, 130784, 130786, 130788 | covariate (delirium) |  |
| Metabolic disorder | 130798, 130800, 130802, 130806, 130808, 130810, 130812, 130814, 130816, 130818, 130820, 130822, 130824, 130826, 130828, 130830, 130832, 130834 | covariate (delirium) |  |
| Cerebrovascular disease | 131360, 131362, 131364, 131366, 131368, 131370, 131372, 131374, 131376, 131378 | covariate (all) |  |
| Chronic lower respiratory disease | 131484, 131486, 131488, 131490, 131492, 131494, 131496, 131498 | covariate (death) |  |
| Liver disease | 131658, 131660, 131662, 131664, 131666, 131668, 131670 | covariate (death) |  |
| Influenza or pneumonia | 131438, 131440, 131442, 131444, 131446, 131448, 131450, 131452, 131454, 131456 | covariate (death) |  |
| Ischaemic heart disease | 131296, 131298, 131300, 131302, 131304, 131306 | covariate (dementia, death) |  |
| Colon cancer | 40005, 40006, 40013 | covariate (death) | for ICD-codes, see **Suppl. Table 3** |
| Prostate cancer | 40005, 40006, 40013 | covariate (death) | for ICD-codes, see **Suppl. Table 3** |
| Lung cancer | 40005, 40006, 40013 | covariate (death) | for ICD-codes, see **Suppl. Table 3** |
| Breast cancer | 40005, 40006, 40013 | covariate (death) | for ICD-codes, see **Suppl. Table 3** |
| Ovarian cancer | 40005, 40006, 40013 | covariate (death) | for ICD-codes, see **Suppl. Table 3** |
| *Social isolation | 709, 1031, 6160 | covariate (dementia, delirium) |  |
| Loneliness | 2020 | covariate (dementia, delirium) | binary variable |
| *Depressive mood | 2050 | covariate (dementia) |  |

**Note: variables that were either recoded or otherwise transformed and for which the explanation is not provided in the main text, are described in further detail in* ***Suppl. Text 1****.*

**Suppl. Table 3**: ICD-codes for diagnoses of cancer which were unavailable as derived variables (first occurrences or algorithmically-defined outcomes) in UK Biobank. We instead used the UK Biobank cancer register (field IDs 40005, 40006, 40013) which is linked to UK national cancer registries.

| **Disorder** | **ICD10 codes** | **ICD9 codes** |
| --- | --- | --- |
| Colon cancer | C18, C180-189, C19-21, C210-212, C218 | 153, 1530-1539, 154, 1540-1543, 1548, 2303, 2304 |
| Prostate cancer | C61 | 185, 2365 |
| Lung cancer | C33, C34, C340-343, C348, C349 | 162, 1620, 1622, 1623, 1624, 1625, 1628, 1629, 2357 |
| Breast cancer | C50, C500-506, C508, C509 | 174, 1740-1746, 1748, 1749, 2330 |
| Ovarian cancer | C56 | 83, 1830, 1832, 184, 1840, 1841, 1844, 1848, 1849, 1986, 2362 |
| Brain tumour | D32, D320, D321, D329, D33, D331-334, D337, C70, C700, C709, C71, C710-719, C793 | 225, 2258, 2259, 2250-2254, 1921, 191, 1910, 1911, 1912, 1916-1919 |
| Endocrine cancer | C25, C250-254, C257-259, C73, C74, C740, C741, C749, C75, C750-755, D093, D34, D35, D351, D352, D44, D440-440 | 157, 1570, 1572, 1574, 1579, 193, 194, 1940, 1941, 1943, 1944, 1949, 226, 227, 2273, 2279, 2370 |

**Suppl. Text 1**: description of selected variables used in the analyses. When we write *inpatient diagnoses*, we are specifically referring to category ID 2002 (field IDs 41270, 41280, 41271, 41281) where diagnostic codes were mapped to dates in the summary diagnoses tables. When we write *primary care*, we are specifically referring to field ID 42040 that contains GP clinical event records. To query the latter, we used *readv2* and *readv3* codes provided by the NHS Digital Technology Reference Data Update Distribution (<https://isd.digital.nhs.uk/trud/user/guest/group/0/pack/9>).

- **Drug burden:** prescriptions were available from ~1990 until May 2017 for Scotland (EMIS/Vision), September 2017 for Wales, June 2017 for England (Vision) and August 2016 for England (TPP) ^26^.
- **Data provider**: each prescription is associated with a data provider (England: TPP, Vision; Scotland: EMIS/Vision, Wales: EMIS/Vision). When the data were transformed into the id-year format and restricted to the year 2015, the most frequent data provider for that period was chosen for each participant. For participants for which periods of continuous electronic health record ascertainment were inferred^27^, but no prescriptions were prescribed in the year 2015, the data provider was imputed by choosing that data provider which appeared closest in time to the year 2015. This imputed the data provider for 30,638 participants. Because some participants for whom the period of continuous health record ascertainment were inferred did not receive prescriptions (n = 1,941), those participants were classed as a separate group. Due to a lack of pollution data for participants in Scotland, the analysis for dementia was performed only on participants from England and Wales. In the sensitivity analysis where prescriptions in the years 2004-2006 were analysed, data providers were imputed in an analogous way, but participants from the Scottish data providers were excluded from the analyses for all three outcomes (as opposed to just dementia as in the main analysis). This was done due to a system-wide block of prescription records for Scotland before the year 2012^26^ and a consequent failure to assign many prescriptions to Scottish participants in that period. This would have led to a much lower prescription count for Scottish participants if the latter were retained in the analyses.
- **Education**: self-report on the ascertained qualifications was recorded in six categories: *college or university degree*, *A levels/AS levels or equivalent*, *O levels/GCSEs or equivalent*, *CSEs or equivalent*, *NVQ or HND or HNC or equivalent*, and *other professional qualifications eg: nursing, teaching*. We recoded the classification into a binary variable, thus distinguishing between participants with and without a college degree.
- **Socioeconomic deprivation**: the Townsend deprivation index^28^ was provided by UK Biobank. The index is calculated based on the national census that preceded the participants’ joining UK Biobank. It scores several socioeconomic indicators and sums them up. The scores are then standardized, and each participant is assigned a score corresponding to their geographical area.
- **General cognitive ability**: a factor of general intelligence was calculated as before^3,29^. In short, we fitted a confirmatory factor analysis in a structural equation modelling framework using tests of visual declarative memory, processing speed, and – for a subsample of participants – tests of working memory, prospective memory, and verbal and numerical reasoning.
- **Air pollution**: a principal component analysis (PCA) was run on pollution due to nitrogen oxides (in the year 2010), nitrogen dioxide (mean for the years 2005, 2006, 2007, 2010), and particulate matter (pm25; in the year 2010). The pollutants were chosen due to having been previously associated with dementia^30^. We used the first principal component.
- **Alcohol use**: alcohol intake frequency was recorded in six categories: *daily or almost daily*, *three or four times a week*, *once or twice a week*, *one to three times a month*, *special occasions only*, and *never*.
- **Physical activity**: the types of physical activity undertaken in the four weeks before the assessment; recorded in six categories: *walking for pleasure (not as a means of transport)*, *other exercises (eg: swimming, cycling, keep fit, bowling)*, *strenuous sports*, *light DIY (eg: pruning, watering the lawn)*, *heavy DIY (eg: weeding, lawn mowing, carpentry, digging)*, and *none of the above*. We recoded the variable into three categories as previously: low (light DIY or none), medium (walking for pleasure, other exercises, or heavy DIY), and high (strenuous sports) ^31,32^.
- **Hearing impairment**: hearing impairment was identified based on self-report, objective hearing test during the assessment, and medical records. For self-report, hearing impairment was determined if the participant indicated to have both difficulties with their hearing and difficulties with their hearing in the presence of background noise. For the objective hearing assessment, hearing impairment was determined if the speech-reception-threshold (SRT) value for the better ear was above -5.5; this threshold was chosen based on previous studies^33,34^. For medical health records, hearing impairment was determined using the first occurrences variables in UK Biobank (H90: conductive and sensorineural hearing loss, H91: other hearing loss) and a custom search of the inpatient and primary-care medical record. The codes used to identify hearing loss in the EHR and a more detailed procedure to derive the variable from all three sources were performed previously^35^.
- **TBI**: TBI was determined through a custom search of the inpatient and primary care medical records. The codes used are available in **Suppl. Table 4**.
- **Social isolation**: based on self-reported number of people in the household, frequency of family/friend visits, and selection of social leisure activities. Each of the three answers was scored for social isolation and the score thus ranged from zero to three. Participants with scores of zero or one were classified as not socially isolated as previously^36^. We took further steps to avoid misclassification in cases where at least one of the three questions was not answered by the participants. Participants with a score of zero were classified as not socially isolated only if at most one of the answers was missing. Participants with a score of one were classified as not socially isolated only if no answers were missing. If two or more answers were missing, the score was set to NA.
- **Depressed mood**: self-reported frequency of depressed mood in the week before the assessment; recorded as four categories: *not at all*, *several days*, *more than half the days*, and *nearly every day*.

**Suppl. Table 5**: descriptive statistics (number and percentages, or median and interquartile range) for covariates specific to the individual outcomes. See main text for descriptive statistics of covariates used in all models.

|  | **N (%)** | | | | | |
| --- | --- | --- | --- | --- | --- | --- |
|  | **Death** | | **Dementia** | | **Delirium** | |
|  | **No** | **Yes** | **No** | **Yes** | **No** | **Yes** |
| Lower respiratory disease | 31,800 (17.2) | 2,509 (27.0) |  |  |  |  |
| Diabetes | 12,211 (6.6) | 1,635 (17.6) | 10,730 (6.7) | 350 (18.9) |  |  |
| Liver disease | 5,014 (2.7) | 594 (6.4) |  |  |  |  |
| Influenza/pneumonia | 15,655 (8.4) | 1,477 (15.9) |  |  |  |  |
| Ischaemic heart disease | 14,570 (7.9) | 2,012 (21.6) |  |  |  |  |
| Colon cancer | 1,721 (0.9) | 309 (3.3) |  |  |  |  |
| Ovarian or prostate cancer | 3,374 (1.8) | 541 (5.8) |  |  |  |  |
| Lung cancer | 225 (0.1) | 215 (2.3) |  |  |  |  |
| Breast cancer | 5,258 (2.8) | 517 (5.6) |  |  |  |  |
| General cognitive ability, median (IQR) |  |  | 0.05 (0.99) | -0.26 (0.99) | 0.04 (0.96) | -0.23 (0.99) |
| *Air pollution, median (IQR) |  |  |  |  |  |  |
| *nitrogen dioxide (μg/m^3^)* |  |  | 27.2 (9.9) | 27.7 (9.7) |  |  |
| *nitrogen oxides (μg/m^3^)* |  |  | 41.5 (15.7) | 41.8 (15.0) |  |  |
| *pm25 (μg/m^3^)* |  |  | 9.9 (1.3) | 9.9 (1.2) |  |  |
| Hypertension |  |  | 51,547 (32.5) | 1,030 (55.7) |  |  |
| Hypercholesterolemia |  |  | 33,619 (21.2) | 760 (41.1) |  |  |
| Depressed mood |  |  |  |  |  |  |
| *not at all* |  |  | 121983 (76.9) | 1,426 (77.1) |  |  |
| *several days* |  |  | 28,758 (18.1) | 314 (17.0) |  |  |
| *more than half the days* |  |  | 4,807 (3.0) | 64 (3.5) |  |  |
| *nearly every day* |  |  | 3,034 (1.9) | 46 (2.5) |  |  |
| Social isolation |  |  | 73,182 (46.1) | 944 (51.0) | 87,684 (46.4) | 1,182 (54.5) |
| Loneliness |  |  | 28,027 (17.7) | 336 (18.2) | 34,438 (18.2) | 479 (22.1) |
| Mood disorder |  |  | 21,300 (13.4) | 350 (18.9) | 25,416 (13.4) | 491 (22.6) |
| CNS disorder |  |  |  |  |  |  |
| *inflammatory* |  |  | 1,169 (0.7) | 14 (0.8) |  |  |
| *atrophic* |  |  | 216 (0.1) | 9 (0.5) |  |  |
| *movement* |  |  | 3,580 (2.3) | 180 (9.7) |  |  |
| *demyelinating* |  |  | 815 (0.5) | 16 (0.9) |  |  |
| *paroxysmal* |  |  | 5,219 (3.3) | 179 (9.7) |  |  |
| *CNS cancer* |  |  | 305 (0.2) | 10 (0.5) |  |  |
| *TBI* |  |  | 4,988 (3.1) | 104 (5.6) |  |  |
| *other* |  |  | 8,731 (5.5) | 140 (7.6) |  |  |
| Hearing impairment |  |  | 47,714 (30.1) | 754 (40.8) | 55,092 (29.1) | 886 (40.9) |
| Psychotic disorder |  |  |  |  | 688 (0.4) | 44 (2.0) |
| Visual impairment |  |  |  |  | 1,313 (0.7) | 46 (2.2) |
| Sleep disorder |  |  |  |  | 9,526 (5.0) | 188 (8.7) |
| Endocrinopathy |  |  |  |  | 20,780 (11.0) | 375 (17.3) |
| Nutritional deficiency |  |  |  |  | 2,559 (1.4) | 80 (3.7) |
| Metabolic disorder |  |  |  |  | 44,199 (23.4) | 1,064 (49.1) |

**Note: for pollution, the statistics of the components that were used to calculate the principal component are shown*.

**Suppl. Figure 3**: histograms of effect sizes for general (**blue**) and anticholinergic (**red**) within-sampling pseudoscales when estimating the effects of drug burden on death, dementia, or delirium. The dashed line represents the effect size for ABS to which the distributions of effect sizes for the pseudoscales correspond (in terms of scale size and distribution of potency scores); the shaded rectangle represents the 95% CI.

**Death**

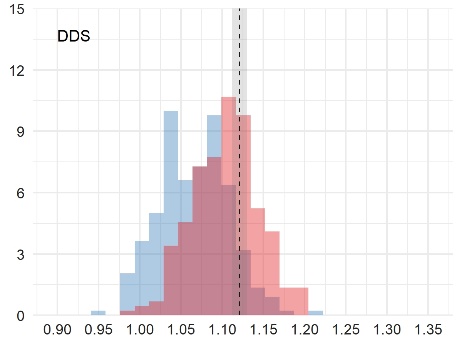

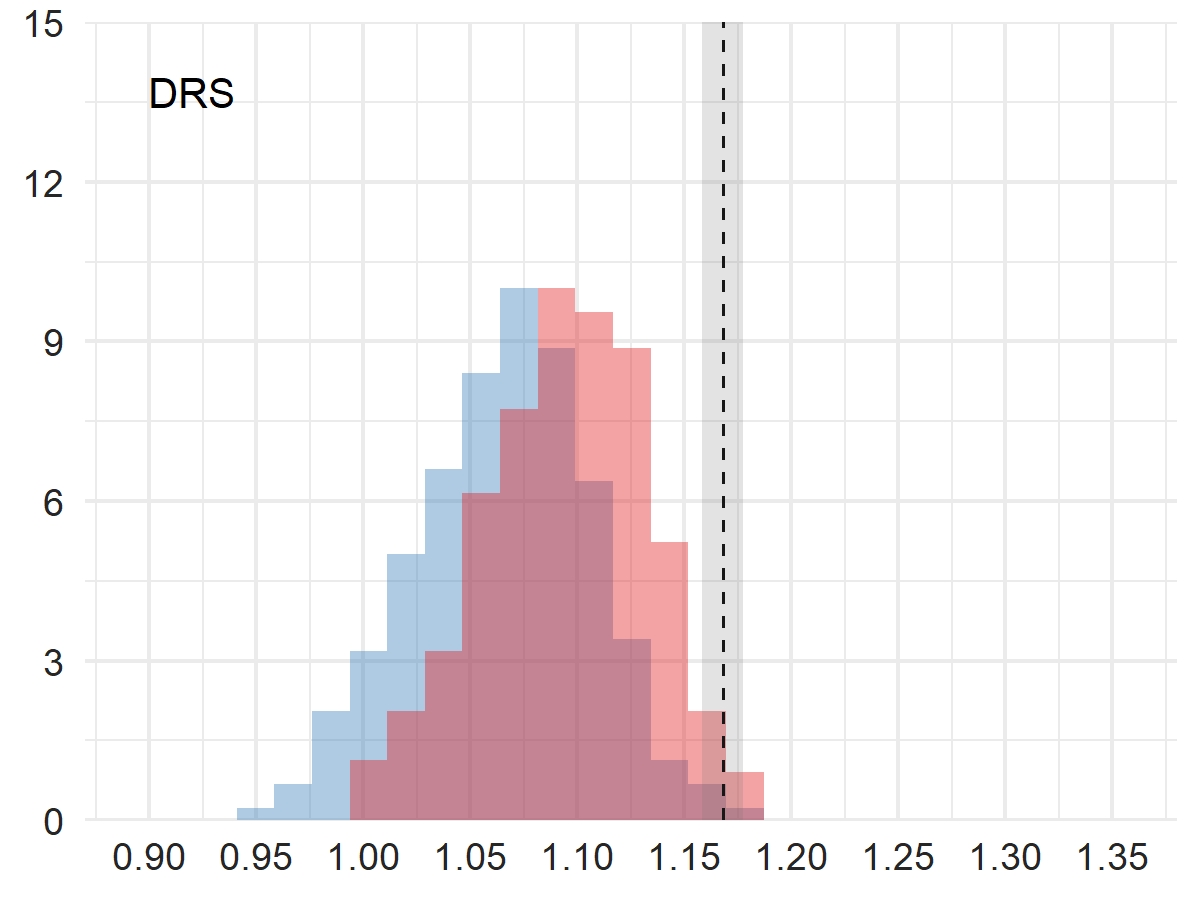

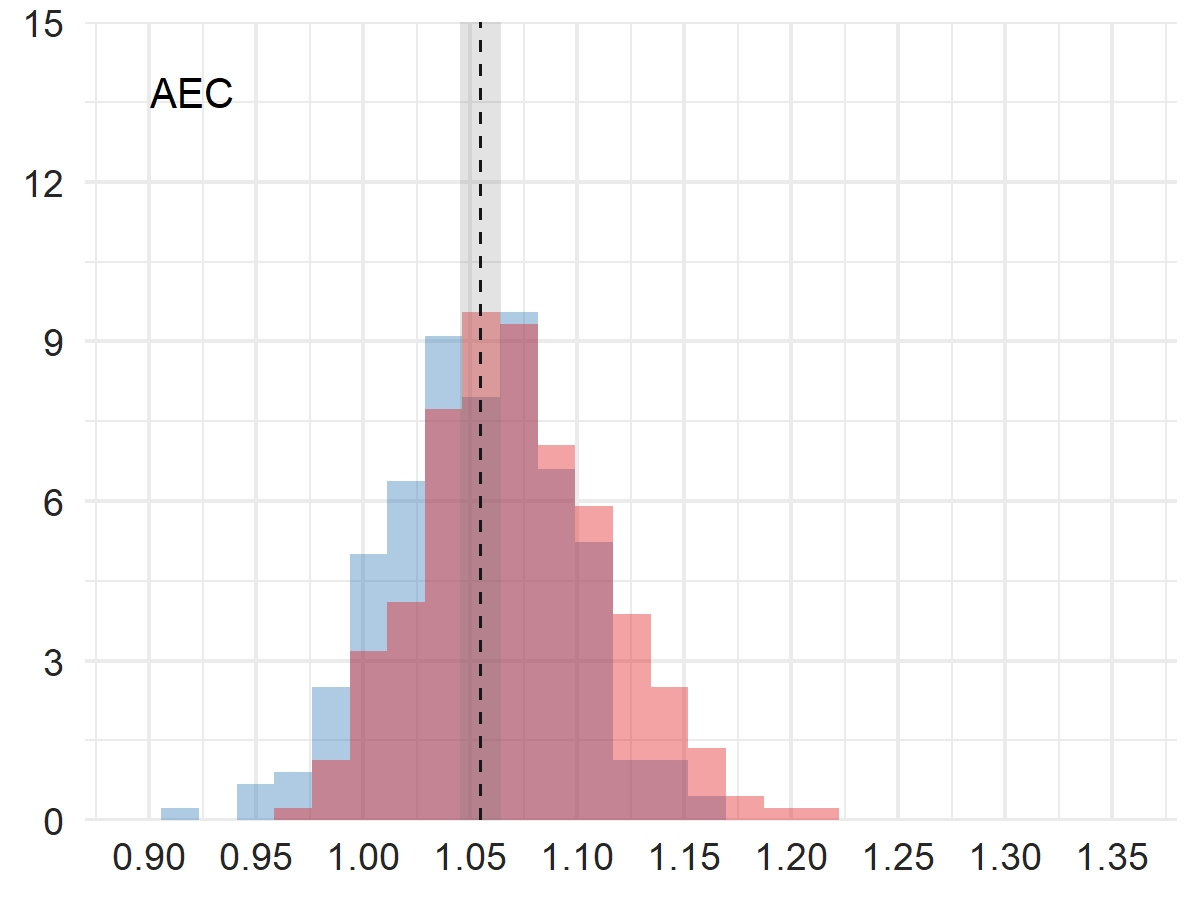

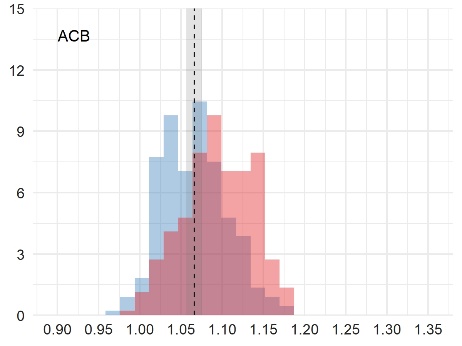

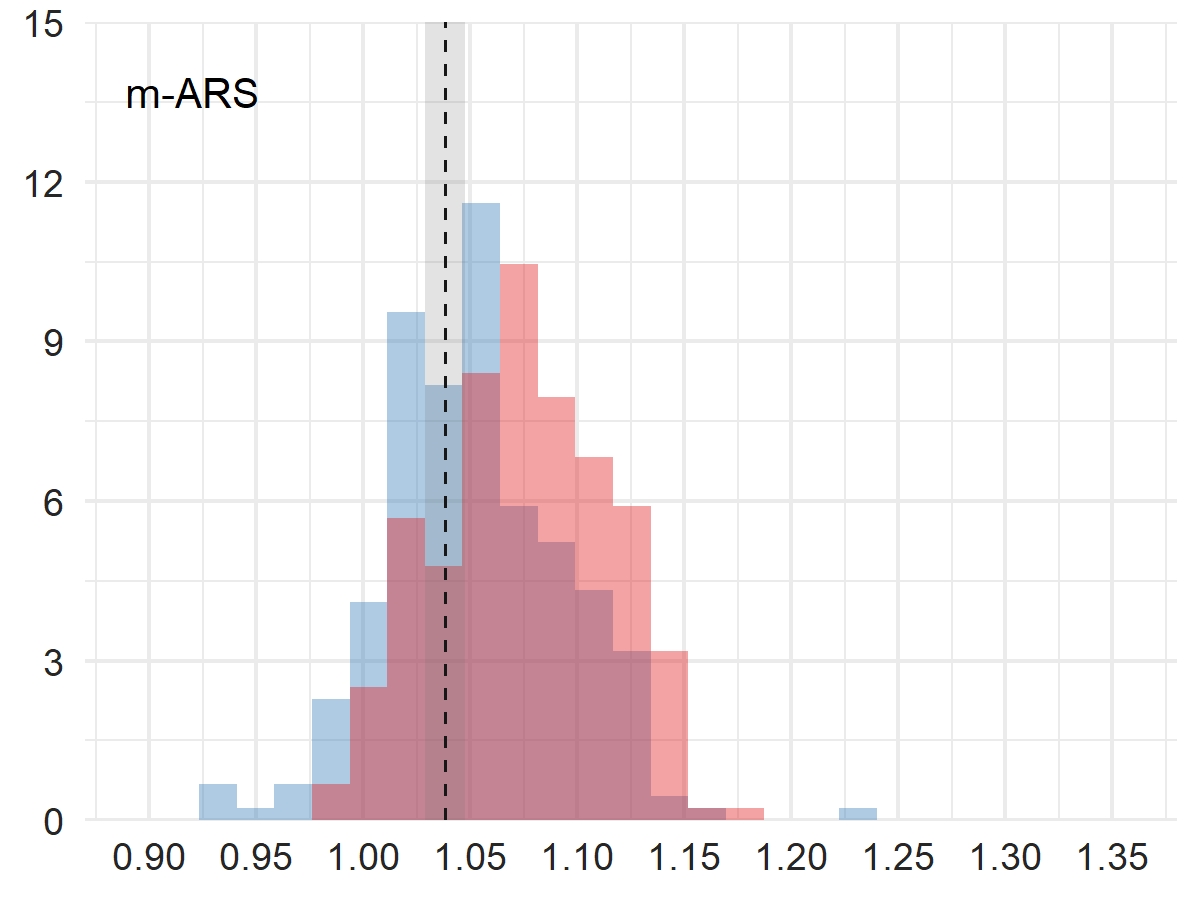

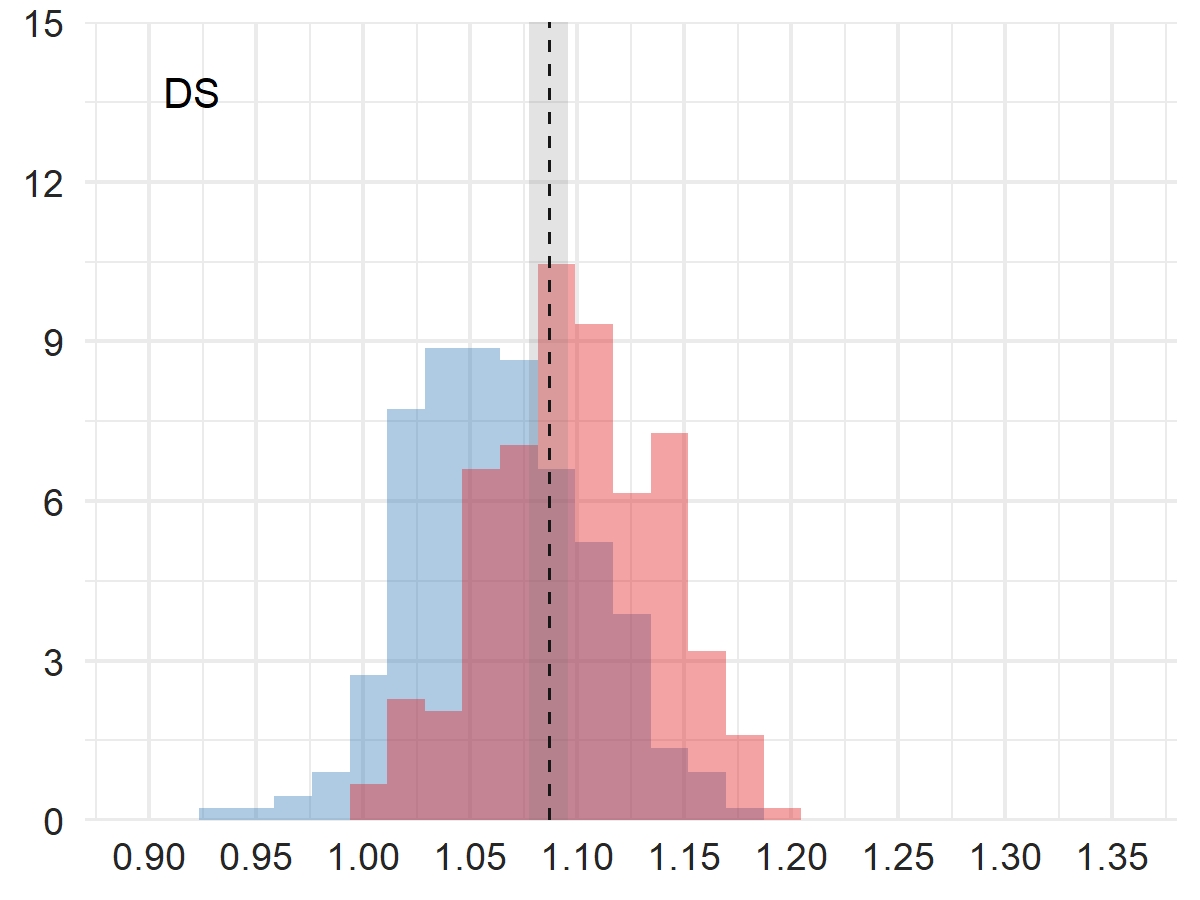

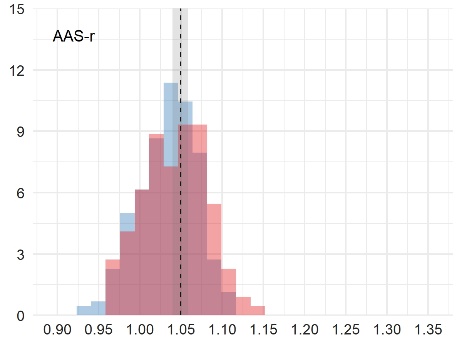

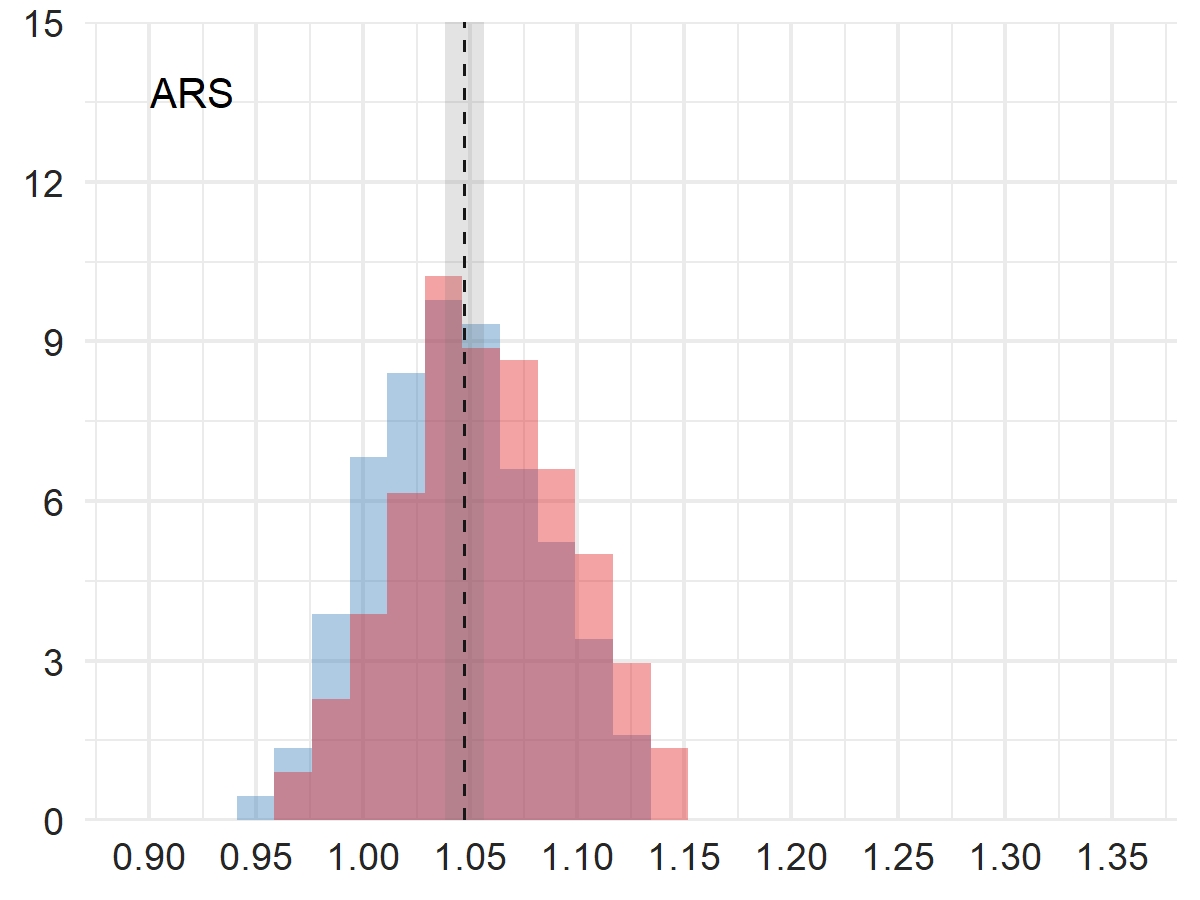

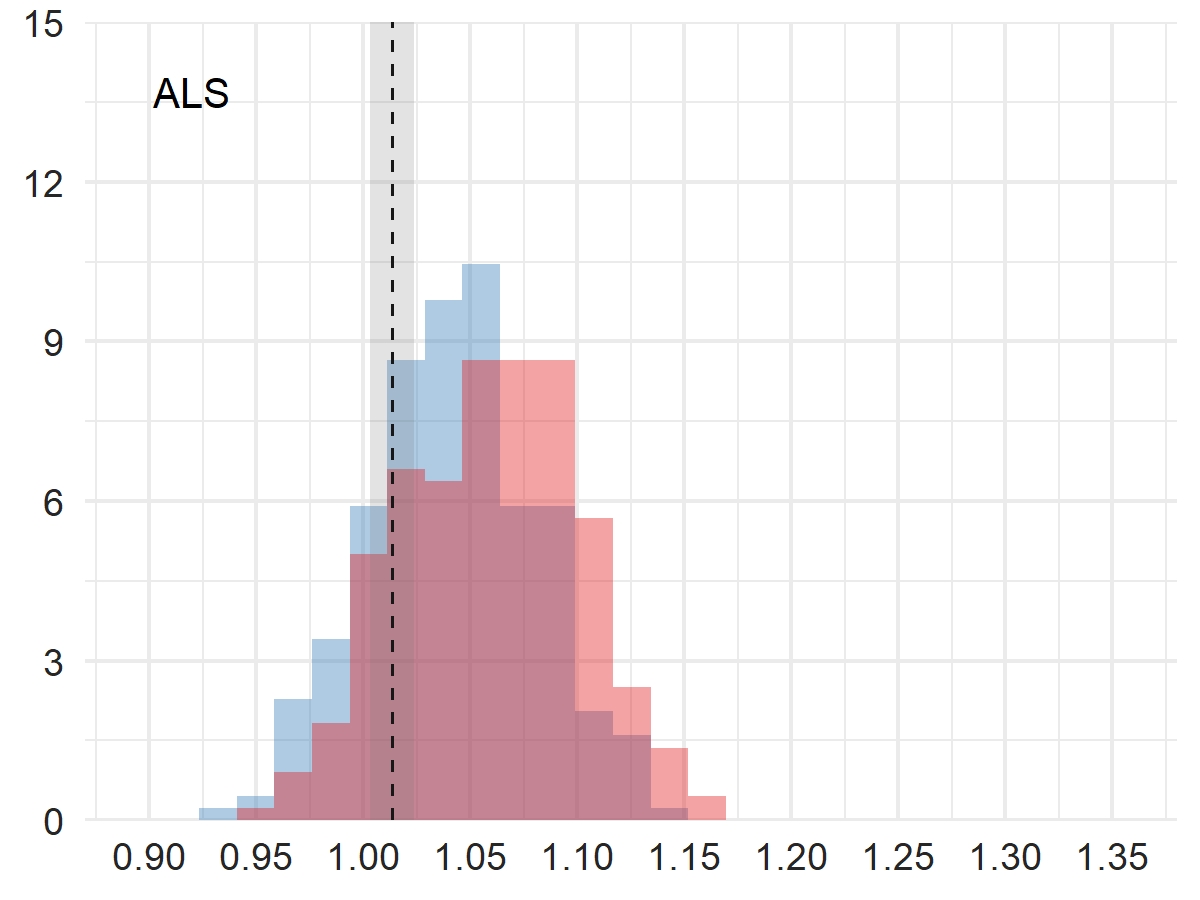

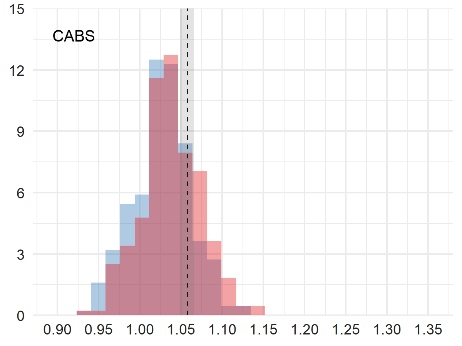

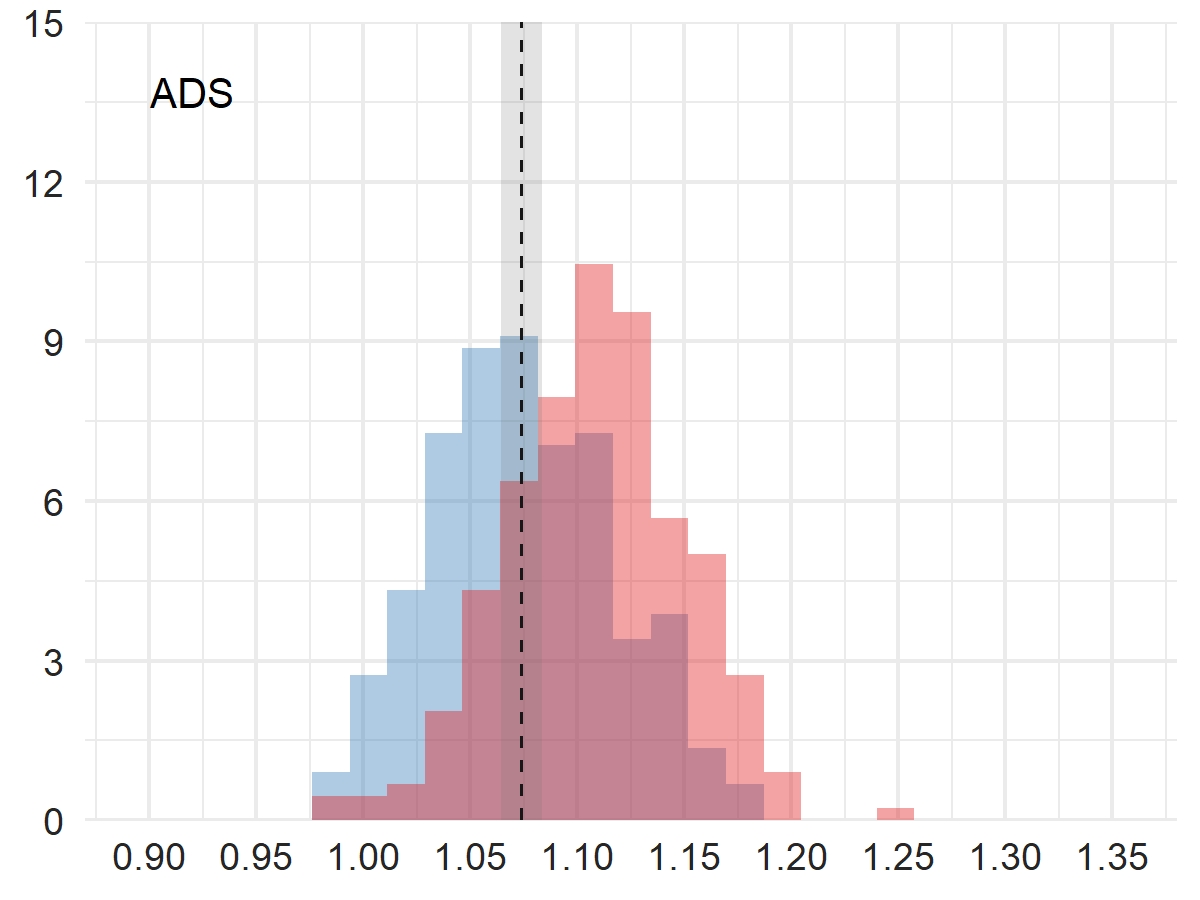

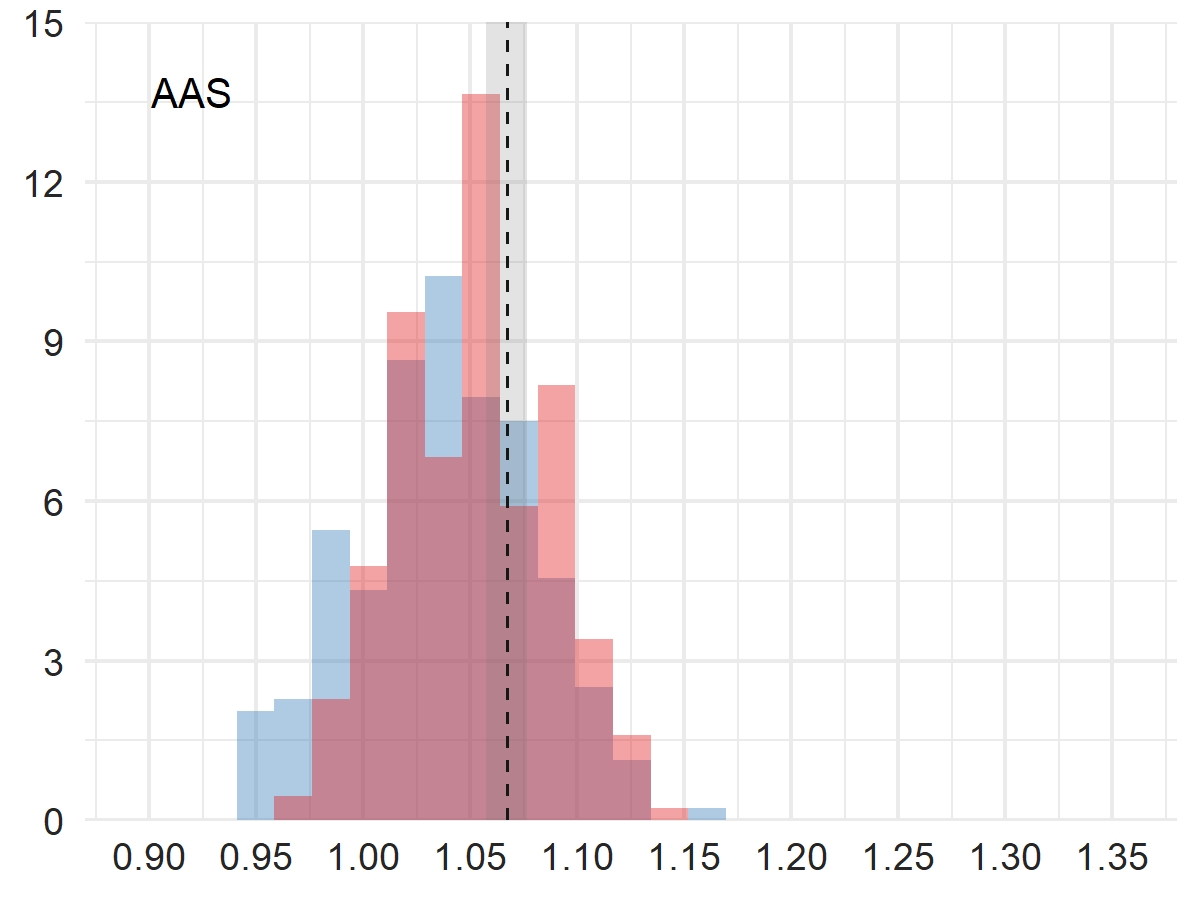

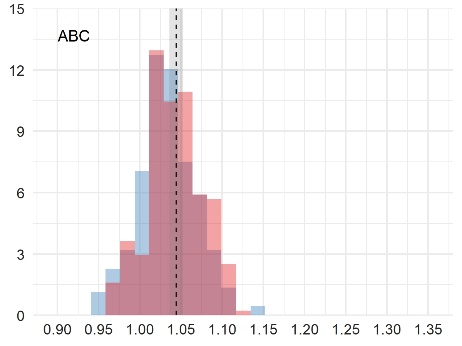

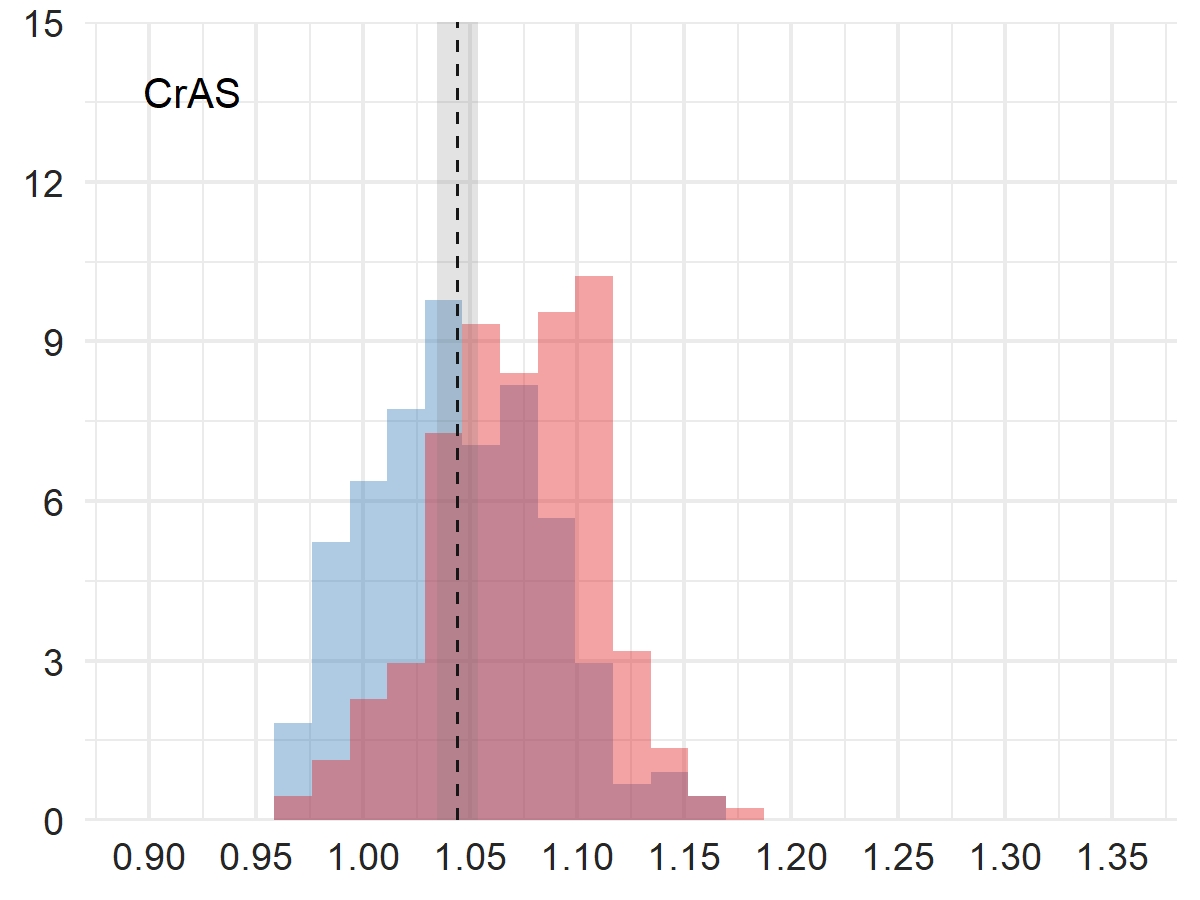

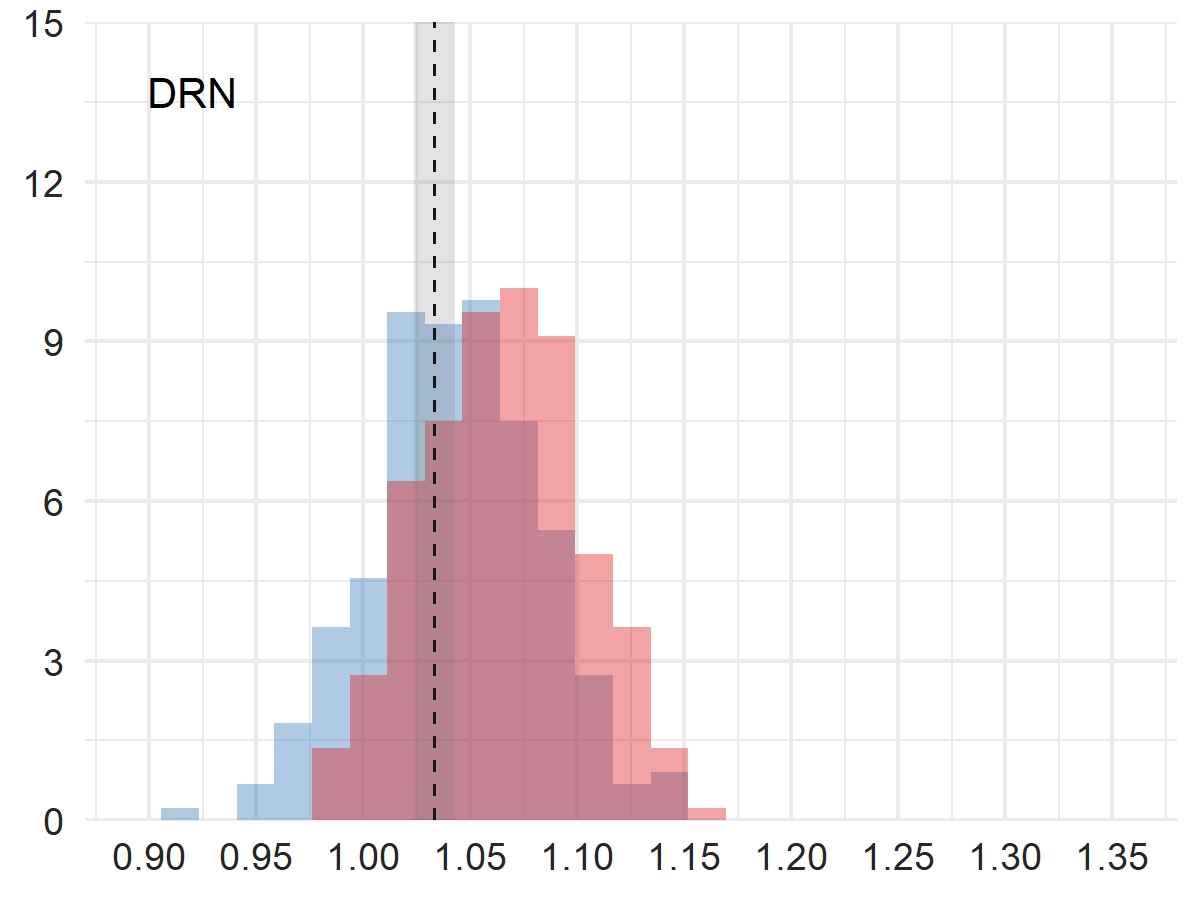

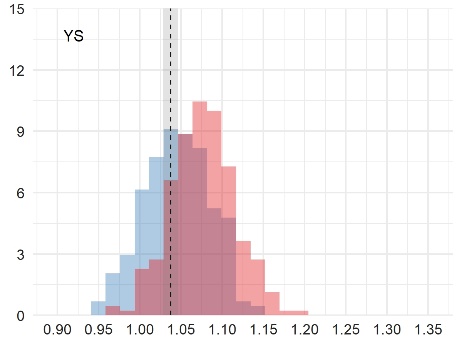

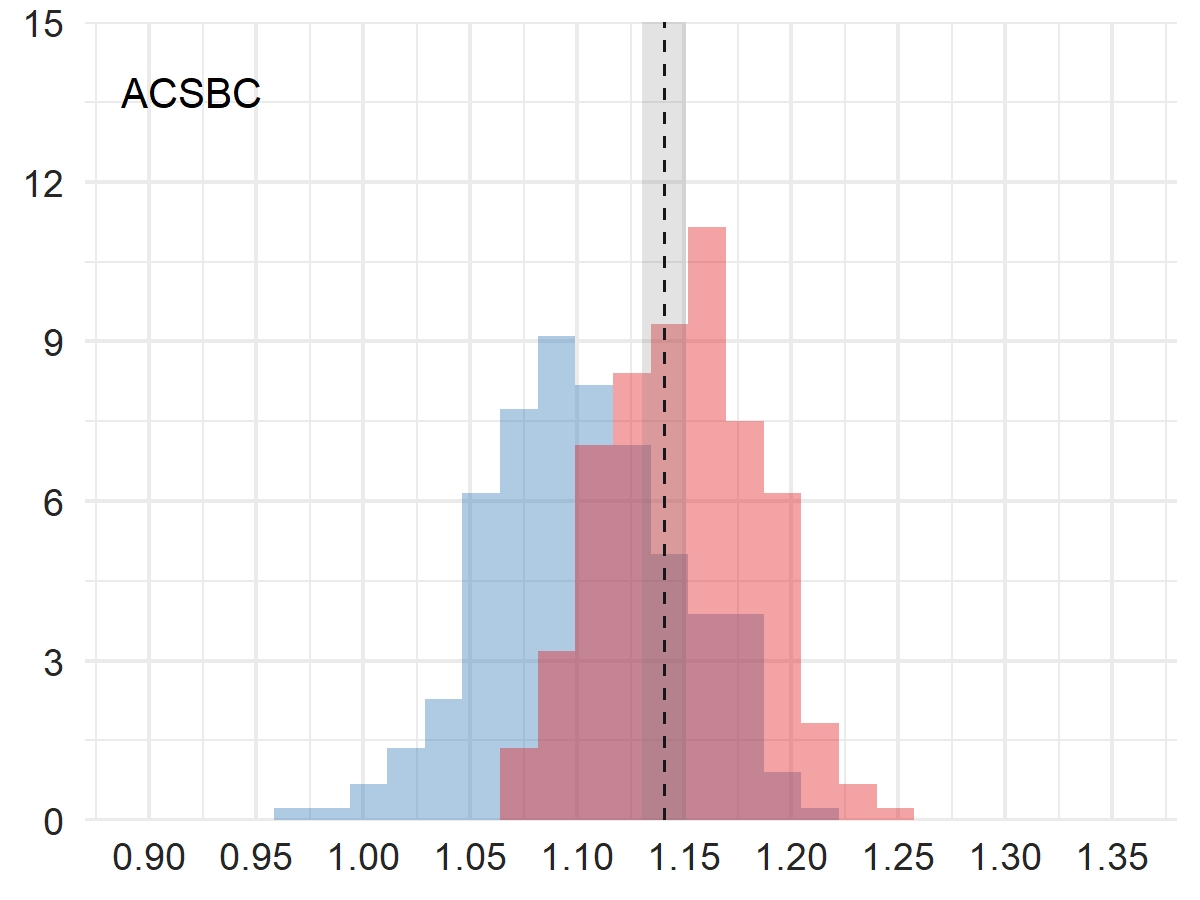

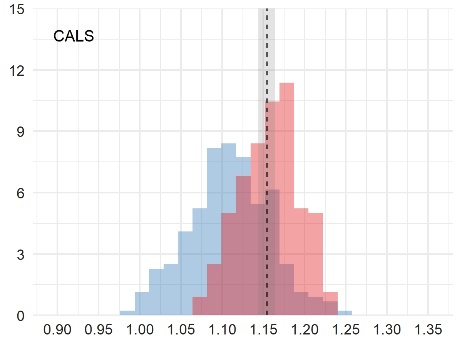

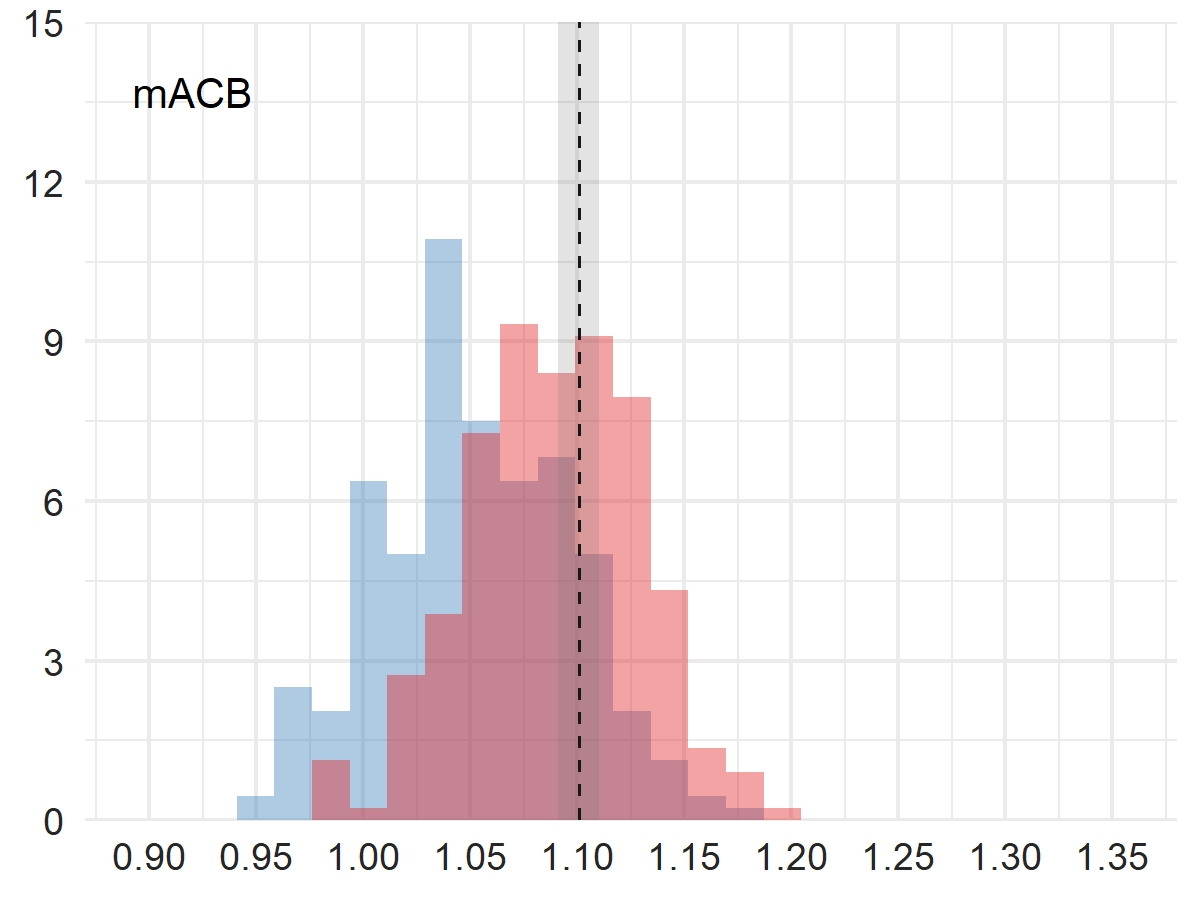

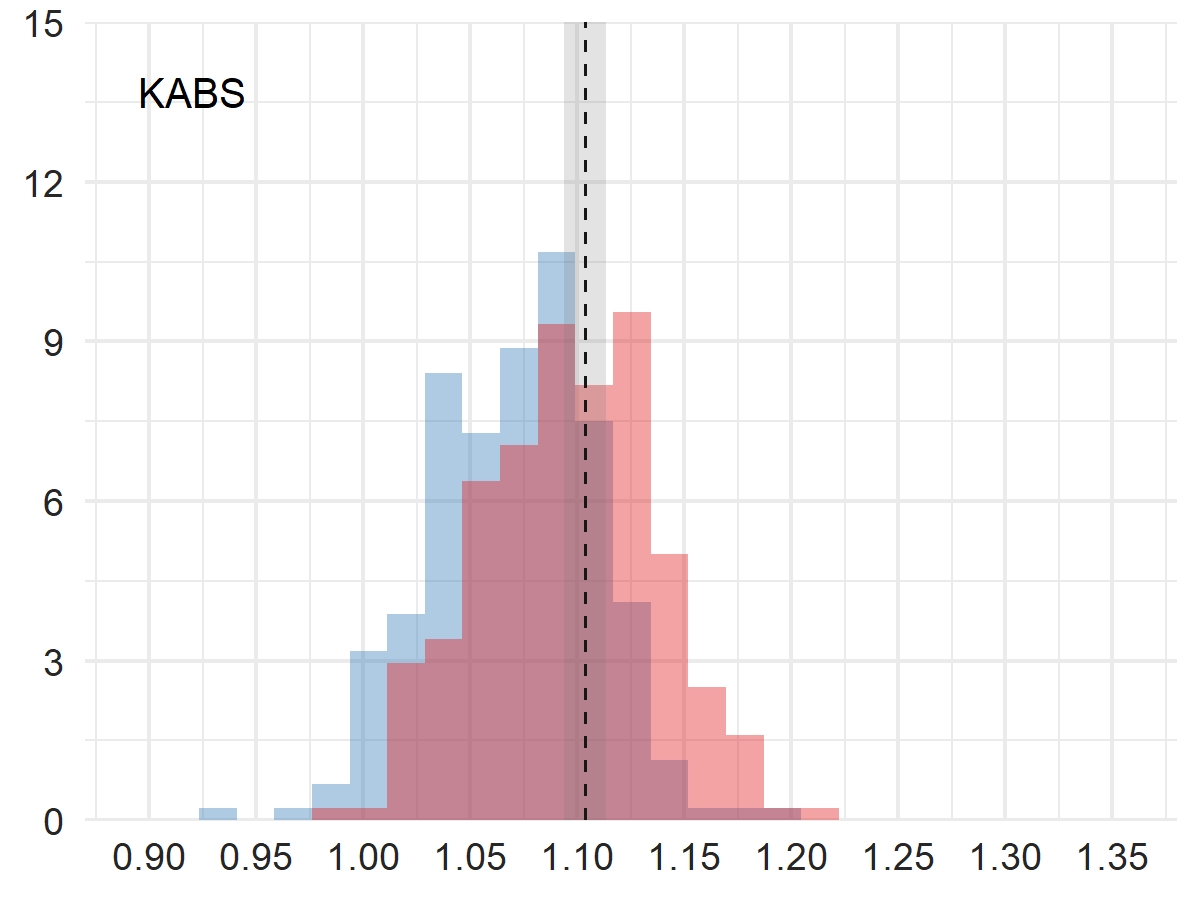

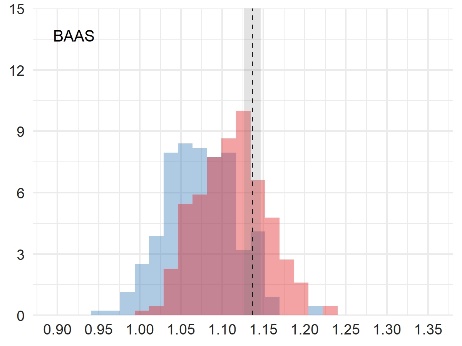

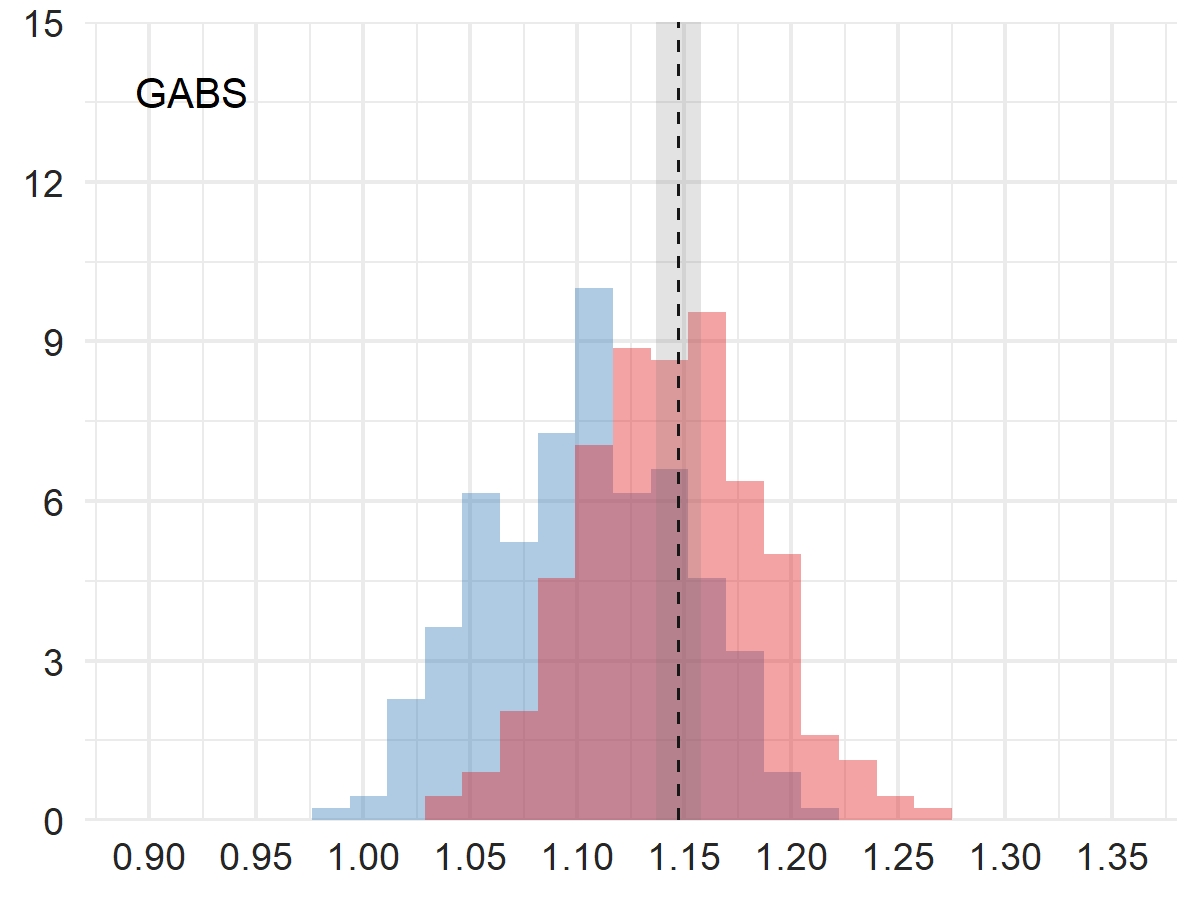

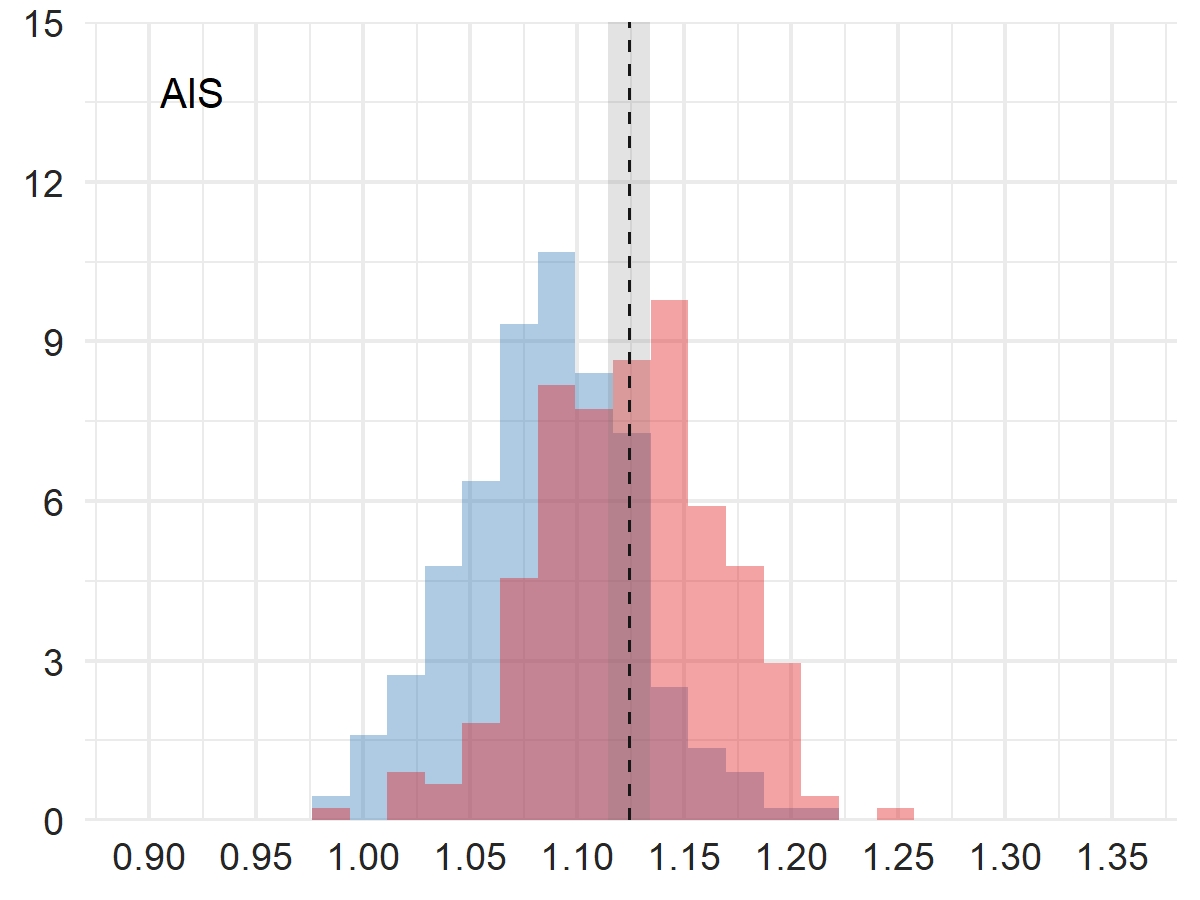

**Dementia**

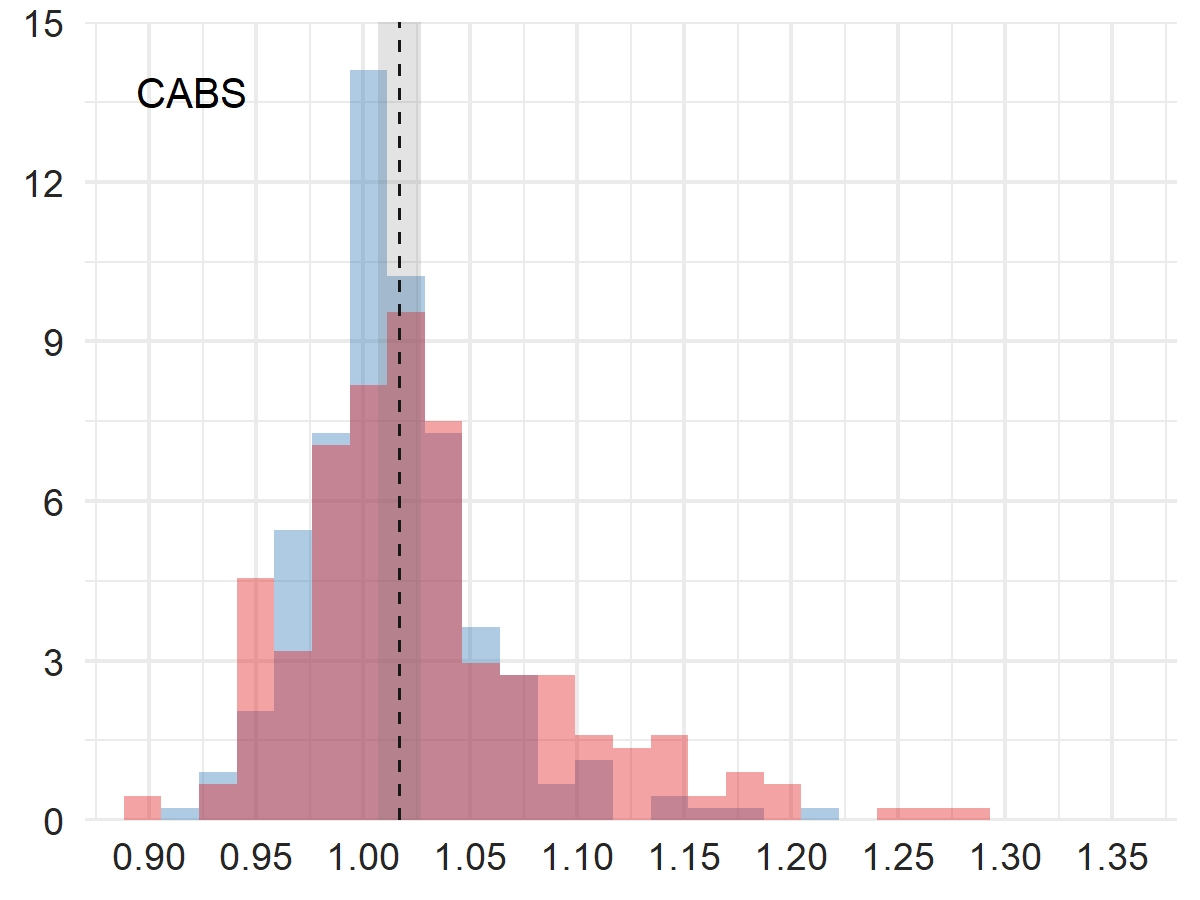

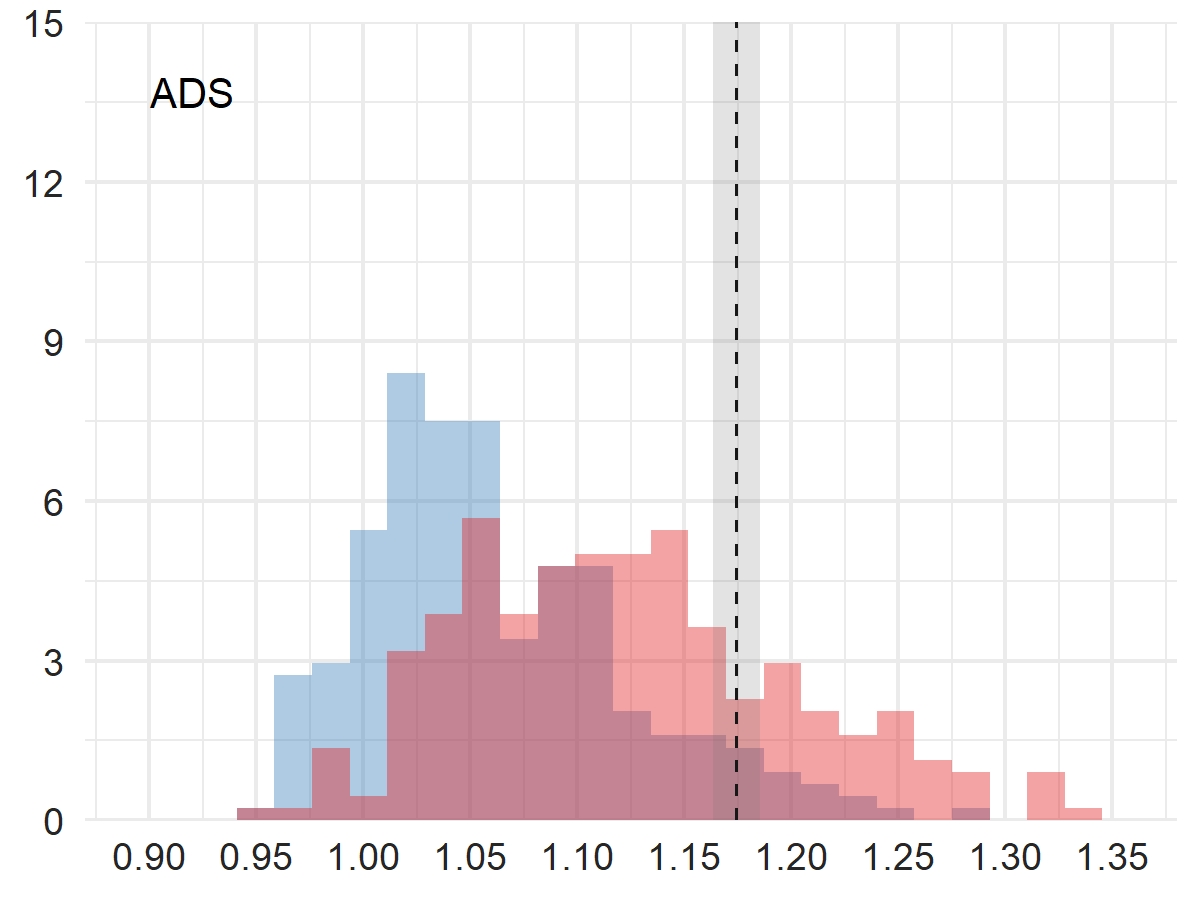

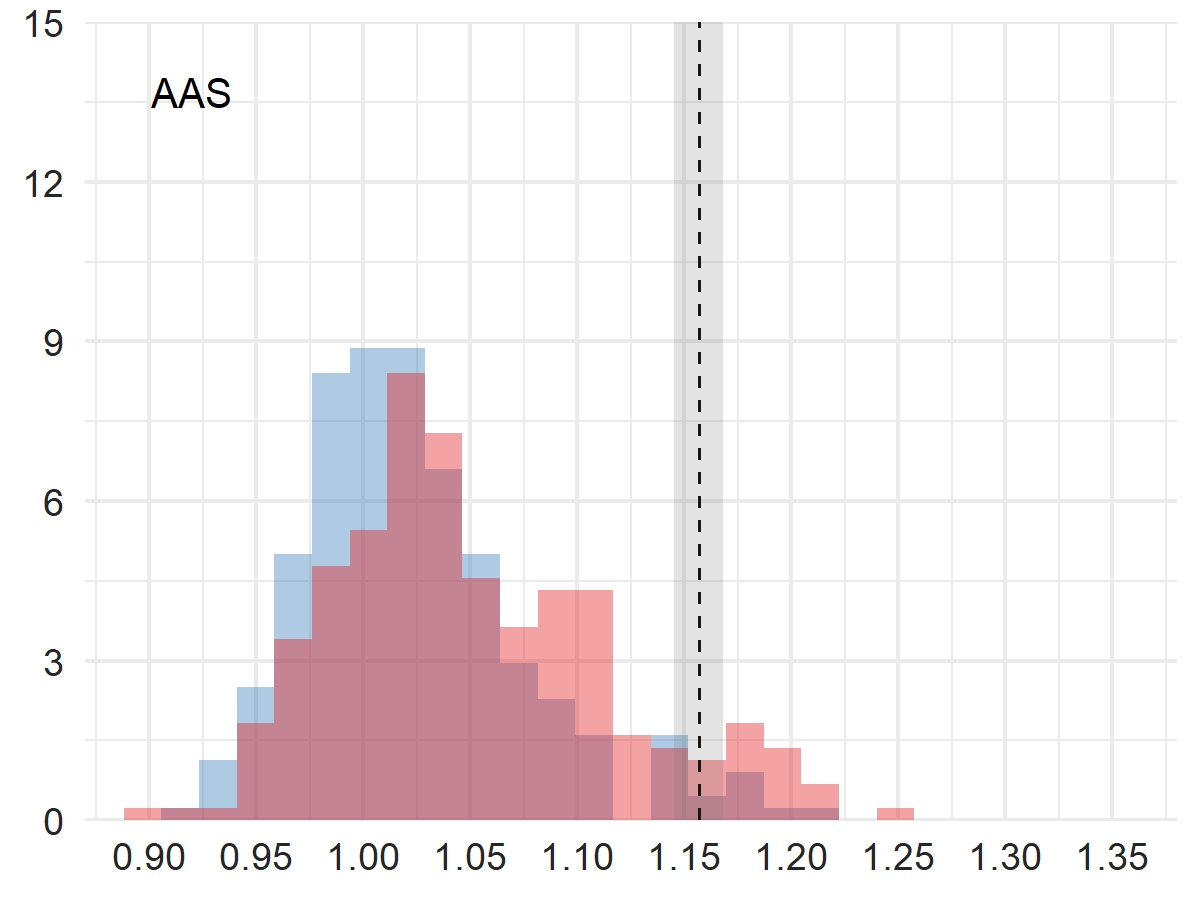

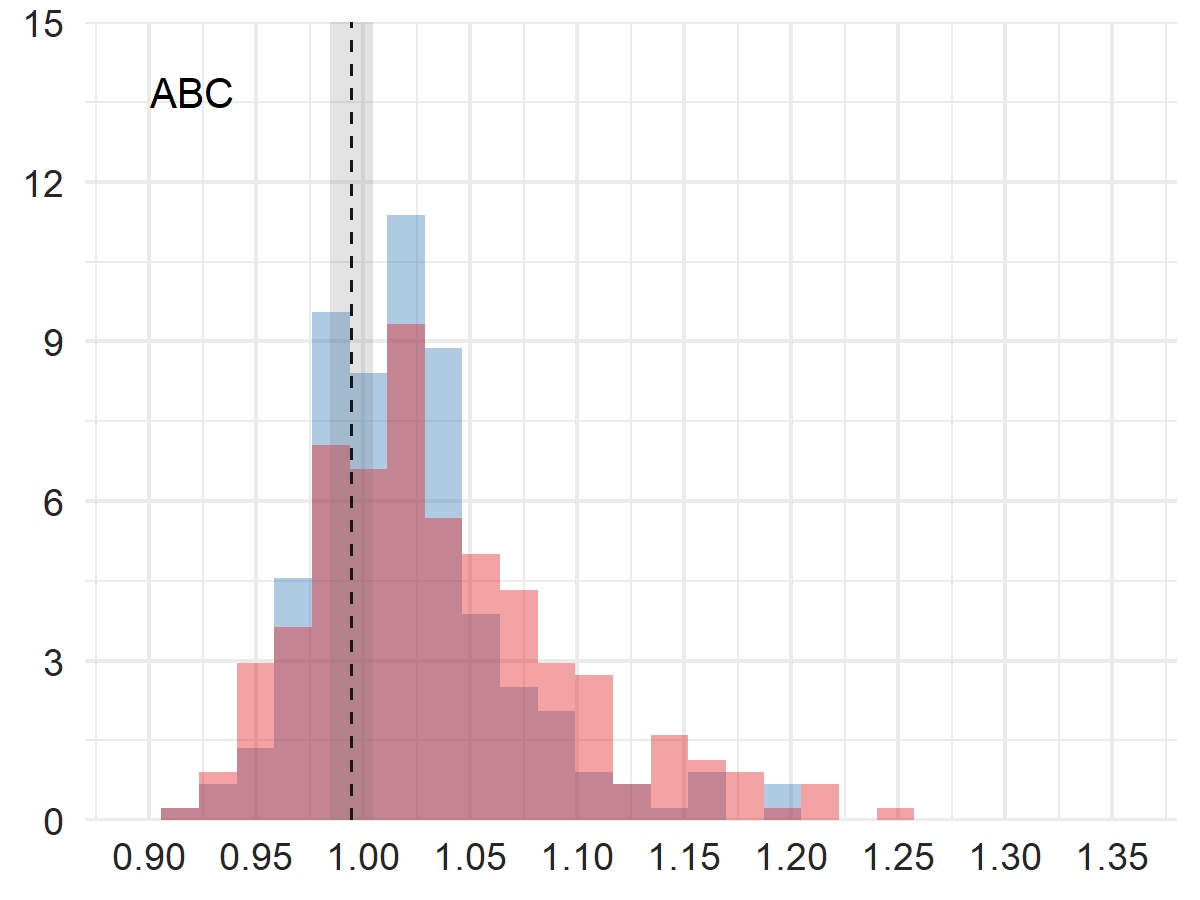

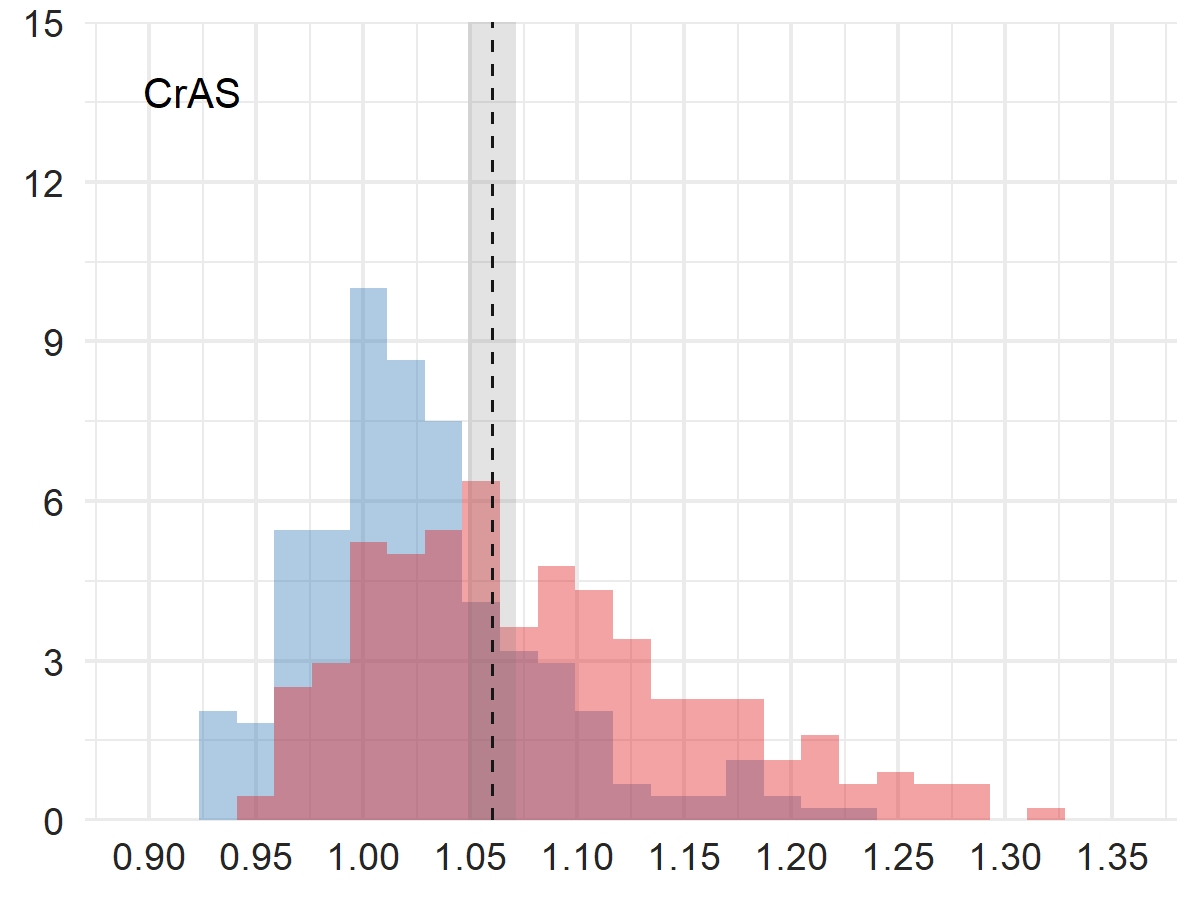

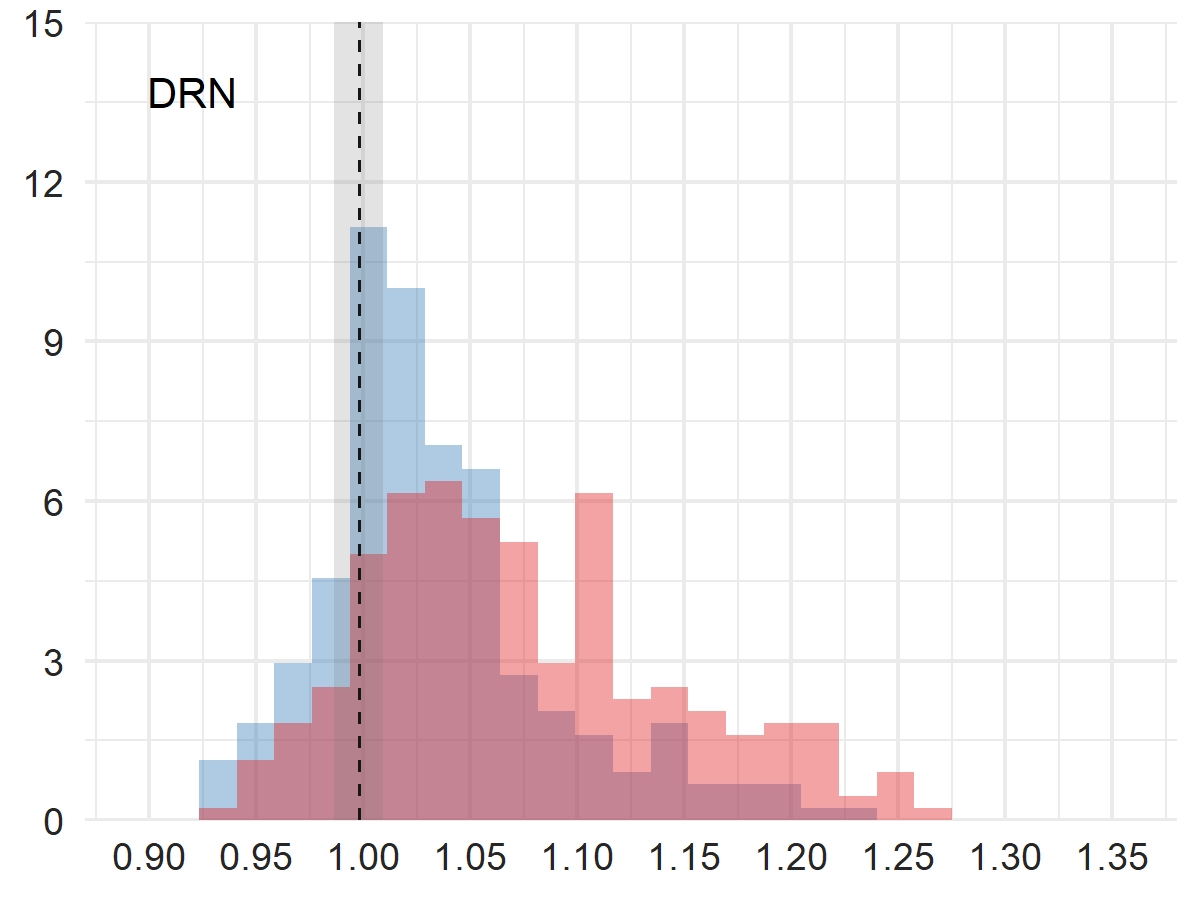

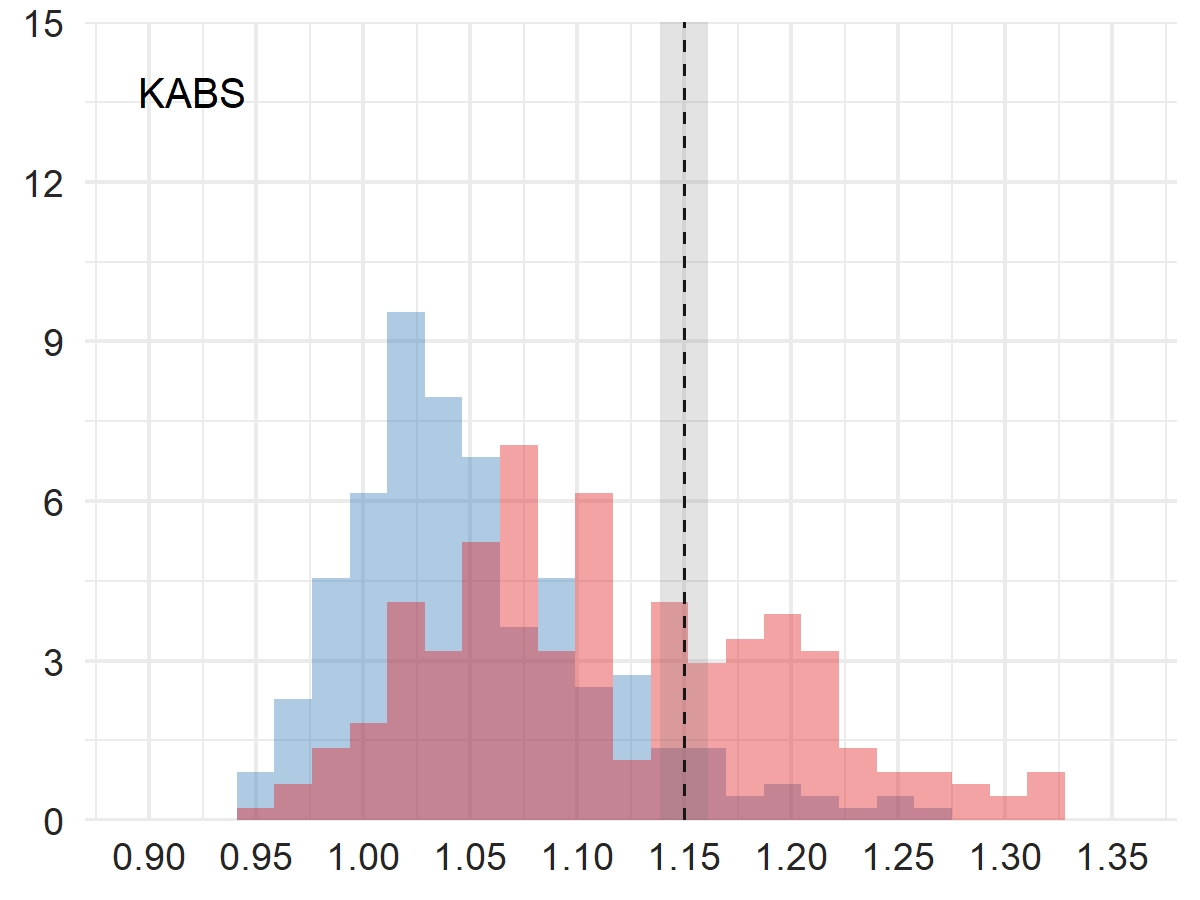

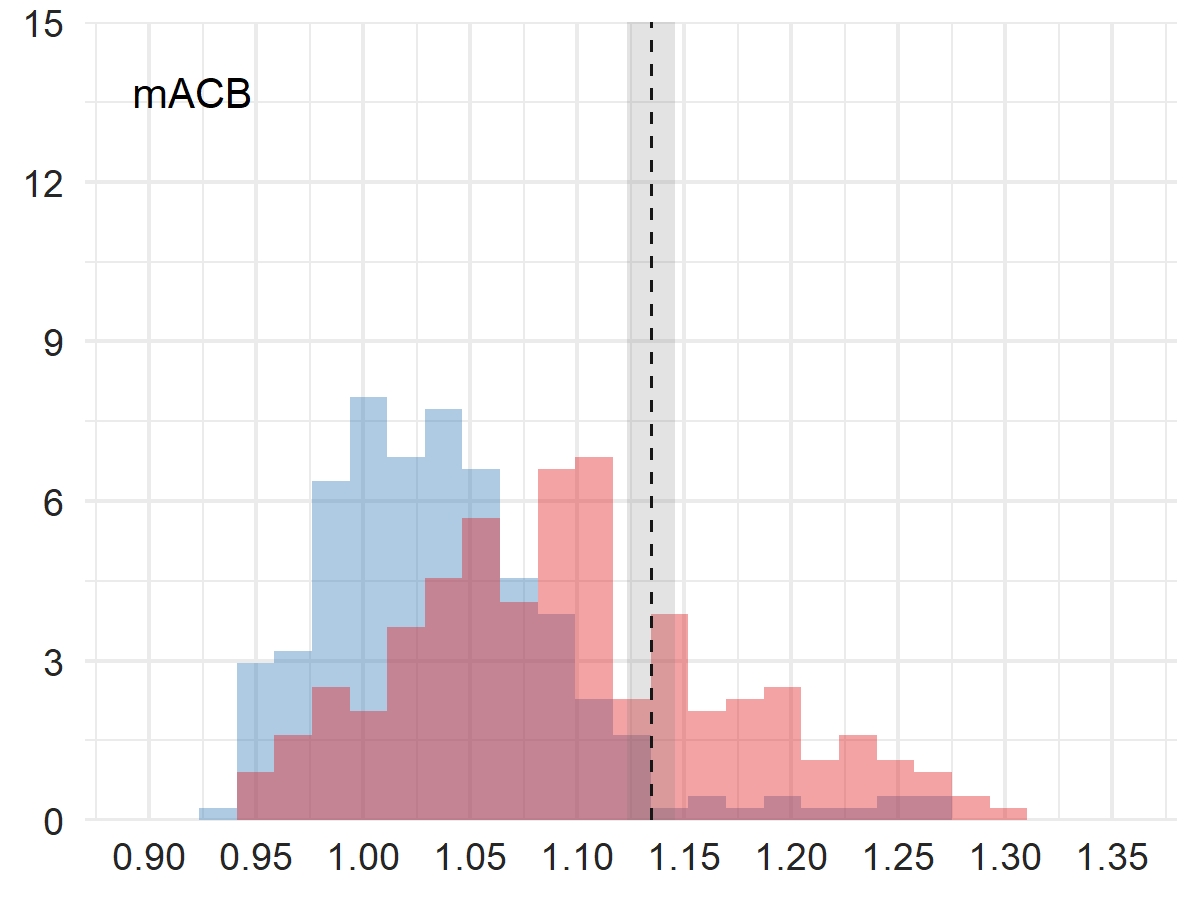

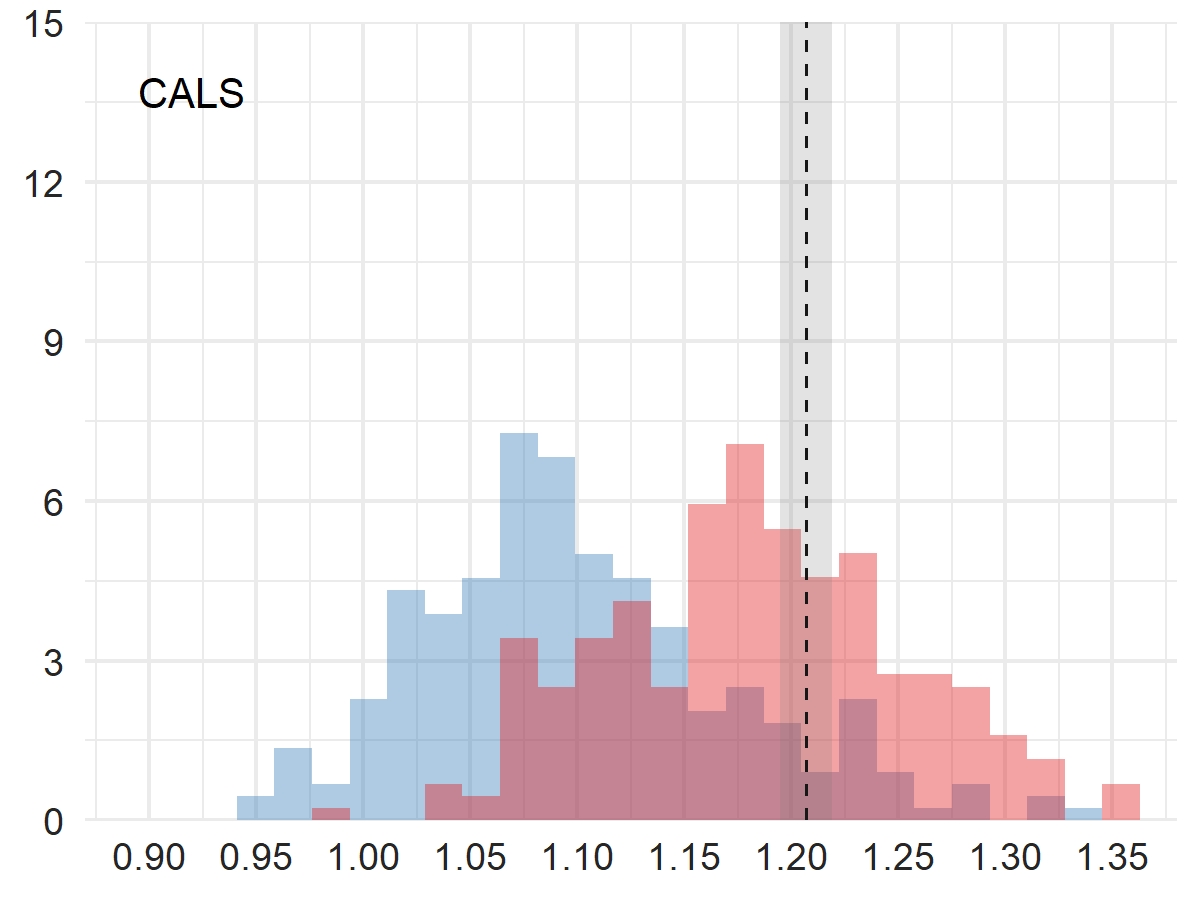

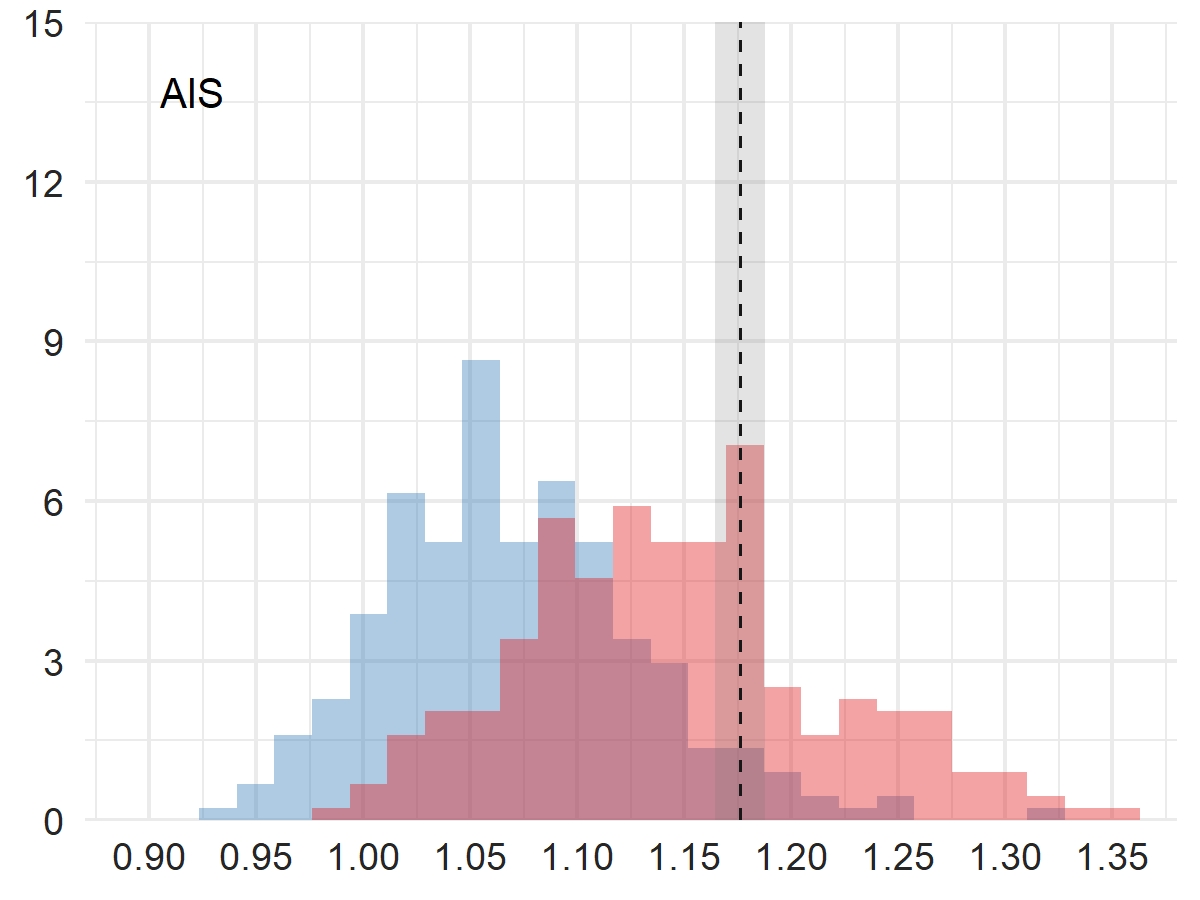

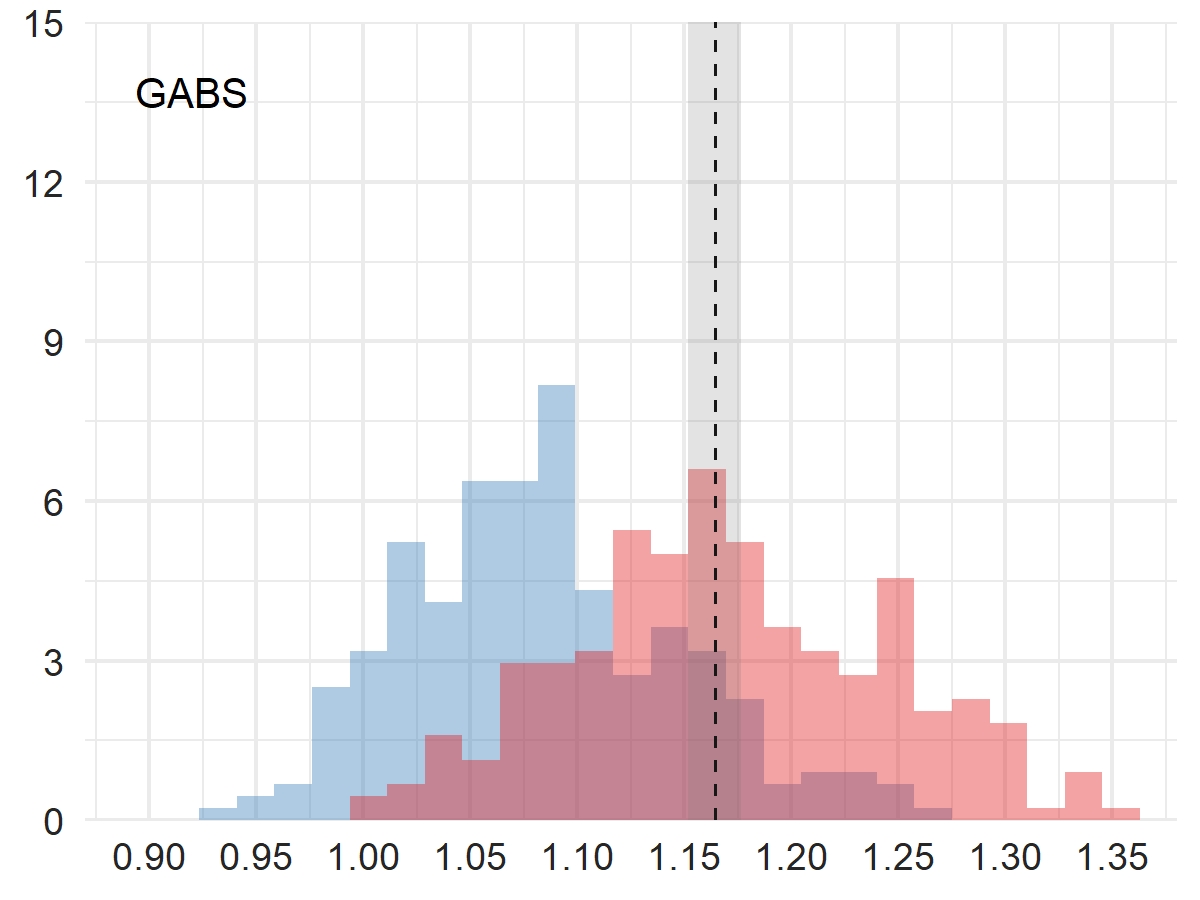

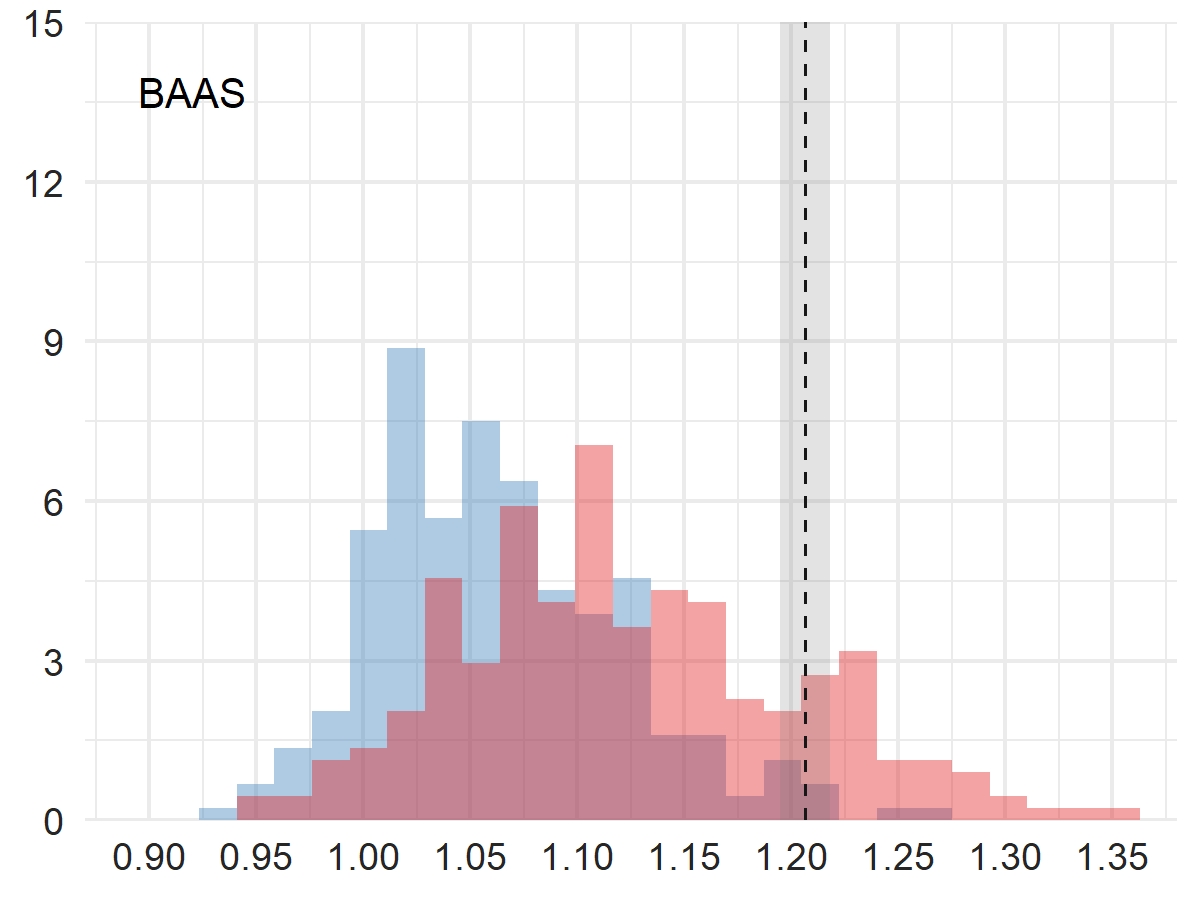

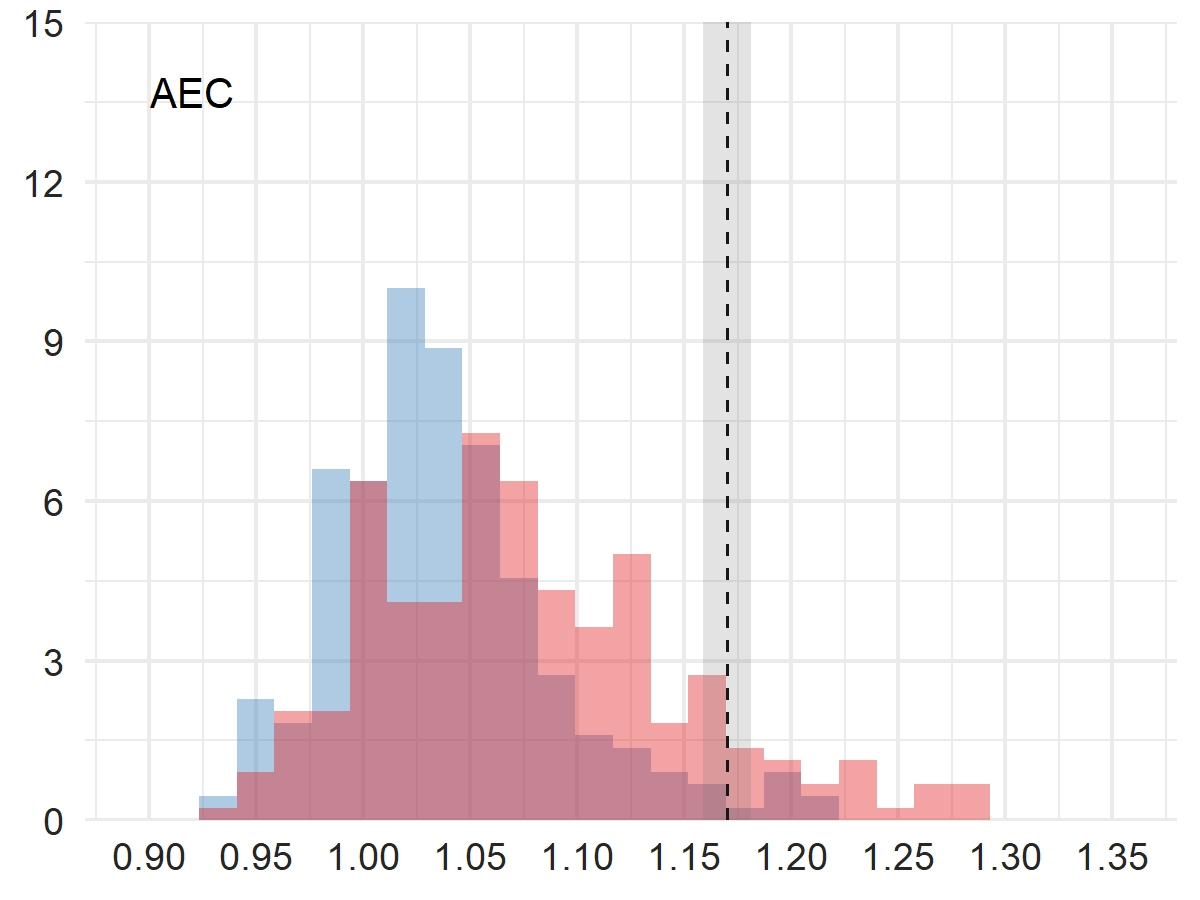

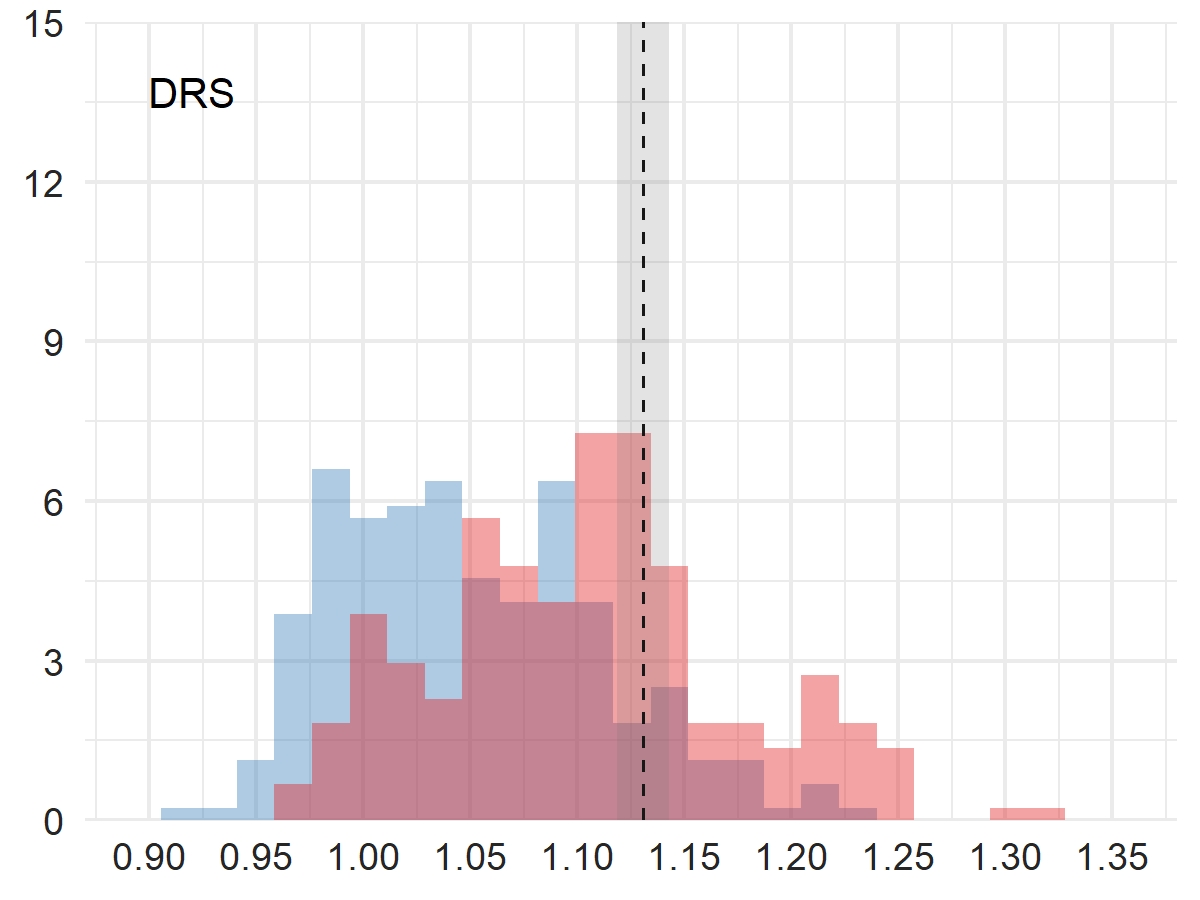

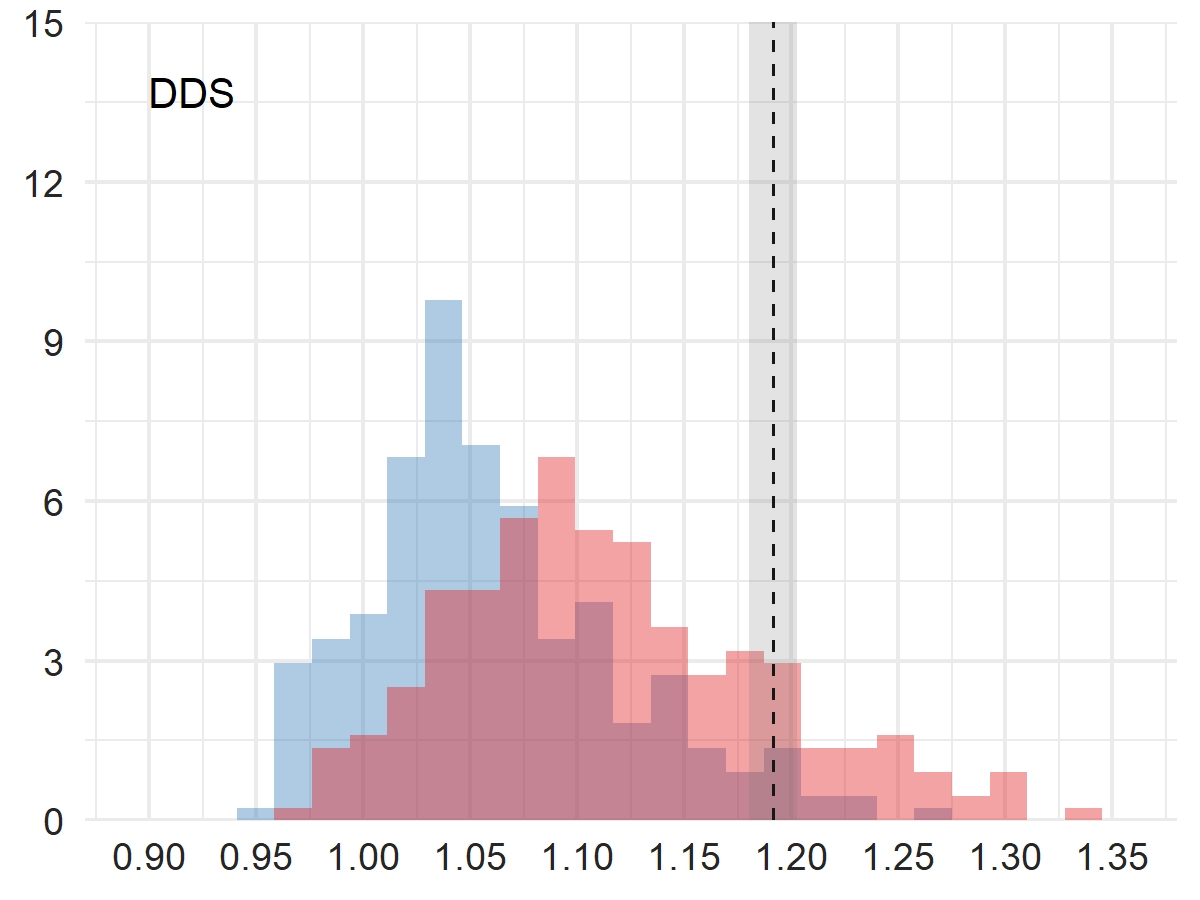

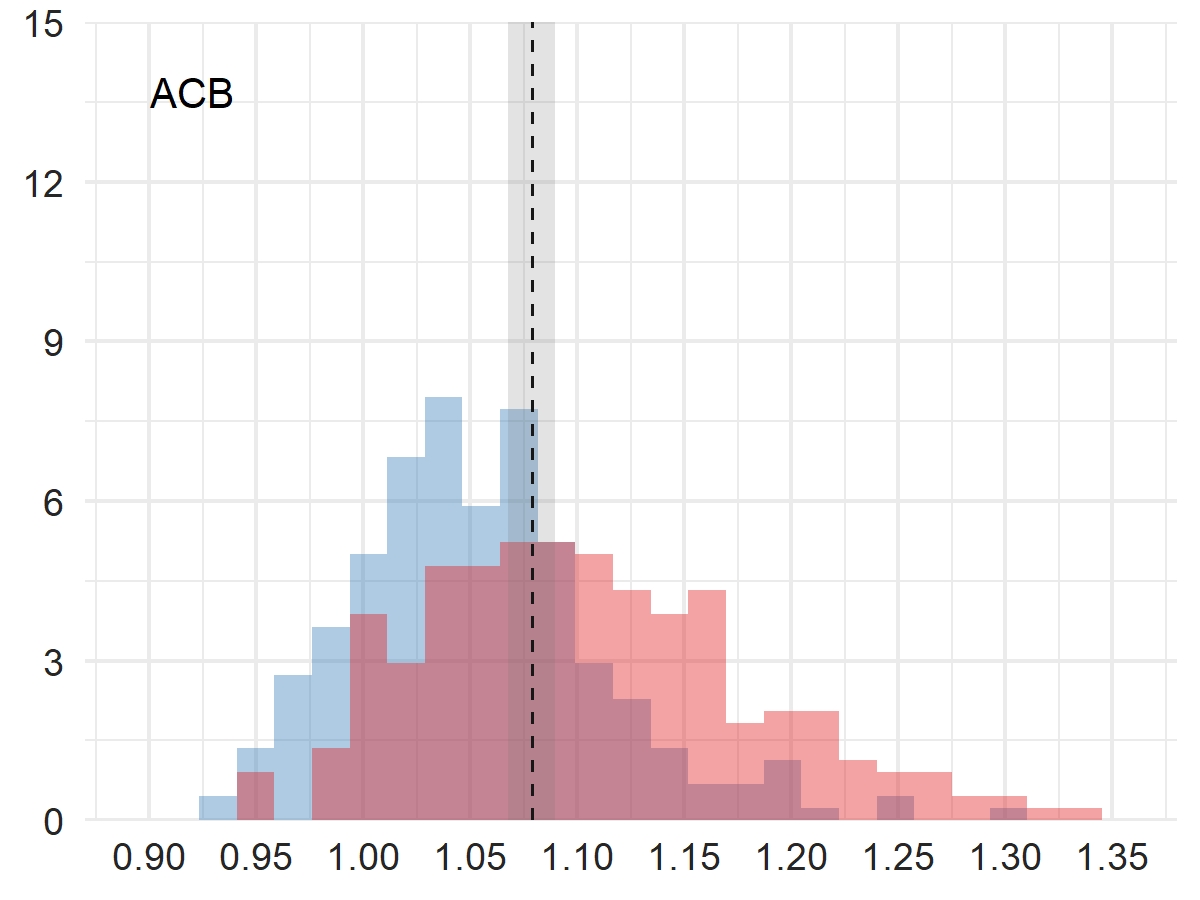

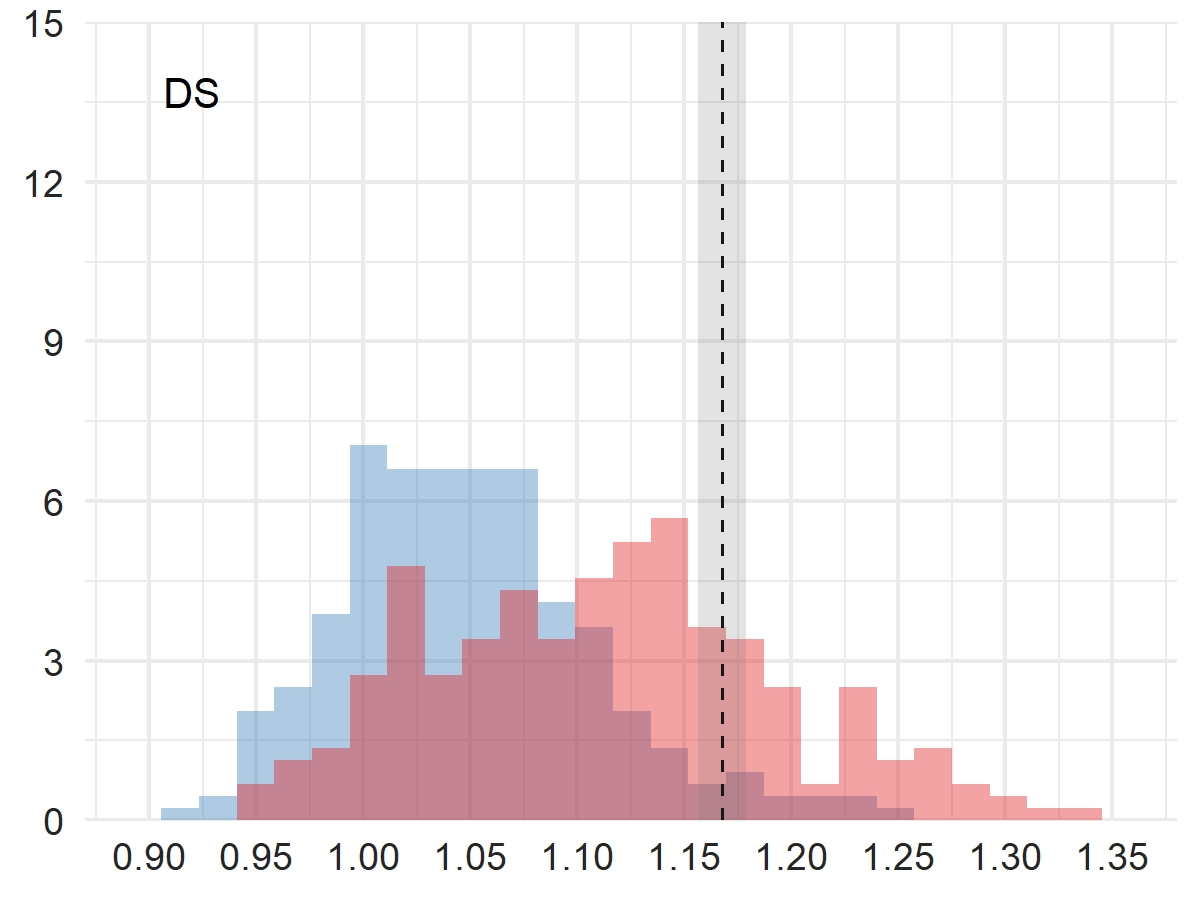

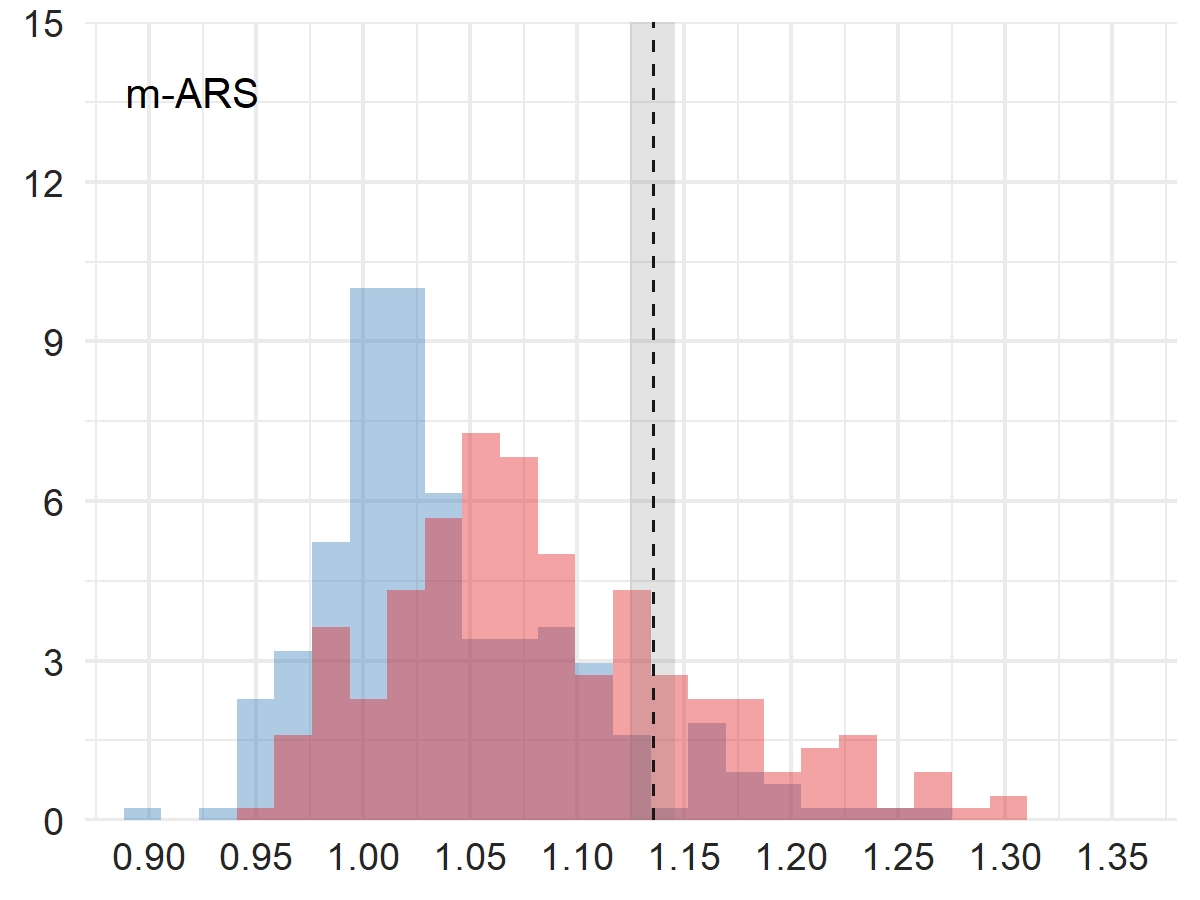

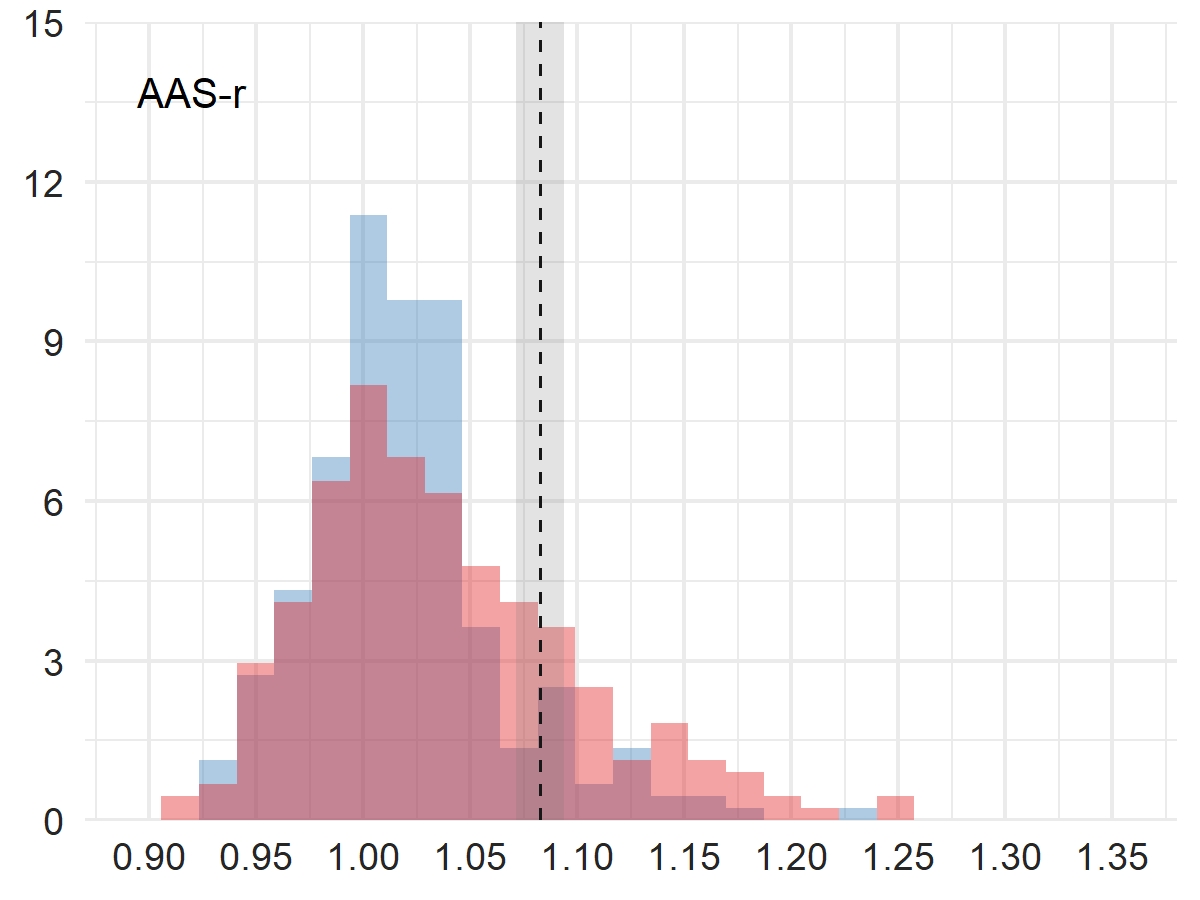

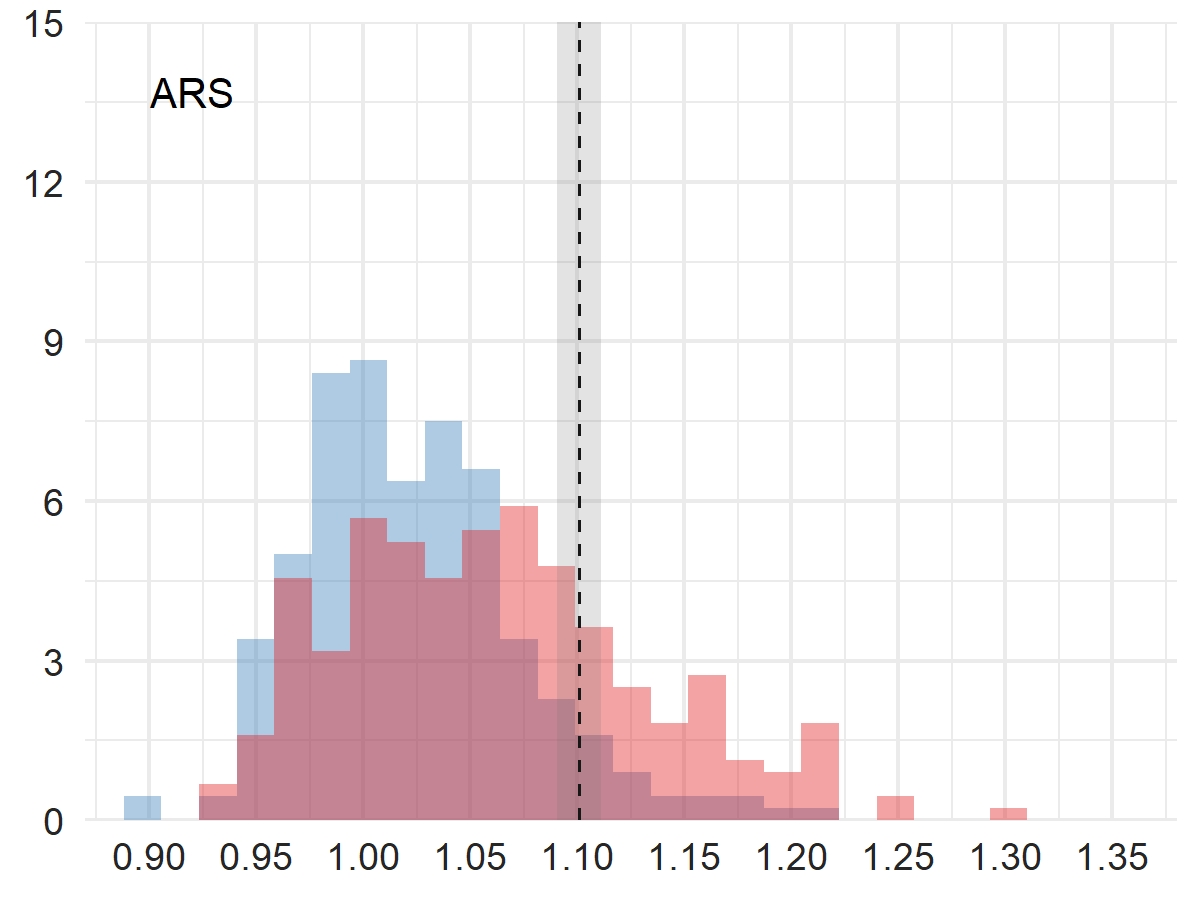

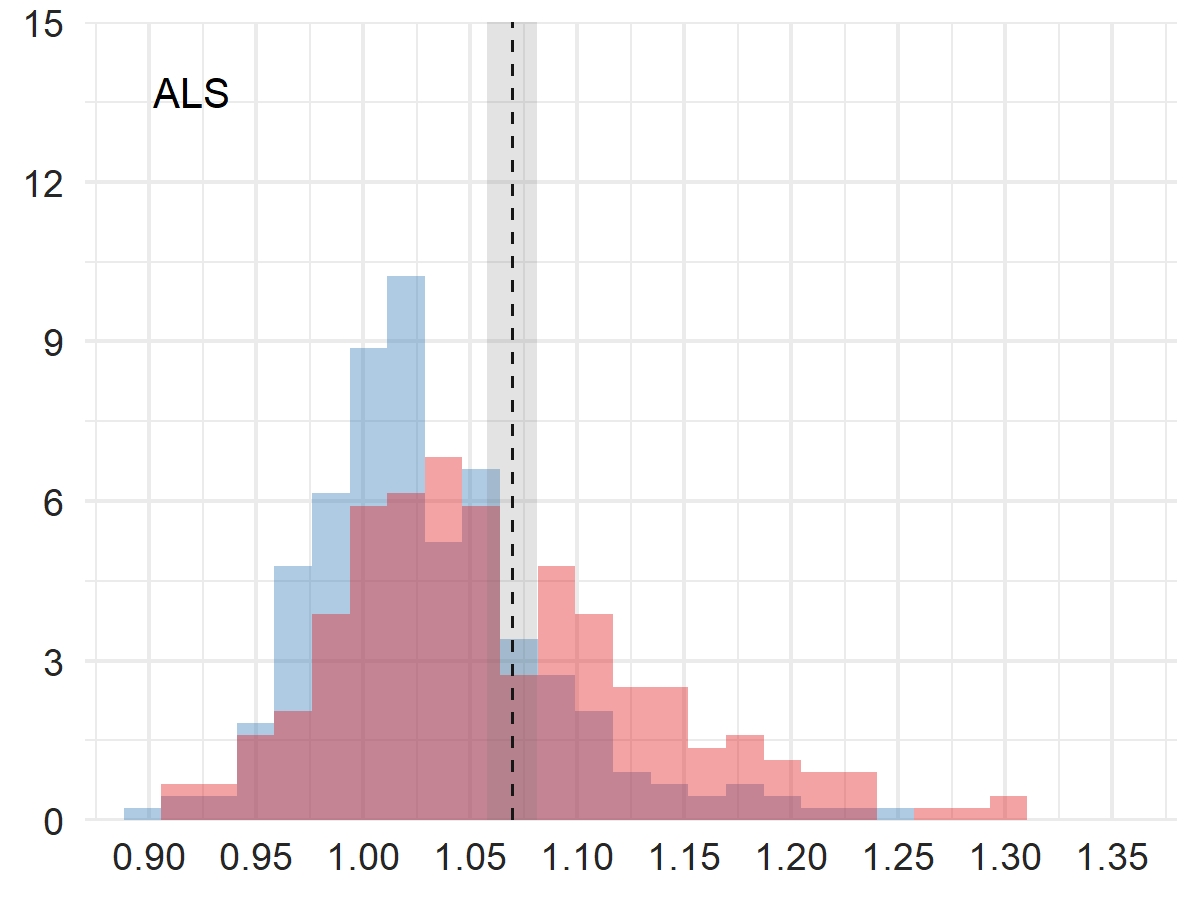

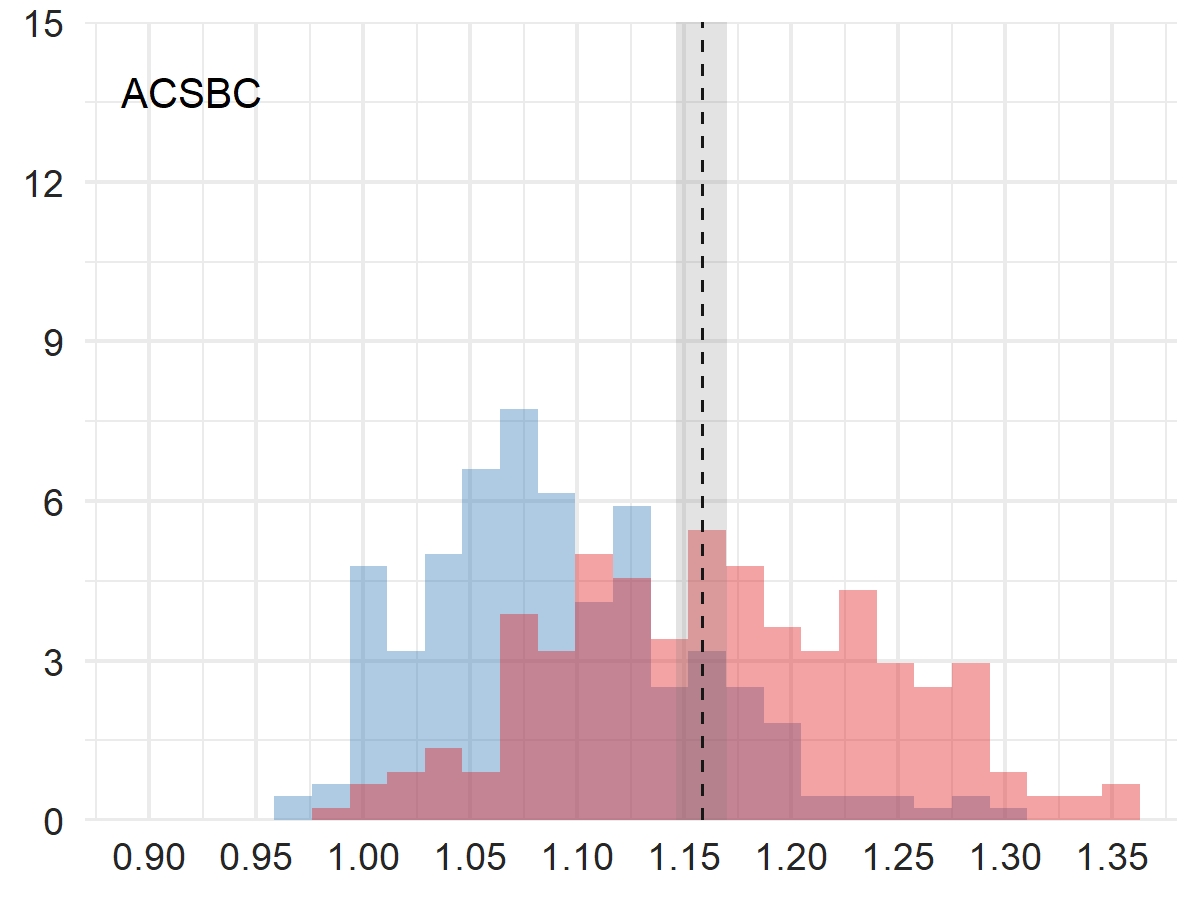

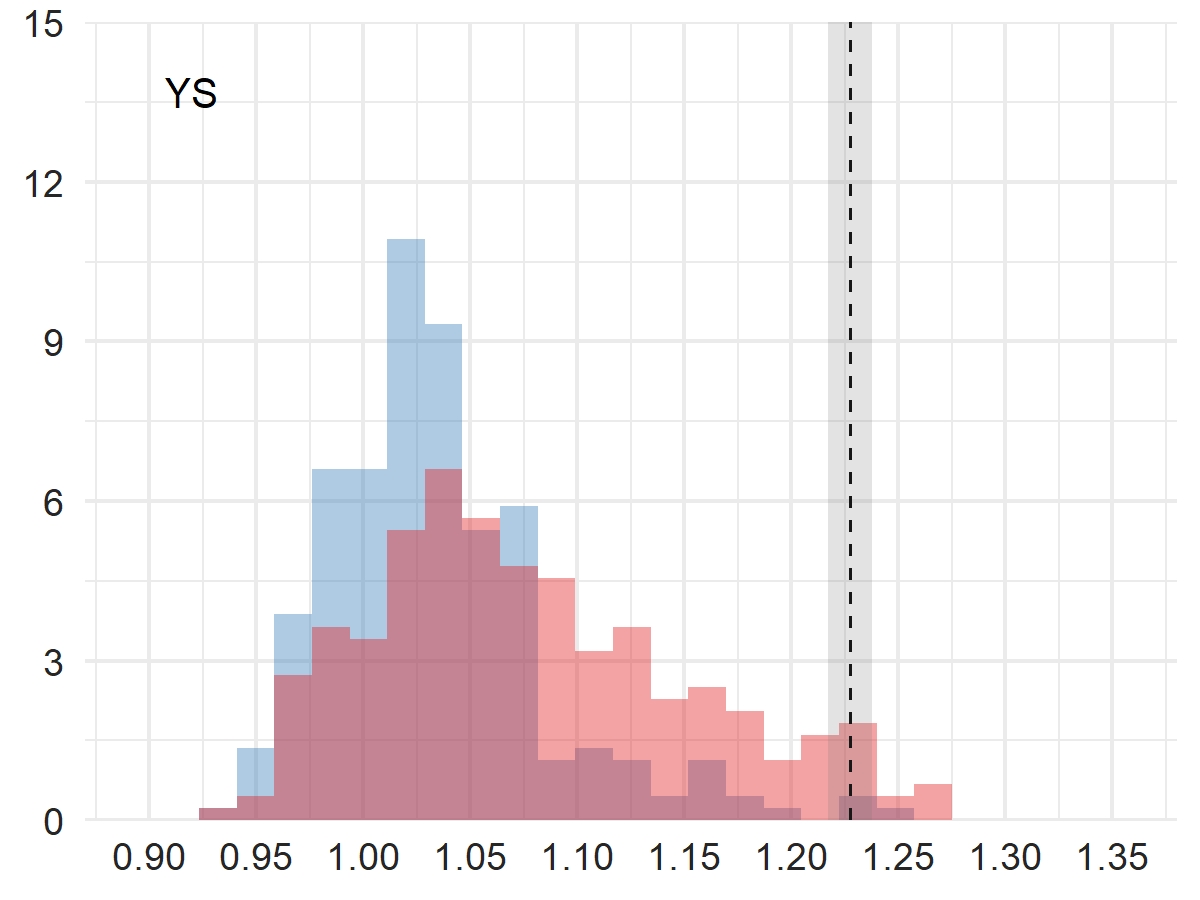

**Delirium**

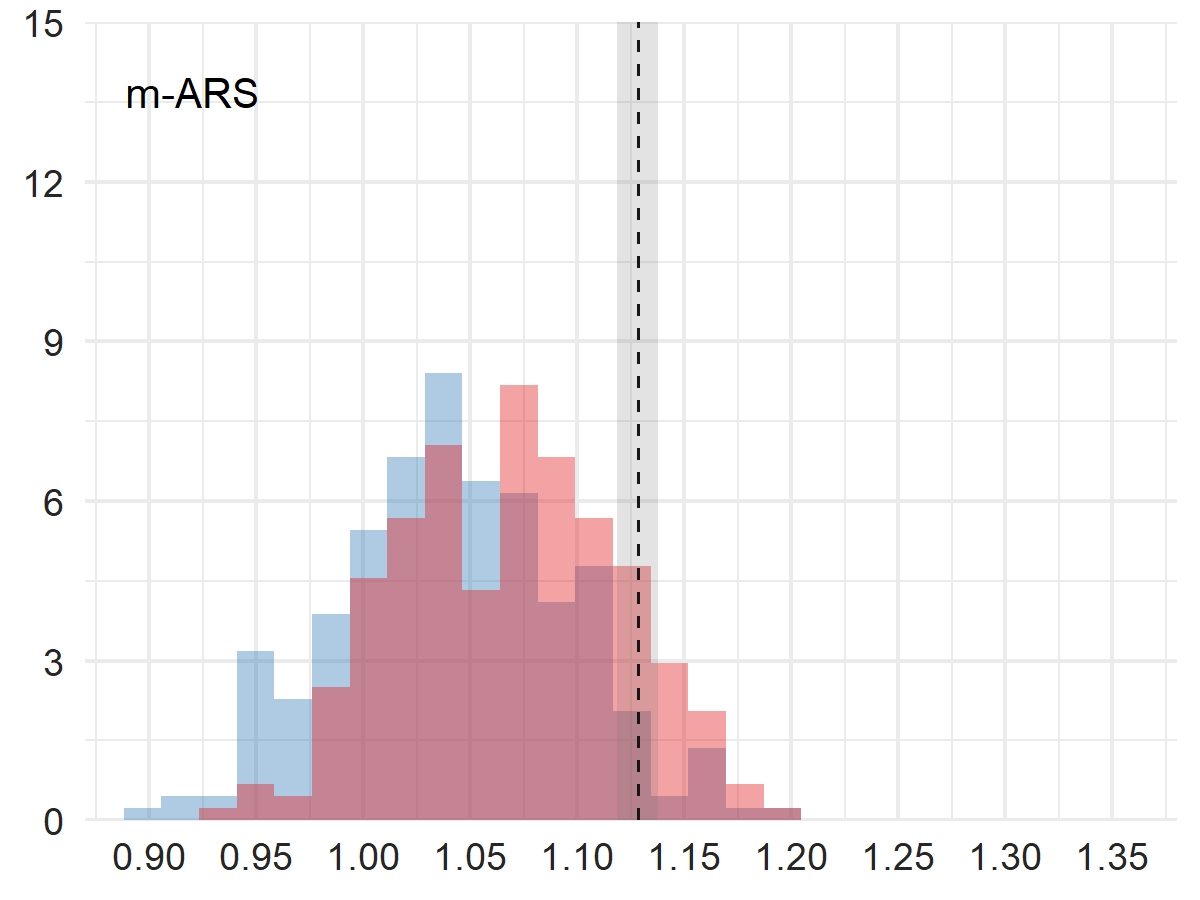

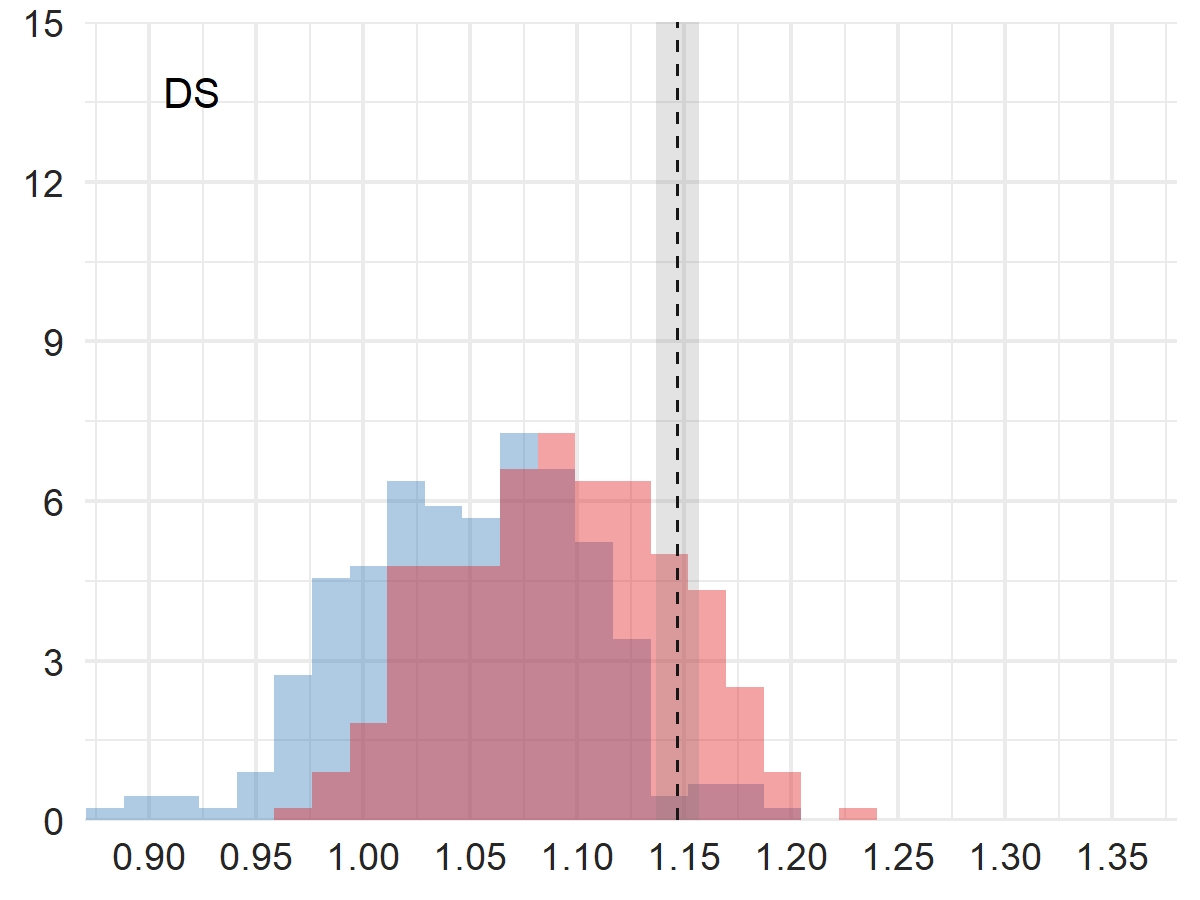

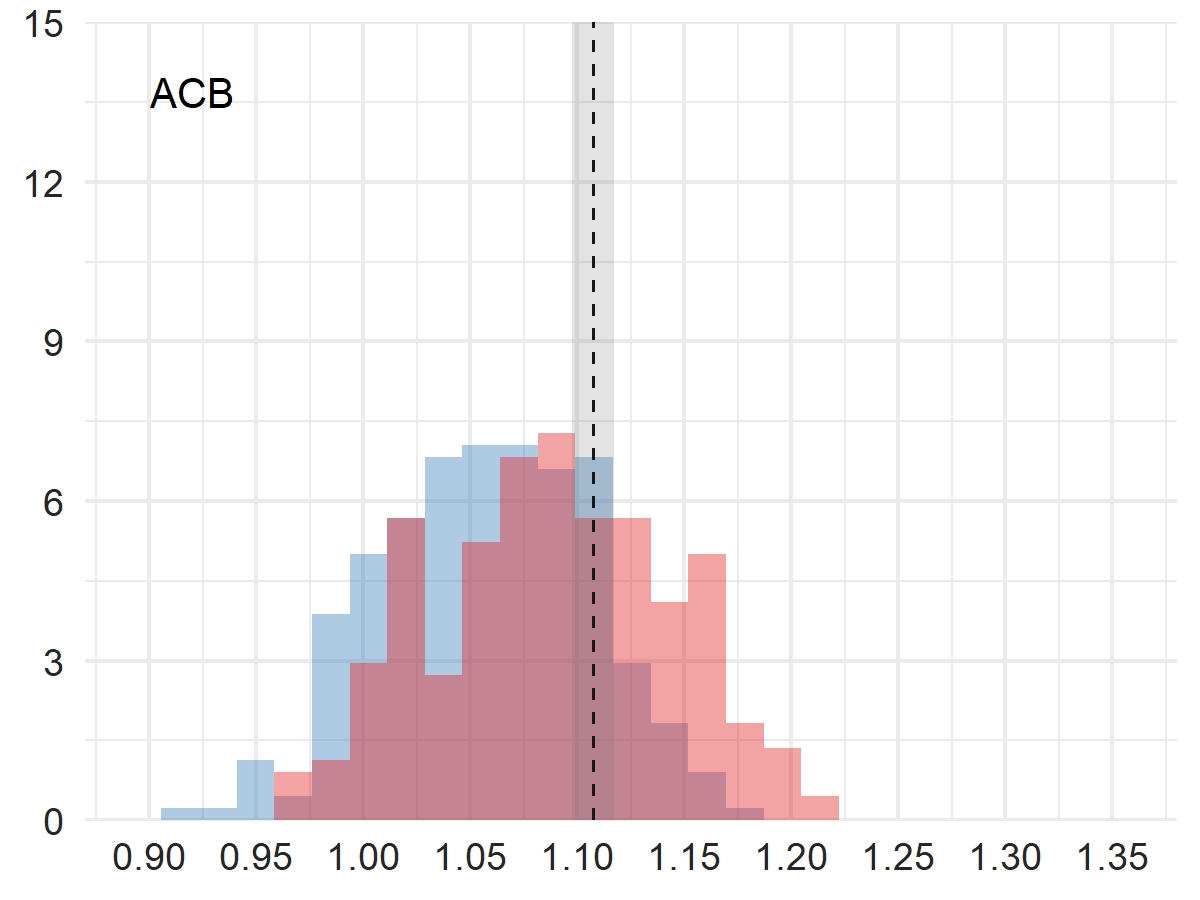

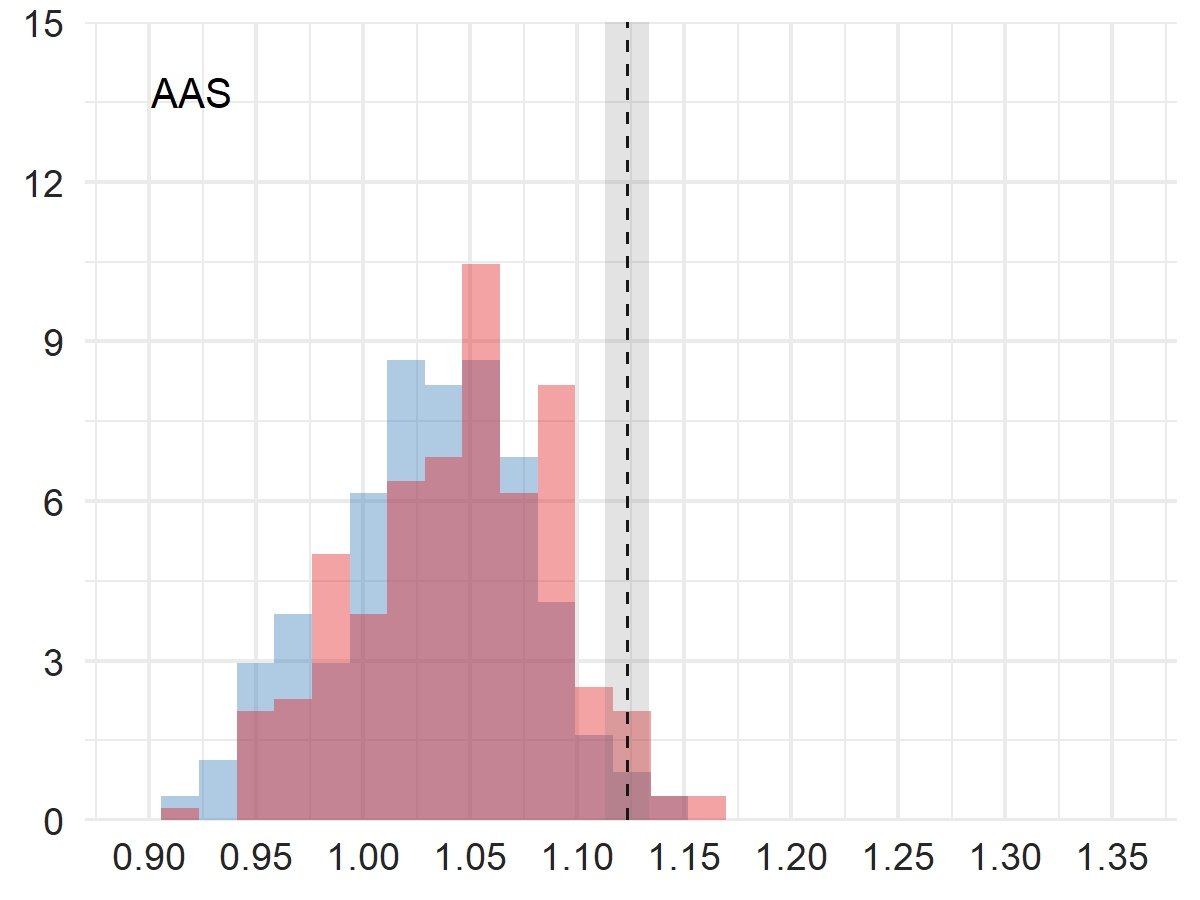

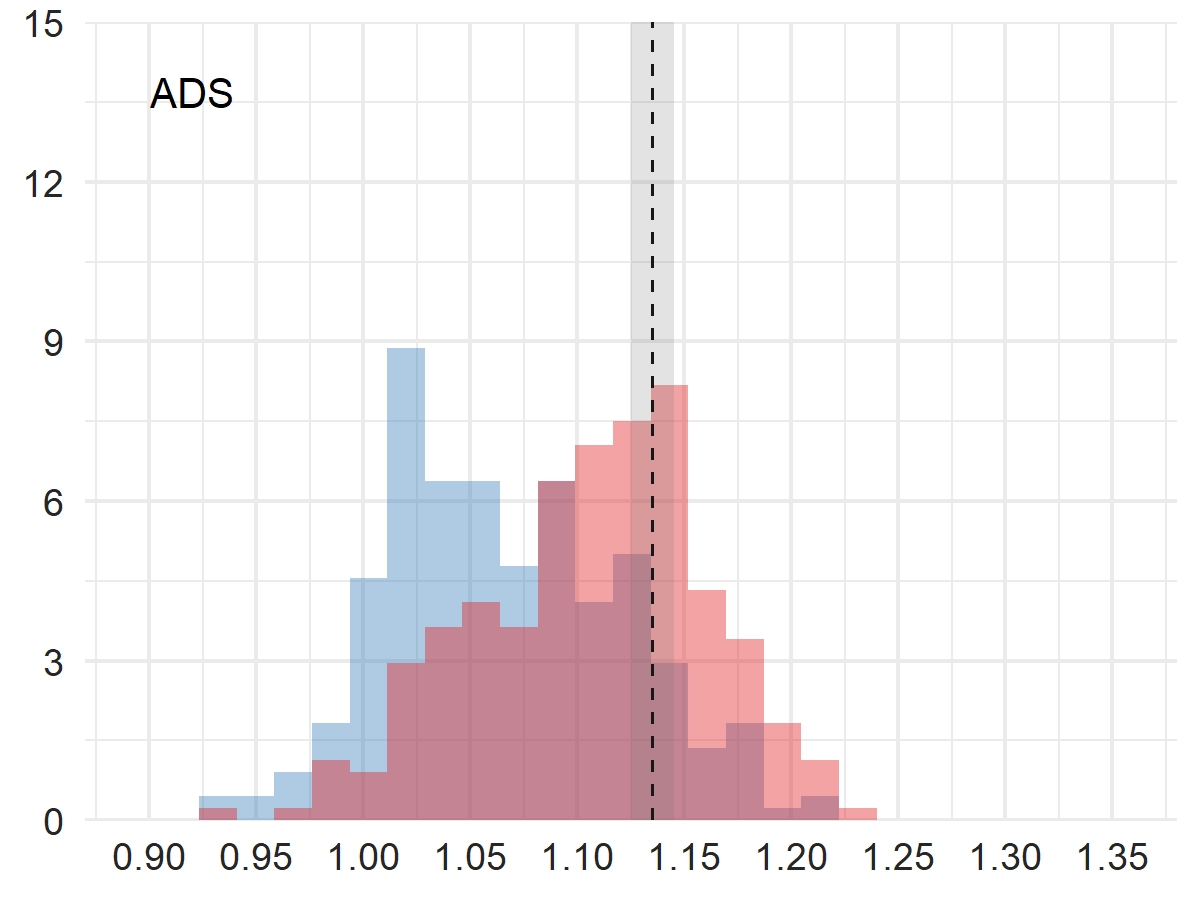

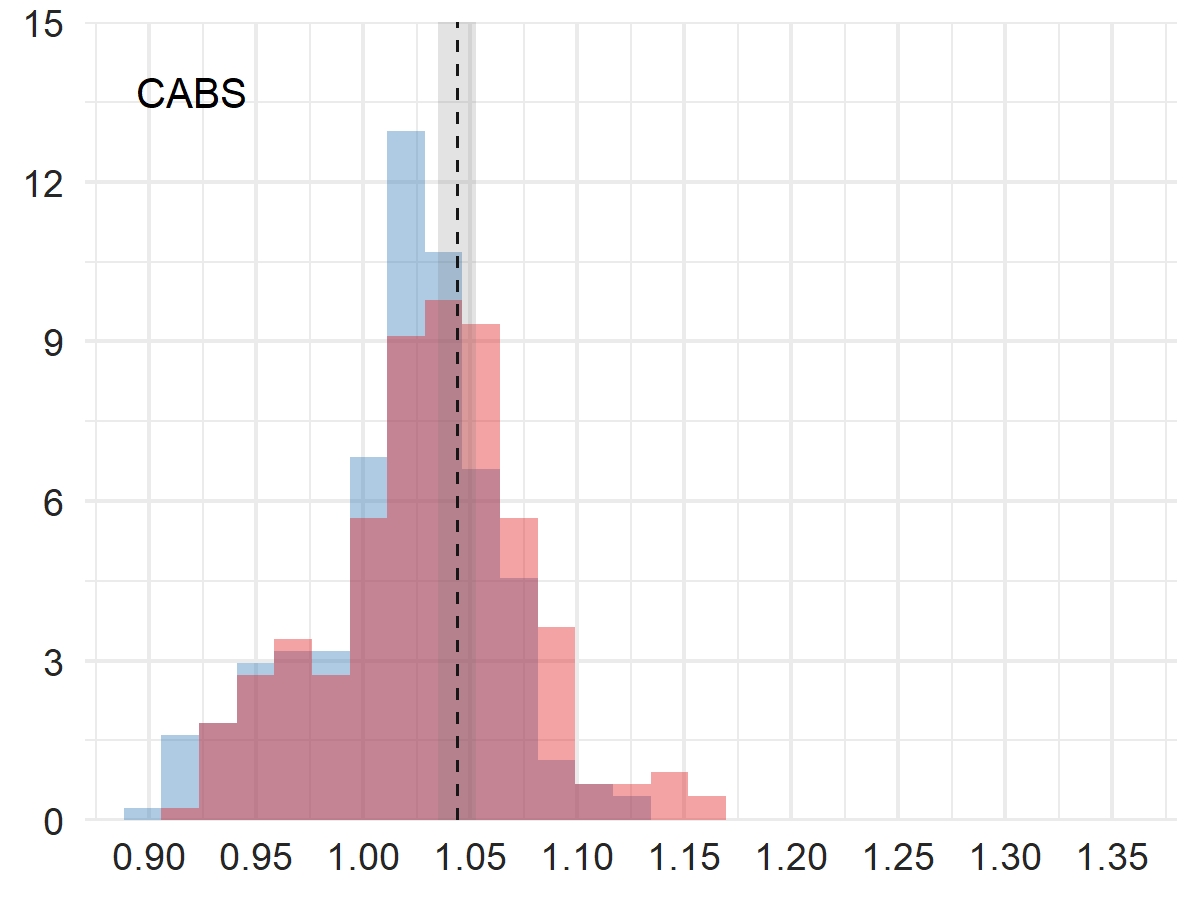

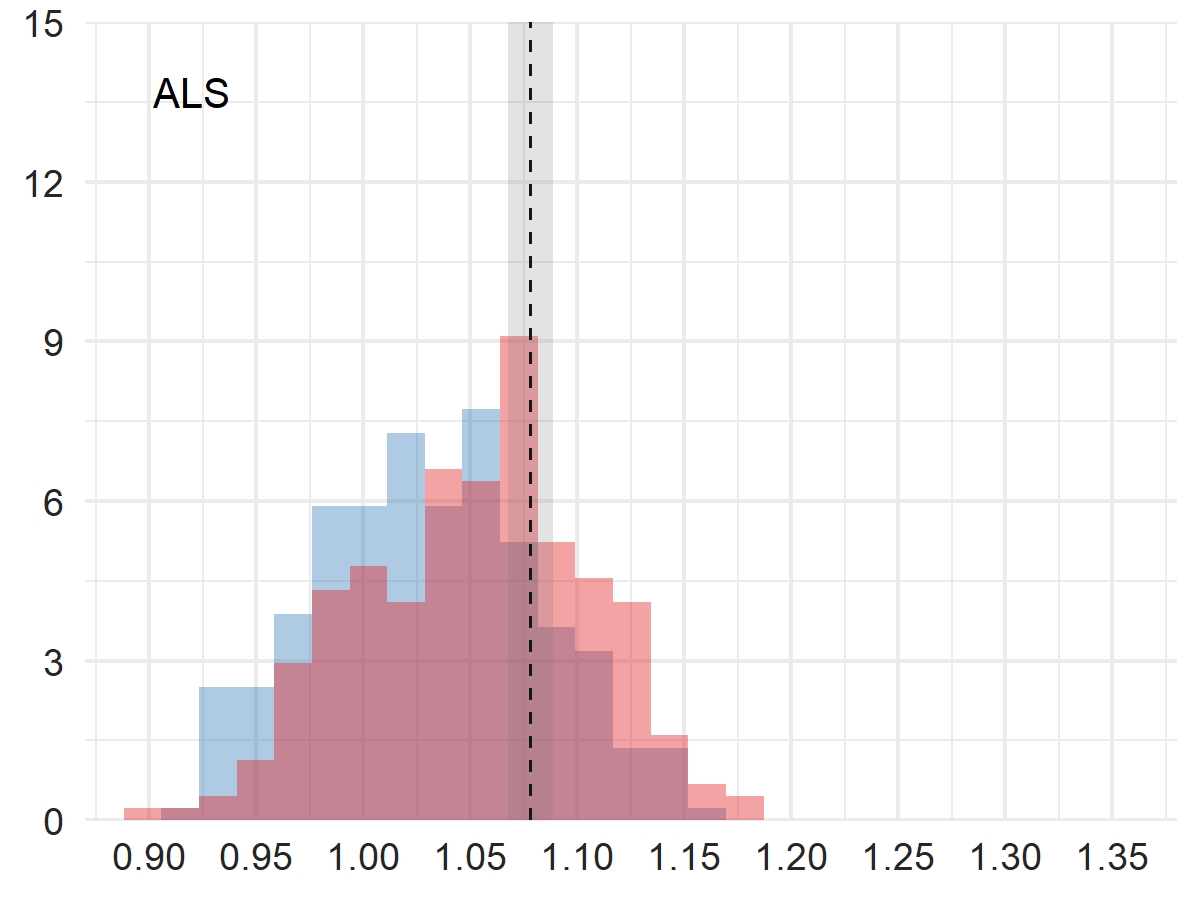

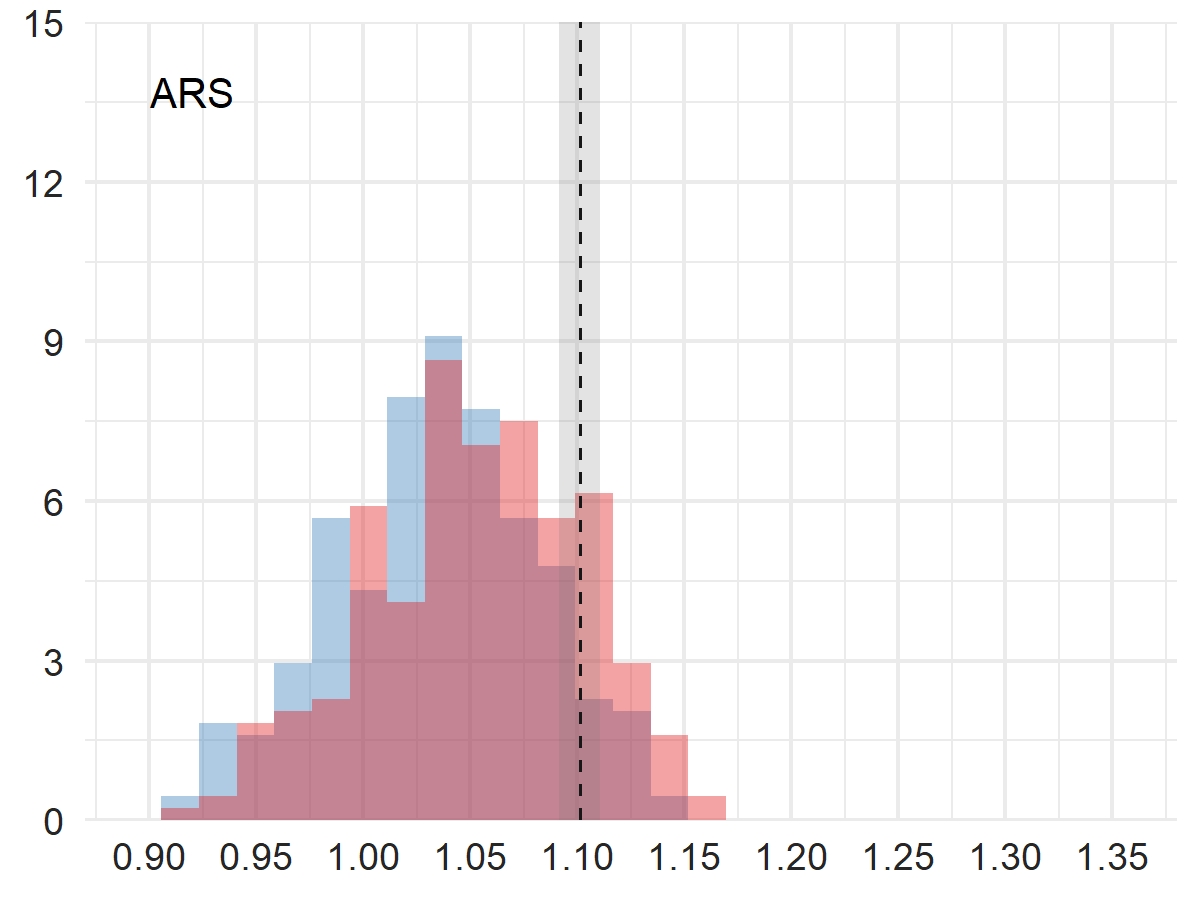

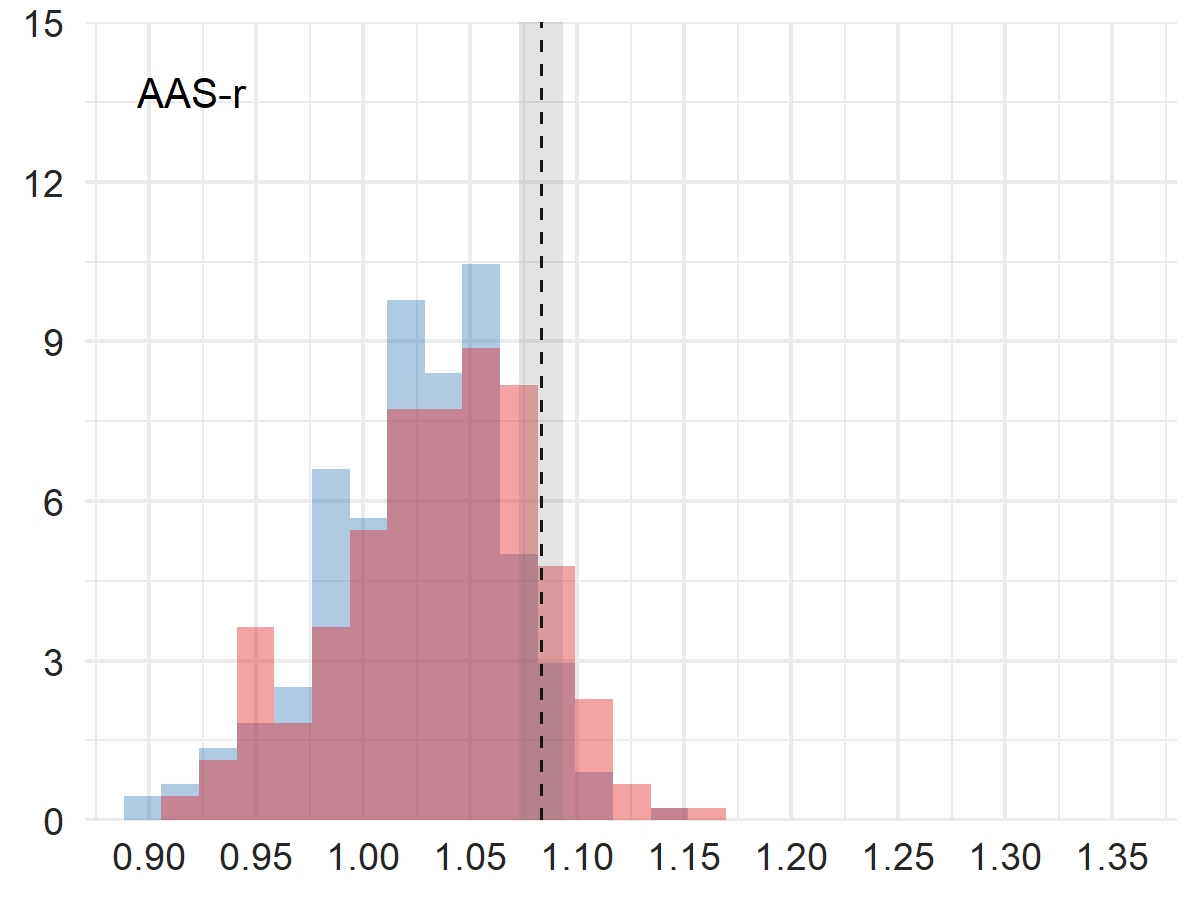

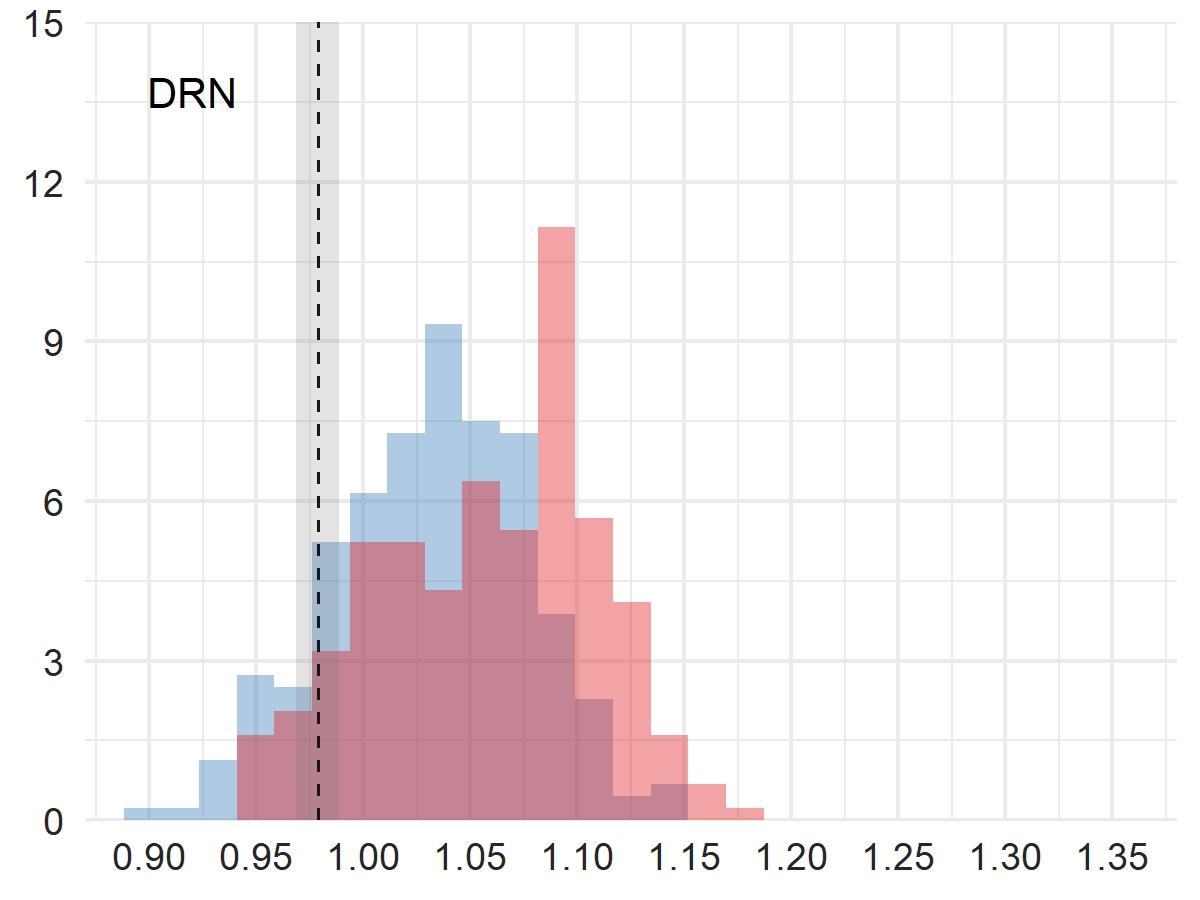

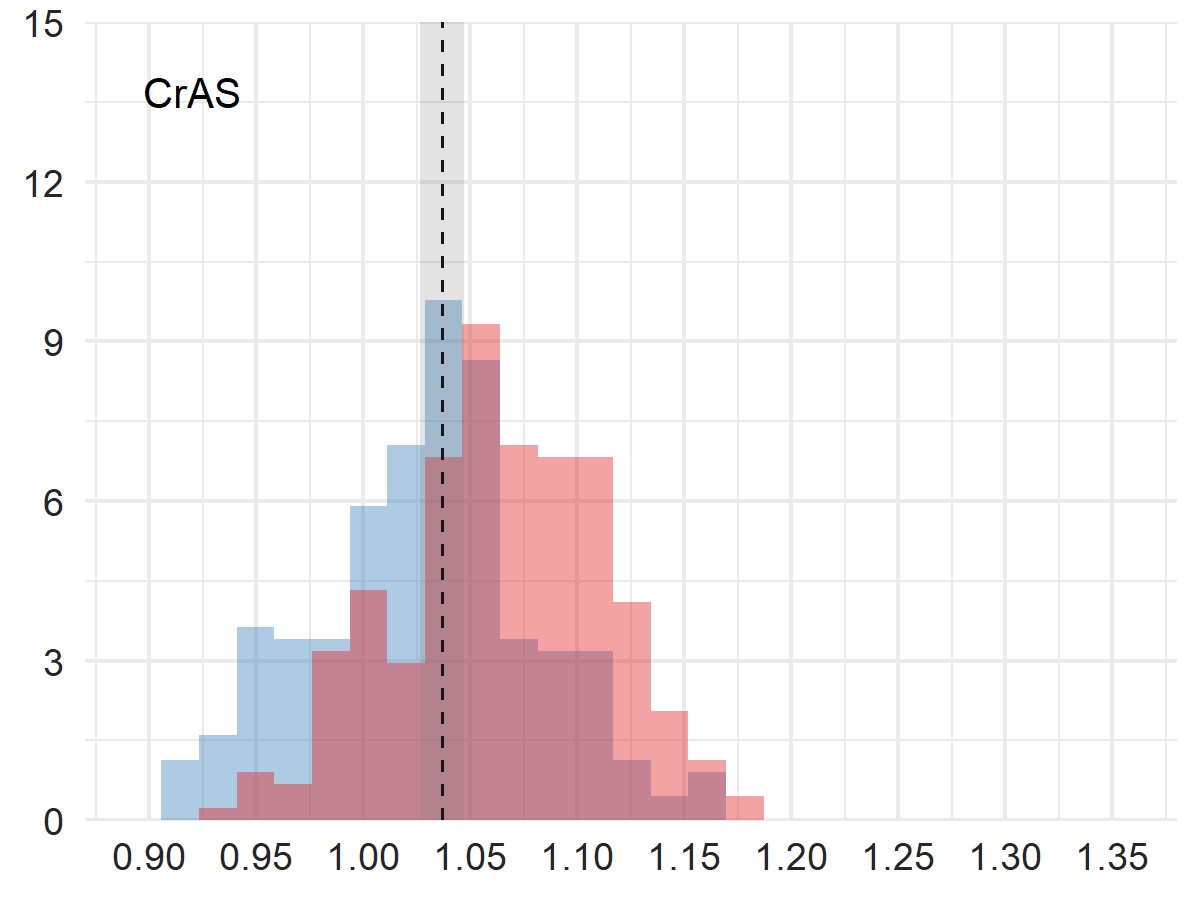

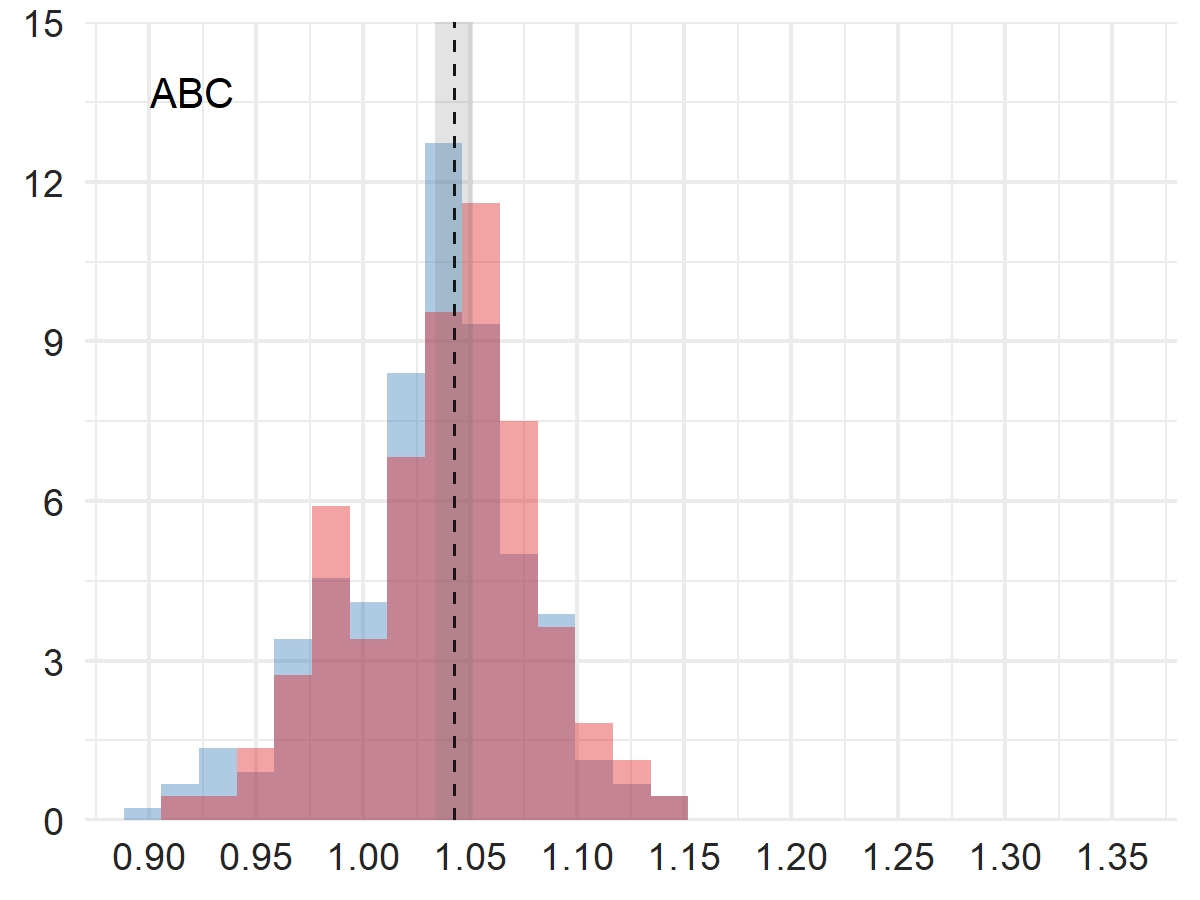

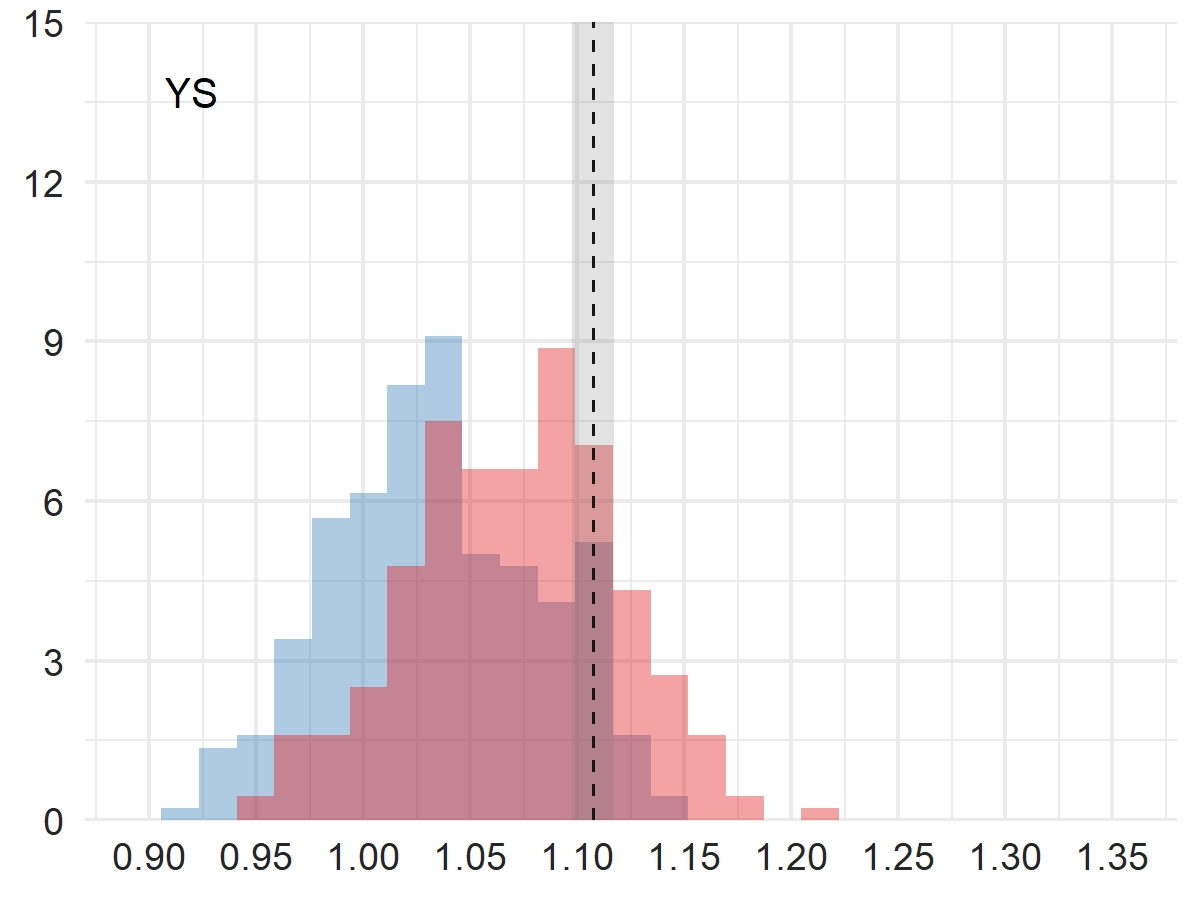

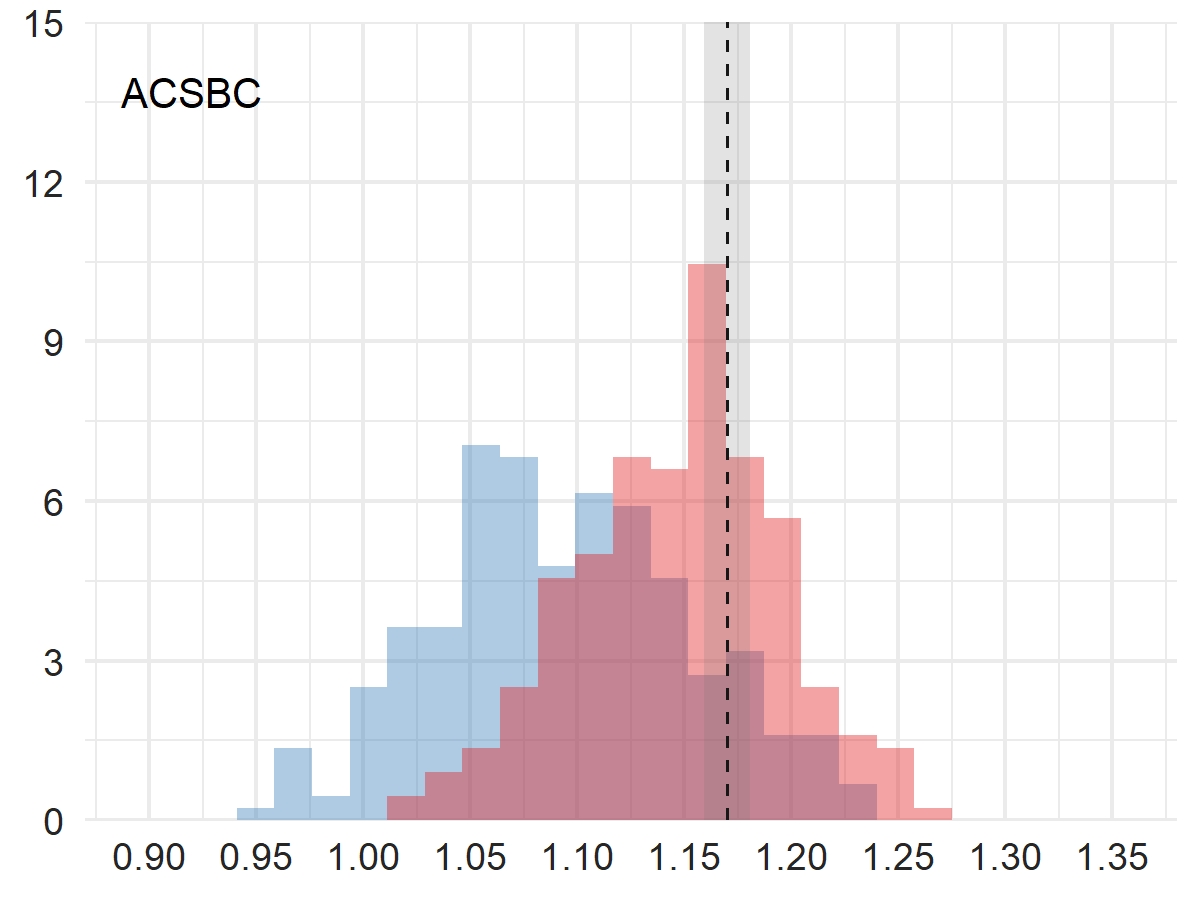

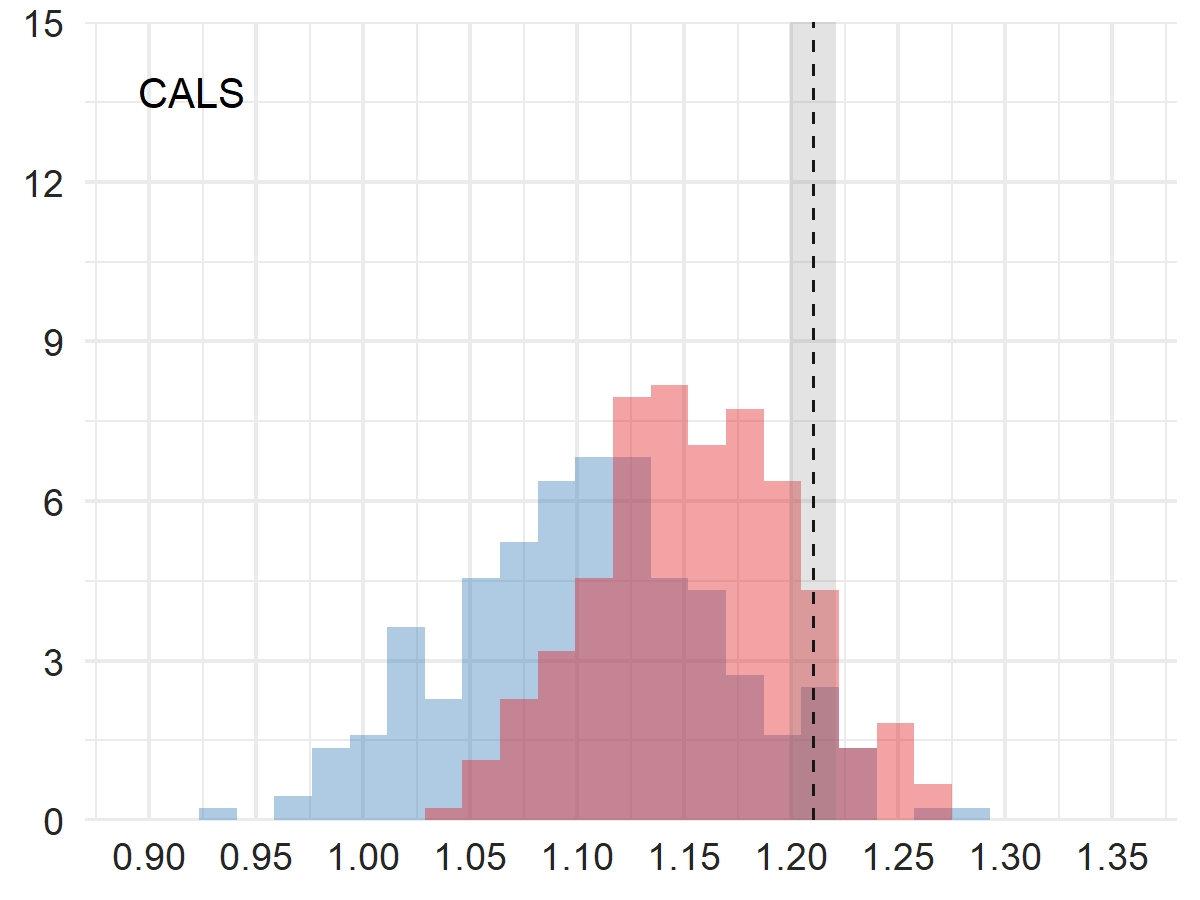

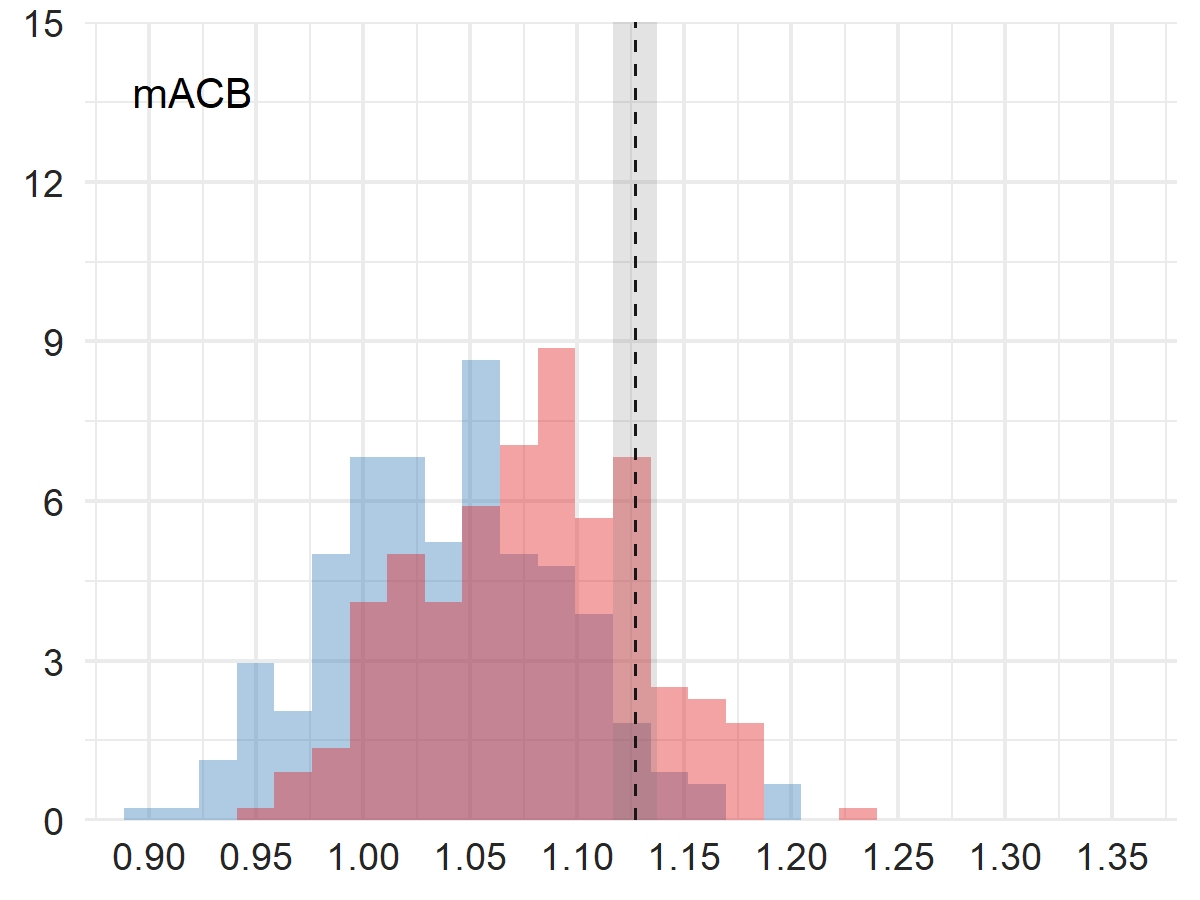

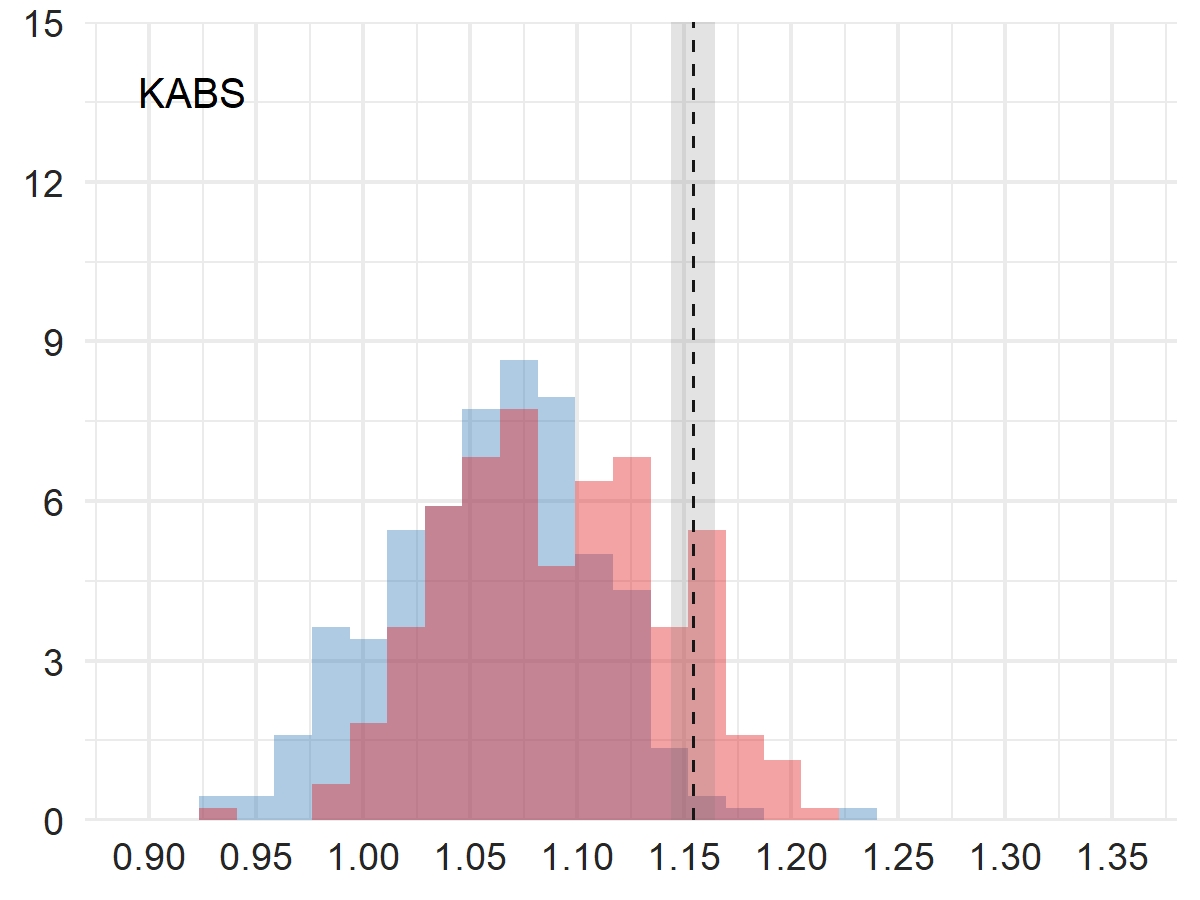

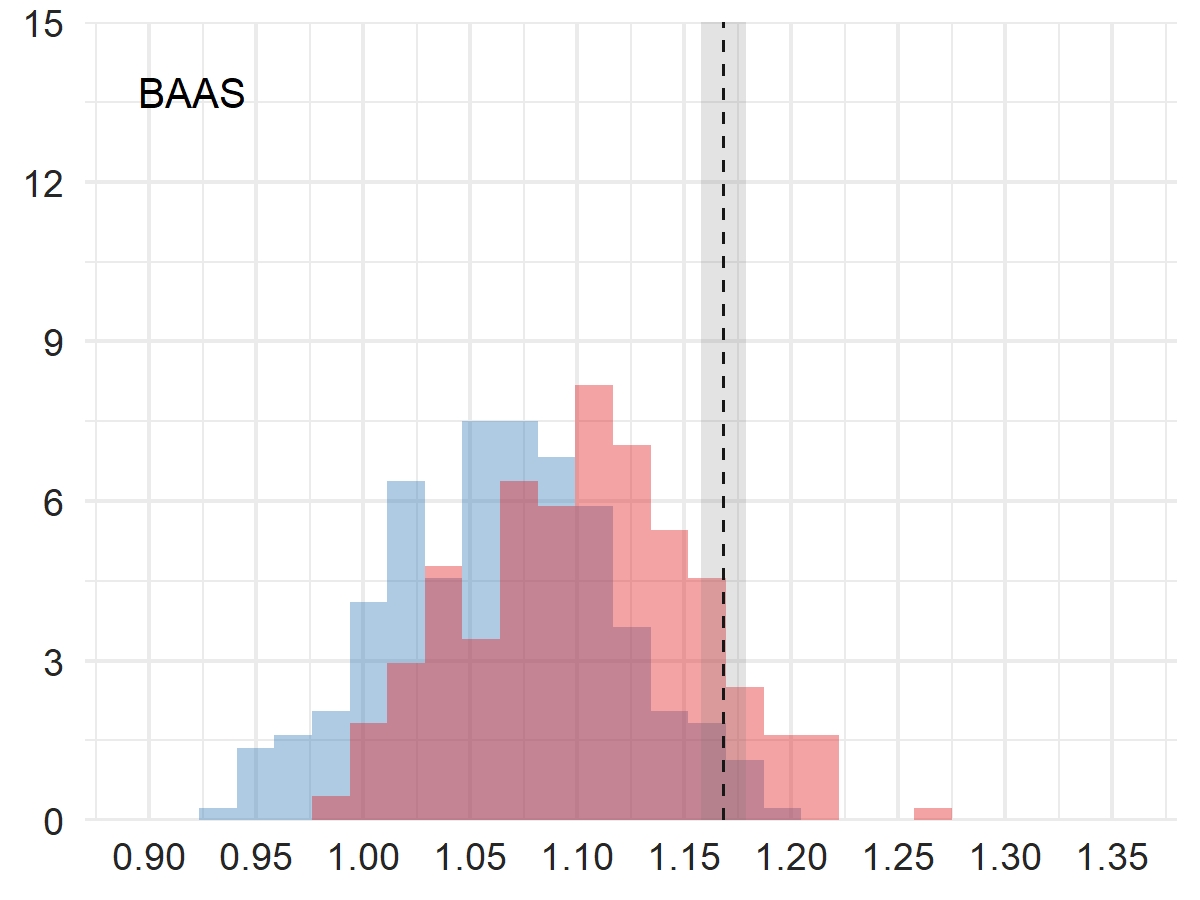

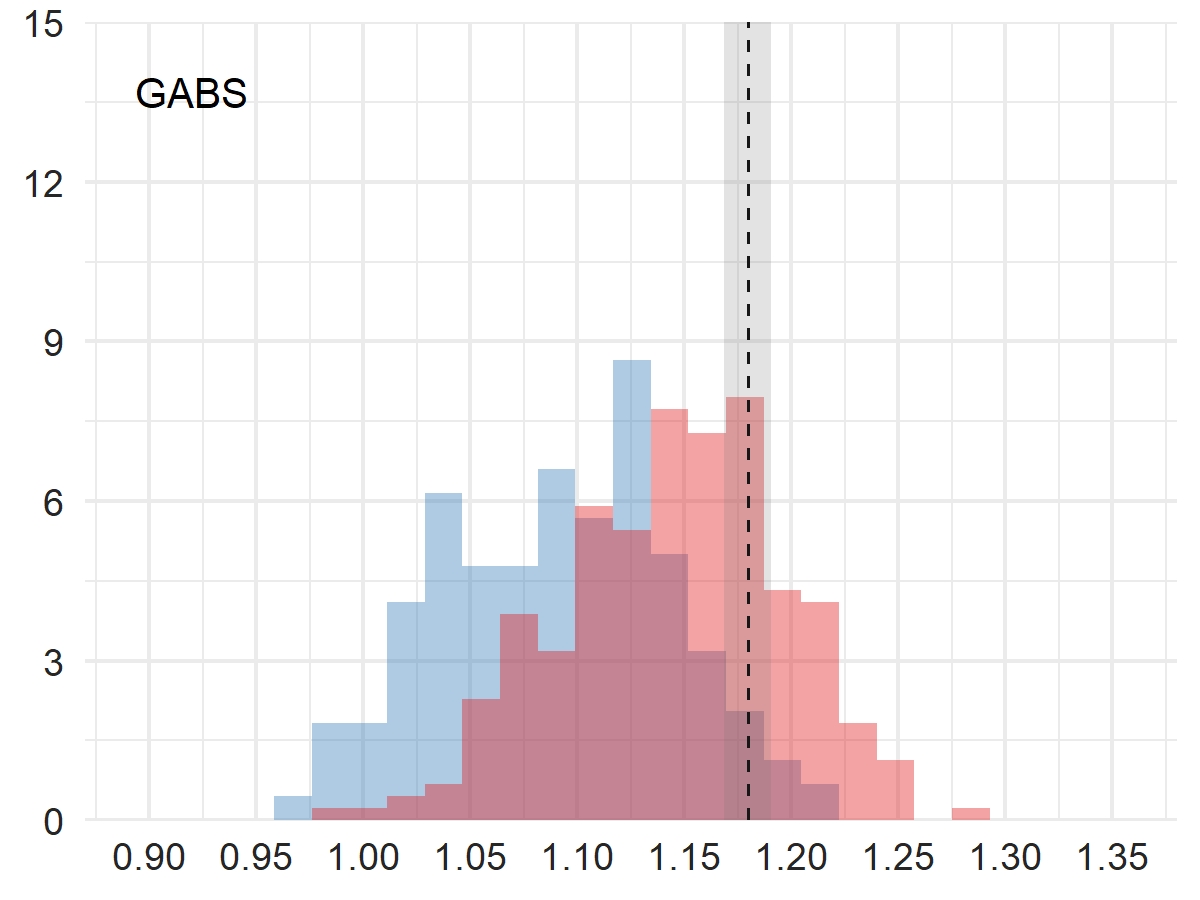

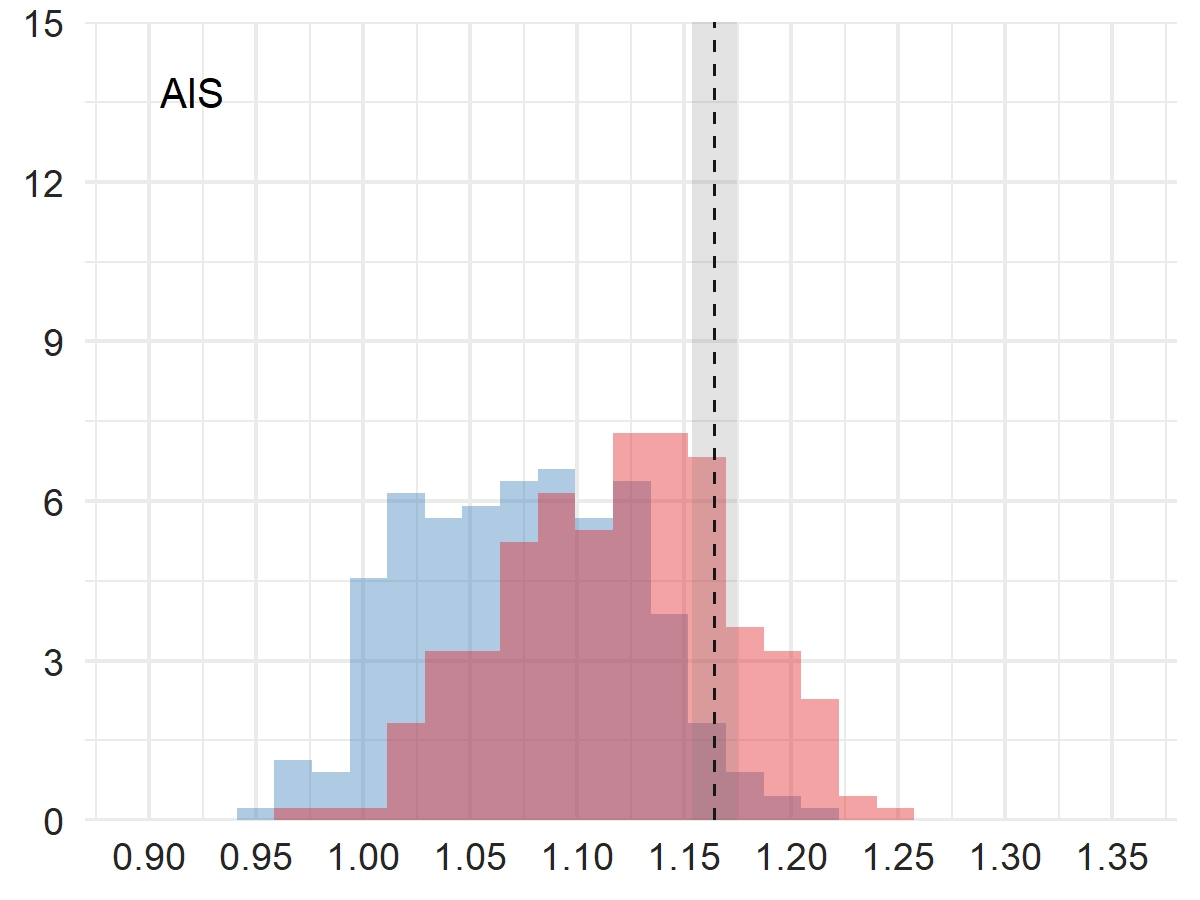

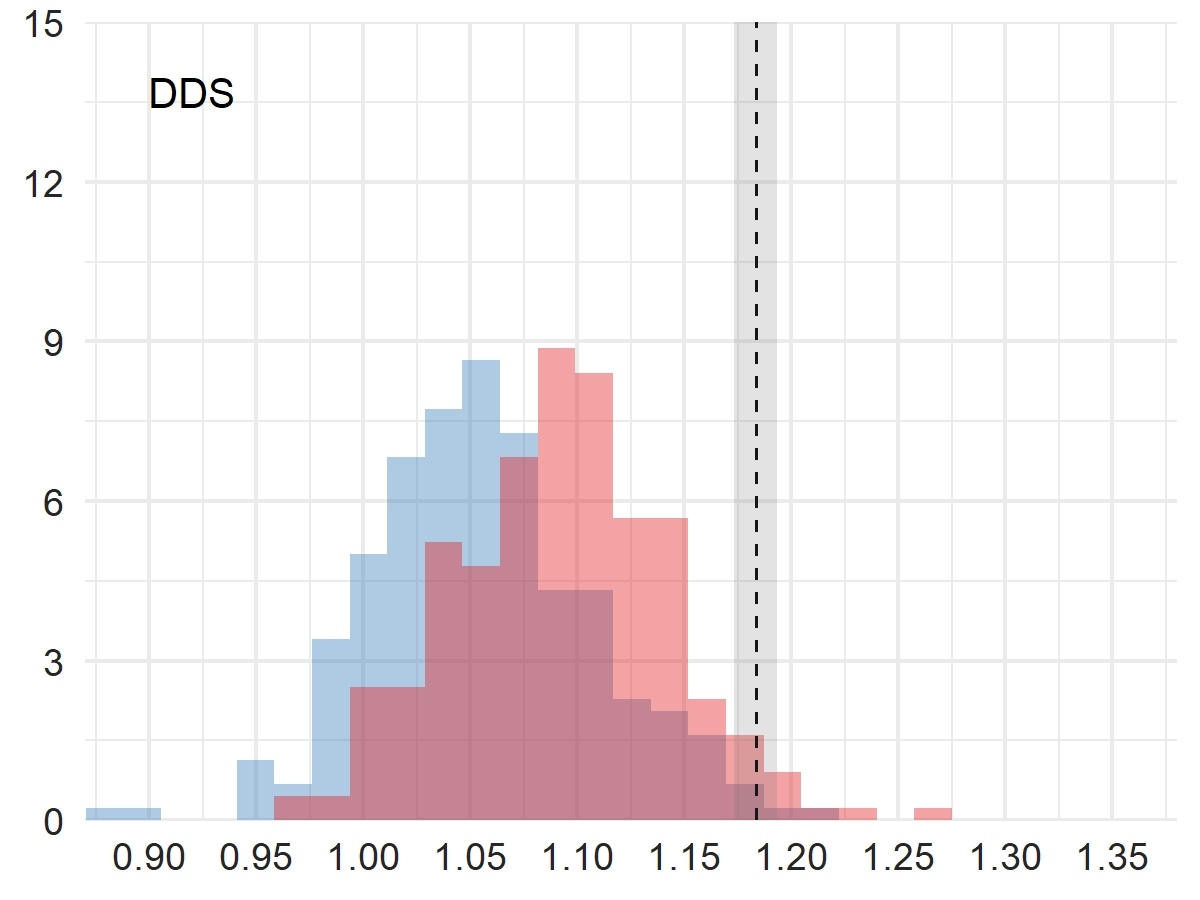

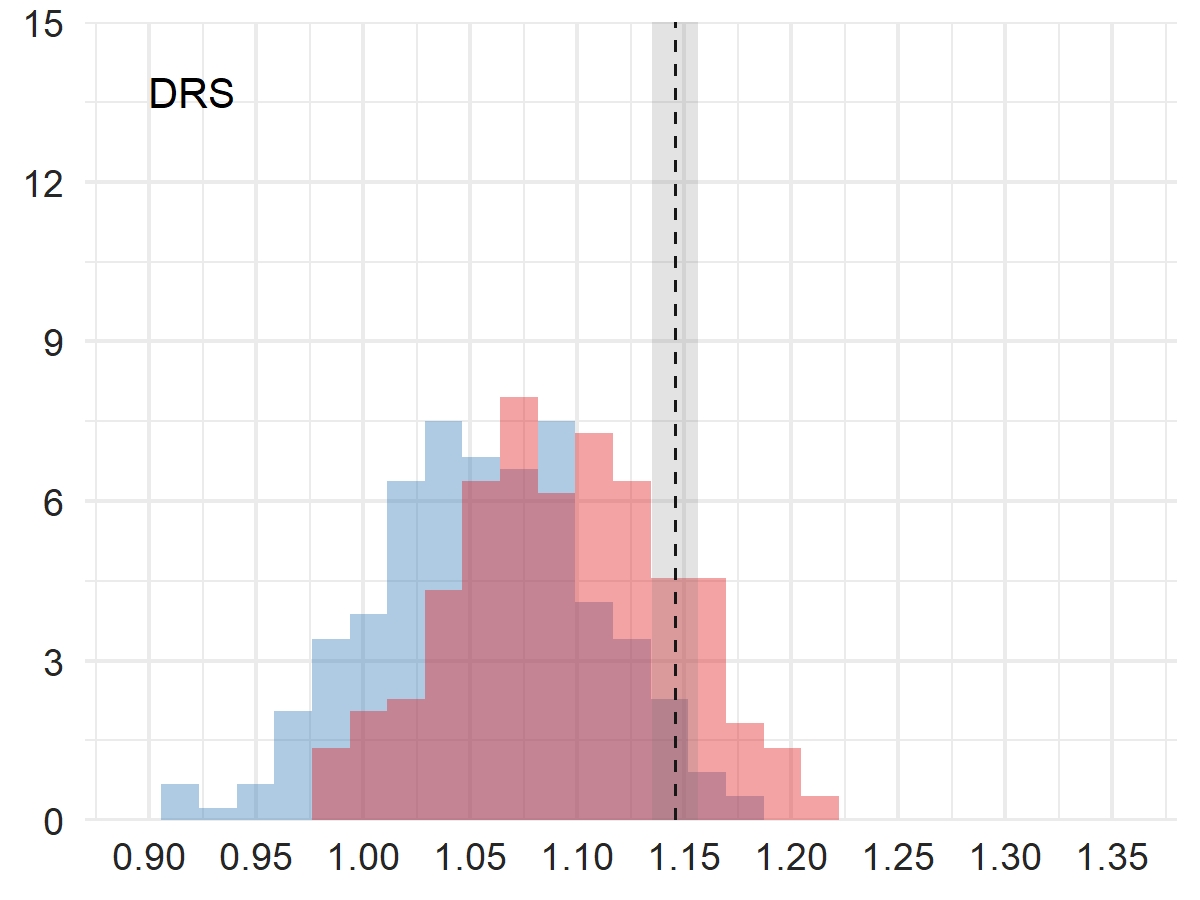

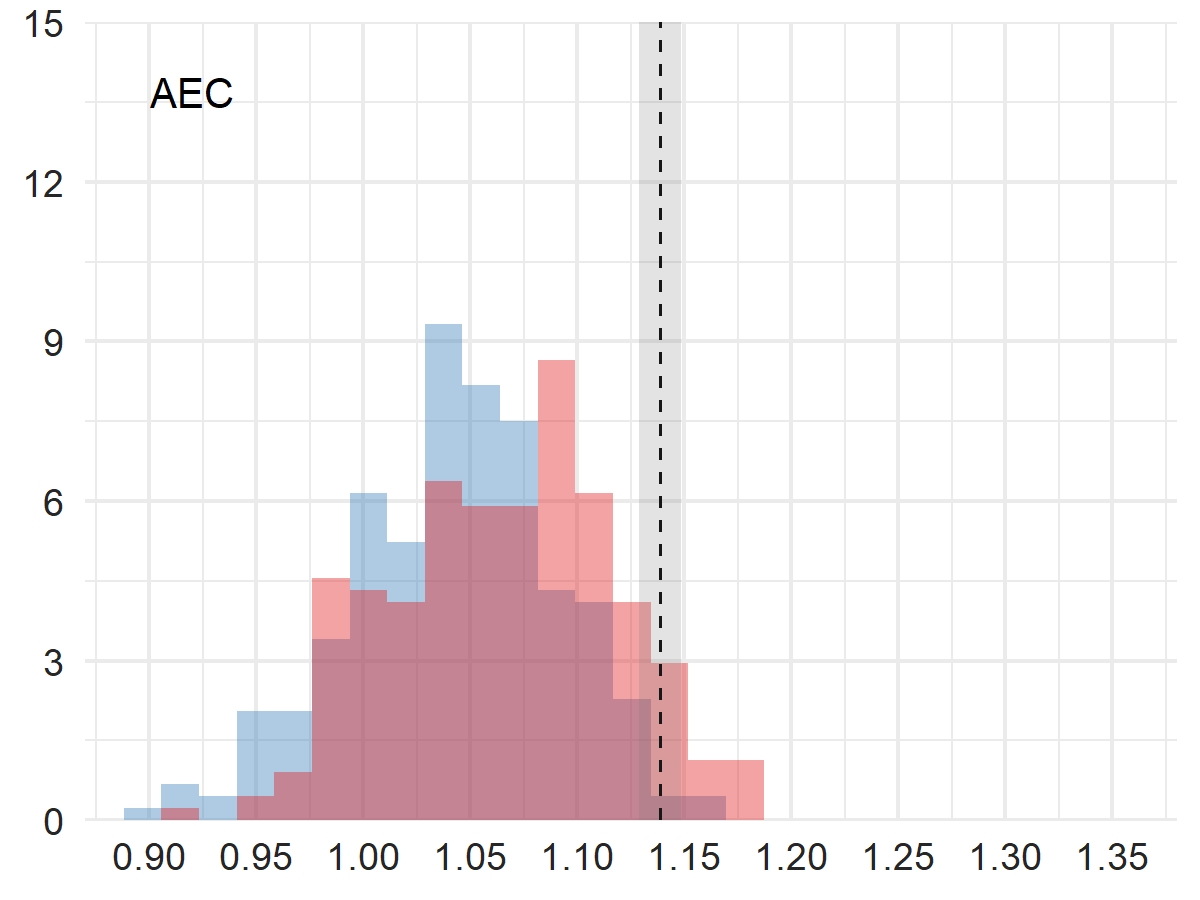

**Suppl. Table 6**: some statistical properties of general and anticholinergic within-sampling pseudoscales when evaluated for the different outcomes. The rows display the median proportion of pseudoscales that exhibited weaker effects than their corresponding ABS; the numbers (and %) of ABS that exhibited effect sizes greater than 50%, 68%, or 95% of their corresponding pseudoscales; the correlation between the above described median proportion and the size of the scale; and the correlation between the overlap between general and anticholinergic pseudoscales on the one hand, and the size of the scale on the other.

| **Outcome** | **Death** | | **Dementia** | | **Delirium** | |
| --- | --- | --- | --- | --- | --- | --- |
| **Pseudoscale type** | **general** | **antichol.** | **general** | **antichol.** | **general** | **antichol.** |
| **Median cum. prop.** | 0.73 | 0.46 | 0.92 | 0.71 | 0.94 | 0.82 |
| **> 50%, n (%)** | 18 (78%) | 10 (43%) | 21 (91%) | 16 (70%) | 22 (96%) | 21 (91%) |
| **> 68%, n (%)** | 12 (52%) | 7 (30%) | 20 (87%) | 13 (57%) | 20 (87%) | 17 (74%) |
| **> 95%, n (%)** | 1 (4%) | 1 (4%) | 7 (30%) | 1 (4%) | 11 (48%) | 2 (9%) |
| **R (cum. prop., n)** | 0.44 (95% CI: 0.04-0.72) | 0.09 (95% CI:  -0.34-0.48) | 0.41 (95% CI: 0-0.70) | 0.05 (95% CI:  -0.37-0.45) | 0.38 (95% CI:  -0.04-0.68) | 0.26 (95% CI:  -0.17-0.61) |
| **R (overlap, n)** | -0.88 (95% CI: -0.95- -0.74) | | -0.92 (95% CI: -0.97- -0.82) | | -0.86 (95% CI: -0.94- -0.69) | |

**Suppl. Text 2**: sensitivity analysis using the years 2004 to 2006; description of properties of across-sampling pseudoscales when evaluated for the three different outcomes.

In across-sampling, the ORs ranged from 0.97 to 1.10 (death), from 0.95 to 1.13 (dementia), and from 0.94 to 1.20 (delirium) for general pseudoscales and from 0.99 to 1.13 (death), from 0.94 to 1.13 (dementia), and from 0.91 to 1.15 (delirium) for anticholinergic pseudoscales. The 95% simulation intervals (SI) that express the values between which 95% of ORs are situated, were similar between general and anticholinergic pseudoscales (**Suppl.** **Table 10**). The distributions of ORs exhibited an overlap of 0.78, 0.77, and 0.95 for death, dementia, and delirium, respectively. Anticholinergic pseudoscales had larger effect sizes on average than general pseudoscales for all outcomes, and similar variability for death and delirium, but not for dementia (**Suppl. Table 8**). At the 95% CI, the association between the drug burden and the outcome would have been significant for 857/1000, 848/1000, and 768/1000 general pseudoscales for death, dementia, and delirium respectively. For anticholinergic pseudoscales, these numbers were 922/1000, 943/1000, and 763/1000. In other words, the association between drug burden and the outcome had an 8% (death), 11% (dementia), and 1% (delirium) higher probability of being significant if the burden scale was constructed using only putatively anticholinergic drugs as opposed to all drugs. The correlation between the OR and the scale size (i.e., number of drugs included on a scale) for ABS was r=0.77 (95% CI=0.52-0.90), r=0.72 (95% CI=0.44-0.87), and r=0.28 (95% CI=-0.15-0.62) for death, dementia, and delirium, respectively. The correlation of effect size with scale size for pseudoscales is listed in **Suppl.** **Table 8**.

**Suppl. Table 8**: sensitivity analysis using the years 2004 to 2006; comparisons between general and anticholinergic pseudoscales in their respective effects of association with the outcomes. The rows depict for all three outcomes the 95% SI, mean OR, standard deviation (SD), the correlation between the effect size (of the association between drug burden and the outcome) and scale size, and the results of the paired T-test of the difference between the means of the ORs of general and anticholinergic pseudoscales.

| **Metric** | **Death** | | **Dementia** | | **Delirium** | |
| --- | --- | --- | --- | --- | --- | --- |
|  | **general** | **antichol.** | **general** | **antichol.** | **general** | **antichol.** |
| 95% SI | 1.00-1.08 | 1.00-1.10 | 0.99-1.09 | 1.00-1.11 | 0.97-1.12 | 0.96-1.12 |
| Mean | 1.03 | 1.05 | 1.04 | 1.05 | 1.04 | 1.04 |
| SD | 0.022 | 0.026 | 0.027 | 0.028 | 0.039 | 0.039 |
| R (95% CI) | 0.57  (0.83-0.61) | 0.70 (0.67-0.73) | 0.46  (0.41-0.51) | 0.64 (0.61-0.68) | 0.49 (0.45-0.54) | 0.38 (0.32-0.43) |
| T-test  (T, p) | 15.9, <0.001 | | 15.1, <0.001 | | 0.01, 0.99 | |

**Suppl. Table 9**: sensitivity analysis using the years 2004 to 2006; some statistical properties of general and anticholinergic within-sampling pseudoscales when evaluated for the different outcomes. The rows display the median proportion of pseudoscales that exhibited weaker effects than their corresponding ABS; the numbers (and %) of ABS that exhibited effect sizes greater than 50%, 68%, or 95% of their corresponding pseudoscales; the correlation between the above described median proportion and the size of the scale; and the correlation between the overlap between general and anticholinergic pseudoscales on the one hand, and the size of the scale on the other.

| **Outcome** | **Death** | | **Dementia** | | **Delirium** | |
| --- | --- | --- | --- | --- | --- | --- |
| **Pseudoscale type** | **general** | **antichol.** | **general** | **antichol.** | **general** | **antichol.** |
| **Median cum. prop.** | 0.74 | 0.54 | 0.78 | 0.56 | 0.71 | 0.67 |
| **> 50%, n (%)** | 19 (83%) | 13 (57%) | 20 (87%) | 13 (57%) | 15 (65%) | 15 (65%) |
| **> 68%, n (%)** | 15 (65%) | 5 (22%) | 15 (65%) | 7 (30%) | 12 (52%) | 11 (48%) |
| **> 95%, n (%)** | 2 (9%) | 0 | 0 | 0 | 3 (13%) | 2 (9%) |
| **R (cum. prop., n)** | 0.30 (95% CI: -0.13-0.63) | 0.05 (95% CI: -0.45-0.37) | 0.16 (95% CI: -0.27-0.54) | -0.14 (95% CI: -0.52-0.29) | -0.27 (95% CI: -0.62-0.16) | -0.22 (95% CI: -0.58-0.21) |
| **R (overlap, n)** | -0.92 (95% CI: -0.97- -0.82) | | -0.81 (95% CI: -0.92- -0.59) | | -0.37 (95% CI: -0.05- -0.68) | |

1. Centre for Genomic and Experimental Medicine, University of Edinburgh, Edinburgh, UK.
2. Centre for Clinical Brain Sciences, University of Edinburgh, Edinburgh, UK.

   Global Brain Health Institute, University of California San Francisco, San Francisco, USA
3. Heritage College of Osteopathic Medicine, Ohio University, Athens, Ohio, USA.
4. Department of Social Sciences, Institute for Research on Socio-Economic Inequality (IRSEI), University of Luxembourg, Esch-sur-Alzette, Luxembourg.
